# A Mendelian randomization study investigating the role of sleep traits and their joint effects on the incidence of atrial fibrillation

**DOI:** 10.1101/2024.05.15.24307169

**Authors:** Nikhil Arora, Ben Michael Brumpton, Bjørn Olav Åsvold, Jan Pål Loennechen, Vegard Malmo, Laxmi Bhatta, Eivind Schjelderup Skarpsno, Rebecca Claire Richmond, Linn Beate Strand

**Affiliations:** HUNT Center for Molecular and Clinical Epidemiology, Department of Public Health and Nursing, Norwegian University of Science and Technology, Trondheim, Norway; Population Health Sciences, Bristol Medical School, University of Bristol, Bristol, United Kingdom; HUNT Research Centre, Department of Public Health and Nursing, Norwegian University of Science and Technology, Levanger, Norway; Department of Medicine, St. Olavs Hospital, Trondheim, Norway; Department of Endocrinology, Clinic of Medicine, St. Olavs Hospital, Trondheim, Norway; Department of Circulation and Medical Imaging, Norwegian University of Science and Technology, Trondheim, Norway; Clinic of Cardiology, St. Olavs Hospital, Trondheim, Norway; FIU-PH, Division of Mental Health Care, St. Olavs Hospital, Trondheim, Norway; Department of Clinical and Molecular Medicine, Norwegian University of Science and Technology, Trondheim, Norway; Department of Public Health and Nursing, Norwegian University of Science and Technology, Trondheim, Norway; Department of Neurology and Clinical Neurophysiology, St. Olavs Hospital, Trondheim, Norway; MRC Integrative Epidemiology Unit, University of Bristol, Bristol, United Kingdom; NIHR Oxford Health Biomedical Research Centre, University of Oxford, Oxford, United Kingdom

**Keywords:** Atrial fibrillation, insomnia, sleep duration, chronotype and Mendelian randomization

## Abstract

**Background and Aims:** Sleep disturbances can induce alterations in functional and electrical properties of the heart, thereby increasing susceptibility to atrial fibrillation (AF). We aimed to test the causal role of different sleep traits and their joint effects on the risk of AF.

**Methods:** We used an observational cohort study design along with one-sample and factorial Mendelian randomization (MR) approaches to test for individual and joint associations of sleep traits (i.e., insomnia symptoms, sleep duration and chronotype) on the risk of AF using UK Biobank and the second survey of the Trøndelag Health Study (HUNT2).

**Results:** Both the observational and the MR analysis showed that insomnia symptoms and genetic predisposition to insomnia symptoms increased risk of AF; however these MR findings were not consistent in HUNT2. One-sample MR analysis in UK Biobank showed that genetic predisposition to short sleep duration increased risk of AF and genetic predisposition to per additional hour of sleep decreased risk of AF; however these findings were not consistent in HUNT2 or in observational analysis. Factorial MR analysis in UK Biobank showed that participants with genetic predisposition to two sleep traits in combination (including insomnia symptoms, short sleep duration, and a morning chronotype) had higher risk of AF than those with genetic predisposition to only one sleep trait; however these findings were not consistent in HUNT2 or in observational analysis. We found no evidence of relative excess risk due to interaction (RERI) for any combinations of sleep traits.

**Conclusions:** Our study indicates that insomnia symptoms and short sleep duration are causal risk factors for AF. However, having two sleep traits in combination did not increase risk beyond the additive risk of each individual trait. This reinforces clinical and public health efforts to effectively manage insomnia symptoms and short sleep, in order to mitigate the risk of AF and improve overall cardiovascular health.

## Introduction

Atrial fibrillation (AF) is the most common sustained cardiac arrythmia, estimated to have affected 59.7 million individuals globally by 2019.^1^ Accompanying the population aging in the west and improved survival with chronic ailments, AF is emerging as a global epidemic.^2,3^ AF is linked with increased morbidity, mortality and substantial healthcare expenditures,^4,5^ underpinning the need for research efforts aimed at identifying modifiable risk factors that can be targeted to improve prevention and treatment strategies.

Sleep is acknowledged as an essential modulator of cardiovascular function, possibly due to autonomic dysregulation and hormonal disturbances.^6,7^ Sleep deprivation can increase the QT interval and atrial electromechanical delay,^8,9^ suggesting that it may induce changes in functional and electrical properties of the heart and promote the occurrence of arrythmias. A previous observational study identified insomnia as an individual risk factor for AF (hazard ratio (HR) 1.14; 95% confidence interval (CI) 1.08, 1.21).^10^ Moreover, a meta-analysis on cohort studies provided strong evidence that short (≤6 h) and long (≥9 h) sleep durations were associated with increased risk of AF (HR 1.21; 95% CI 1.02, 1.44 and HR 1.18; 95% CI 1.03, 1.35, respectively).^11^ While studies found increased risk of myocardial infarction in people with evening chronotype (characterized by staying up late and/or waking up late),^12,13^ no study has examined potential impact of chronotype on the risk of AF.

Sleep is a complex and multifaceted biological phenomenon comprising several traits.^14^ Some studies have found evidence that sleep traits interact to increase the risk of cardiovascular outcomes.^12,15–17^ In particular, insomnia with short sleep duration, considered the most biologically severe sleep disorder phenotype,^18^ has been associated with increased cardiometabolic risk.^19,20^ It is therefore plausible that individuals experiencing poor sleep, characterized by a combination of extreme sleep traits, may have higher incidence of AF.

Disentangling causal from spurious associations can be challenging in conventional observational studies of sleep traits due to confounding, reverse causation, and measurement error bias.^21^ For example, obesity and smoking status are possible common causes of sleep traits and cardiovascular disease, and not all possible confounders are known or measured. Mendelian randomization (MR) uses genetic variants robustly associated with an exposure (sleep traits) as instrumental variables to investigate causal effects on an outcome (AF).^22^ MR relies on the fact that genetic variants are randomly assigned at conception, therefore allocating individuals to the exposure group based on genotype (analogous to treatment assignment in a randomized controlled trial). Given the random and independent assignment of alleles at conception, bias arising from reverse causation can be avoided, and the impact of confounding can be reduced. Moreover, genetically-determined sleep traits may be more reflective of sleep behaviours across the lifespan, thereby decreasing measurement error compared to a single self-reported measurement.^22^

Recent MR studies have suggested that insomnia symptoms have an adverse effect on AF (odds ratio (OR) ranged from 1.04 to 1.13), as did short sleep duration (≤6 h) (OR ranged from 1.11 to 1.13).^23–27^ Whereas longer sleep duration had a protective effect on AF (OR ranged from 0.68 to 0.89 per hour increase).^26,27^ Chronotype has not been explored in relation to AF using MR. Further, MR investigations are warranted to investigate the joint effects of sleep traits on incidence of AF, which could provide robust evidence on the causal risk of AF from concomitant exposure to two sleep traits.^28^

The aim of this study was to examine association of individual sleep traits (insomnia symptoms, sleep duration and chronotype) and their joint effects on the risk of AF using data from the UK Biobank and the second survey of the Trøndelag Health Study (HUNT2). We compared findings from observational and MR analyses, to determine possible causality of individual sleep traits and their joint effects on the incidence of AF.

## Methods

### Study design and population

We conducted parallel prospective cohort and factorial MR studies in two large cohorts – UK Biobank (2006-10) and HUNT2 (1995-97). Owing to the non-availability of sleep duration in HUNT1 and HUNT3, and short follow-up in HUNT4, we chose to use HUNT2. Over 500 000 adults (5.5% of the invited) ranging between 40 and 70 years participated in UK Biobank,^29^ and 65 228 adults (69.5% of the invited) aged 20 years or older participated in HUNT2.^30^ All participants provided written informed consent. The sample in the present study were restricted to participants with European decent, as genetic variants used for MR analyses were investigated in genome-wide association studies (GWAS) with European ancestry. This enabled comparison of observational and MR estimates within the same underlying population. UK Biobank was approved by the National Health Service (NHS) Research Ethics Service (reference number 11/NW/0382). The HUNT Study has ethical clearance from the Data Inspectorate of Norway and HUNT2 was approved by the Regional Committee for Ethics in Medical Research (REK; reference number 152/95/AH/JGE). Approval to conduct this study was also received from the Regional Committee for Ethics in Medical Research (REK nord; reference number 2020/47206).

### Sleep traits

#### Insomnia symptoms

In UK Biobank, participants were asked a single question if they have trouble falling asleep at night or if they wake up in the middle of the night with response options “never/rarely”, “sometimes”, “usually” or “prefer not to answer”. Those who responded “usually” were classified as having insomnia symptoms, and “sometimes” or “never/rarely” as not having insomnia symptoms. Other responses were coded as missing.

In HUNT2, participants were asked two questions: (1) if they had difficulty falling asleep in the last month, and (2) if they have woken too early and not been able to get back to sleep during the last month, both with the response options “never”, “sometimes”, “often” or “almost every night”. Participants who responded “often” or “almost every night” to both of these questions were classified as having insomnia symptoms. For participants who answered only one of these questions, we did the following: (1) if they responded “often” or “almost every night” to one of these, they were classified as having insomnia symptoms, and (2) if they responded “never” or “sometimes” to one of these, but did not answer the other, they were excluded to avoid possible misclassification. The remaining participants were classified as not having insomnia symptoms.

Our definition of insomnia included two night-time insomnia symptoms (i.e., difficulty falling asleep, difficulty maintaining sleep or waking up too early) without information about daytime impairment. Since this definition does not fulfil the clinical criteria to be diagnosed with insomnia,^31^ we used the term *insomnia symptoms* throughout this manuscript.

#### Sleep duration

In UK Biobank, participants were asked about the number of hours of sleep they get every 24 h (including their naps). In HUNT2, participants were asked about the number of hours they usually spend lying down (i.e. sleeping and/or napping) during a 24-h period. In both cohorts, the answers could only contain integer values. Implausible short or long sleep durations were avoided by excluding extreme responses of less than 3 h or more than 18 h. Participants were categorized into normal (7-8 h), short (≤6 h) or long (≥9 h) hours of sleep duration. Binary variables for short (≤6 h vs. 7-8 h) and long (≥9 h vs. 7-8 h) sleep durations were constructed.

#### Chronotype

Chronotype (diurnal preference) was only reported in UK Biobank. Participants were asked if they consider themselves to be “definitely a ‘morning’ person”, “more a ‘morning’ than ‘evening’ person”, “more an ‘evening’ than a ‘morning’ person”, “definitely an ‘evening’ person”, “do not know”, or “prefer not to answer”. Participants were classified as having a morning chronotype if they reported “definitely a ‘morning’ person” or “more a ‘morning’ than ‘evening’ person”, and as having an evening chronotype if they reported “more an ‘evening’ than a ‘morning’ person” or “definitely an ‘evening’ person”. Other responses were coded as missing.

### Genetic variants

DNA was extracted from blood samples collected by trained professionals in both cohorts. In UK Biobank, participants were genotyped using either one of the UK BiLEVE or the UK Biobank Axiom genotyping chips. The genetic variants used were from the UK Biobank imputed dataset (merged Haplotype Reference Consortium and UK10K plus 1000 Genomes phase 3 reference panels), that were quality controlled using a standard protocol.^32,33^ In HUNT2 and HUNT3, participants were genotyped with one of three different Illumina HumanCoreExome genotyping chips (HumanCoreExome 12 v.1.0, HumanCoreExome 12 v.1.1, and UM HUNT Biobank v.1.0), where genotypes from different chips were quality controlled separately and reduced to a common set of variants. The quality control measures used were similar to UK Biobank.^34^

A total of 248 single nucleotide polymorphisms (SNPs) were identified as robustly associated with insomnia symptoms at P < 5×10^−8^ based on the meta-analysis of UK Biobank (n = 386 533) and 23andMe (n = 944 477) cohorts in a GWAS by Jansen *et al.*^35^ A large GWAS performed by Dashti *et al.*^36^ based on UK Biobank (n = 446 118) identified 78 SNPs as robustly associated with 24-hour sleep duration. They additionally identified 27 SNPs specific for short and 8 SNPs specific for long sleep duration. A genome-wide association meta-analysis by Jones *et al.*^37^ based on UK Biobank (n = 403 195) and 23andMe (n = 248 100) identified 351 SNPs robustly associated with chronotype (morning preference). A comprehensive list with detailed information on discovery GWASs used to obtain the genetic instruments is provided (see supplementary material, Table S1).

### Ascertainment of atrial fibrillation (AF)

In UK Biobank, participants were linked to the National Health Service (NHS) central registers for health-related outcomes (Field IDs: 41270, 41271, 41280 and 41281) and mortality records (Field IDs: 40001 and 40000), and in HUNT2, via linkages to the medical records from the hospitals of the Nord-Trøndelag region and the National Cause of Death Registry.

Hospitalizations or deaths due to AF were identified using ICD-9 code 427.3 and ICD-10 code I48. Incident cases were defined as the first occurrence of either hospitalization or death due to AF during follow-up time. Each participant was followed until either first diagnosis/death due to AF, death due to other cause, loss to follow-up, or end of follow-up (March 23, 2021 for UK Biobank and December 31, 2020 for HUNT2), whichever came first.

### Covariates

The following covariates were considered as potential confounders: age, sex, marital status (married, unmarried or separated/divorced/widowed), frequency of alcohol intake (never, monthly, weekly, or daily), smoking history (never, ex-smoker or current smoker), body mass index (BMI), level of physical activity (inactive/low, moderate, or high), Townsend deprivation index (TDI; for UK Biobank only), education attainment (≤10 years, 11-13 years, or ≥14 years), shift work (yes or no), employment status (employed or not employed), systolic blood pressure (SBP), blood cholesterol levels, blood glucose levels, depression (yes or no in UK Biobank; and Hospital Anxiety and Depression Scale (HADS) – Depression scores in HUNT2), anxiety (yes or no in UK Biobank; and HADS – Anxiety scores in HUNT2), use of sleep medication (yes or no) and chronic illness (yes or no). The details on how covariates were handled are described in the supplementary methods.

### Statistical analysis

Figure 1 shows the participant selection. Among 441 565 participants in UK Biobank who had information available for sleep traits of interest, a total of 287 352 and 331 748 participants were included in the observational and MR analyses, respectively, after exclusions. Among 52 008 participants in HUNT2, 31 458 and 45 322 were included in the observational and MR analyses, respectively. Baseline characteristics of the study participants were presented by categories of AF diagnosis for study samples for both UK Biobank and HUNT2 cohorts.

**Figure 1:**
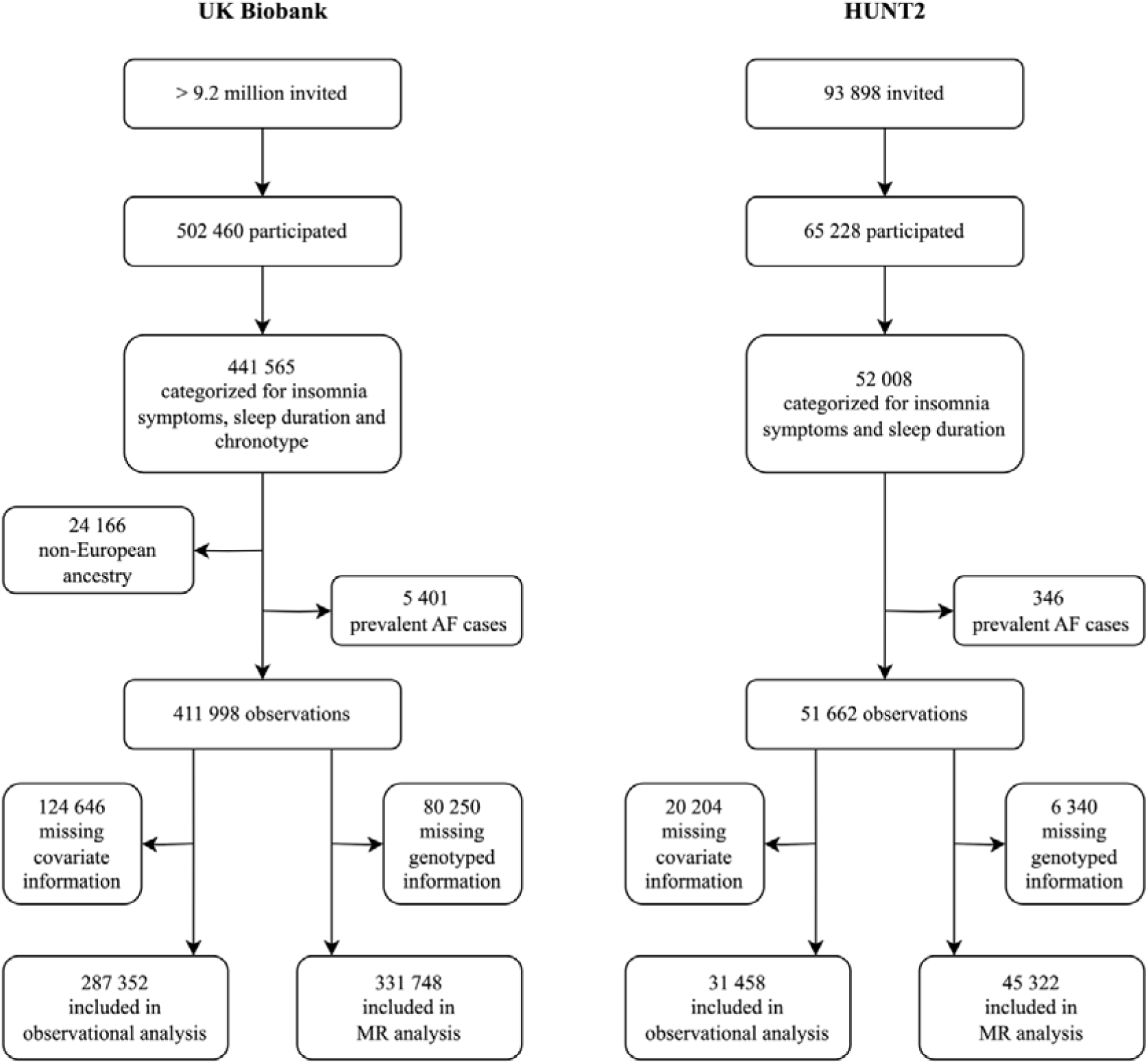
Flow chart of participant selection process.

#### Observational analysis

We used the Cox proportional hazard model to examine the associations of sleep traits on the risk of AF separately in UK Biobank and HUNT2. HRs with 95% CIs were calculated to estimate: (1) the associations of individual sleep traits on the risk of AF and, (2) the joint associations of any two sleep traits together on the risk of AF. The main model was adjusted for age at recruitment, sex, marital status, alcohol intake frequency, smoking status, BMI, physical activity, education, TDI (for UK Biobank only), shift work and employment status. Relative excess risk due to interaction (RERI) was assessed for the joint associations as a test of interaction, when none of the HRs were less than 1 (i.e. preventive).^38,39^ RERI equals 0 implies exact additivity (no interaction), RERI > 0 implies more than additivity (positive interaction or synergism), and RERI < 0 implies less than additivity (negative interaction or antagonism).

#### MR analysis

Genetic risk scores (GRS) were created as instruments for each sleep trait to overcome the weak individual effect of most SNPs on their corresponding sleep trait.^40^ Weighted GRS (wGRS) were computed by summing up the participants’ sleep trait increasing alleles (morning preference alleles for chronotype), weighted by the variant effect sizes from the external GWAS. We used wGRS for the main analysis in HUNT2 only, whereas in UK Biobank, we used unweighted GRS (uwGRS) computed simply by summing up the sleep trait increasing alleles. Since all included discovery GWASs used the UK Biobank cohort, the use of internal weights to calculate wGRS is not recommended.^40^ Instrument strength was assessed by regression of each sleep trait on their respective GRS and reporting variance explained (R^2^) and F-statistics.

MR requires meeting three core assumptions (see supplementary methods).^21,41^ We performed one-sample MR analysis to examine the causal effects of individual sleep traits on the risk of AF separately in UK Biobank and HUNT2. A two-stage predictor substitution (TSPS) regression estimator method was used to calculate average causal HRs with 95% CIs. The first stage involved regression of each sleep trait (linear regression for 24-hour sleep duration, and logistic regression for other sleep traits) on their GRS, and the second stage consisted of a Cox regression of AF status on the fitted values from the first stage regression, adjusted for age at recruitment, sex, assessment centre (in UK Biobank), genetic principal components (40 in UK Biobank and 20 in HUNT2), and genotyping chip in both stages. As recommended for MR analysis with a binary outcome, the first stage regression was limited to participants who did not experience AF.^42^ The causal estimates for binary exposures (insomnia symptoms, short sleep, long sleep and chronotype) were scaled to represent the risk increase in AF per doubling in the odds of these exposures, by multiplying the obtained β values by 0.693 as previously recommended.^43^ The causal estimate for sleep duration indicates the risk increase in AF per additional hour of sleep.

Further, we performed a 2×2 factorial MR analysis to examine the joint causal effects of sleep traits on the risk of AF separately in UK Biobank and HUNT2. Participants were dichotomized across their median GRS (uwGRS for UK Biobank and wGRS for HUNT2) for each sleep trait, with the group equal to or below the median representing low genetic risk for the sleep trait, and the group above the median representing high genetic risk for the sleep trait. Thus, for any combination of two sleep traits, participants were categorized into 4 groups according to their genetic predisposition. For instance, when combining insomnia symptoms and short sleep duration, participants were categorized into: *“Both GRS* ≤ *median”* (representing low genetic risk for both insomnia symptoms and short sleep duration), *“Insomnia GRS > median”* (representing high genetic risk for insomnia symptoms only), *“Short sleep GRS > median*” (representing high genetic risk for short sleep duration only) and *“Both GRS > median”* (representing high genetic risk for both insomnia symptoms and short sleep duration). We then used Cox regression to estimate the causal HRs with 95% CI, adjusted for age at recruitment, sex, assessment centre (in UK Biobank), genetic principal components (40 in UK Biobank and 20 in HUNT2), and genotyping chip. RERI was assessed for the joint causal effect as a test of interaction, as above.^38,39^

To check the proportional hazards assumption, we used Pearson correlations to test Schoenfeld residuals from both observational Cox regression and MR Cox regression for an association with follow-up time.

#### Sensitivity analyses

We performed several sensitivity analyses to test the robustness of our findings:

1. The observational analysis was repeated for a crude model adjusted for only age at recruitment and sex; and an additional model adjusted for variables including those that could be both confounders and mediators, i.e. SBP, serum cholesterol level, blood glucose level, time since last meal, use of sleep medication(s), depression and anxiety.
2. The one-sample MR and 2×2 factorial MR analyses were repeated using uwGRS in HUNT2.
3. To assess the second and third MR assumptions that the genetic instruments used are independent of confounders of the exposure-outcome relation and only affect the outcome through the exposure of interest, associations between the GRS and potential confounders were investigated in UK Biobank and HUNT2 to potentially identify any pleiotropic paths. Furthermore, one-sample MR analysis adjusted for any potential confounders found to be strongly associated with the sleep trait GRS in two cohorts (beyond a Bonferroni significance threshold of P < 5.88×10^−4^ in UK Biobank and P < 7.81×10^−4^ in HUNT2) were explored.
4. To further investigate potential directional pleiotropy, we derived estimates on SNP-exposure and SNP-outcome associations from the same individuals and applied two-sample MR sensitivity analysis methods: inverse variance weighted (IVW), MR-Egger, weighted median, and weighted mode-based methods. These summary-level methods can be used in a one-sample setting,^44^ and the estimates were then compared with the estimates from the individual-level methods. Each of these methods makes different assumptions about the genetic instruments used (see supplementary methods).^45–47^
5. To further assess consistency in effects estimated for insomnia symptoms, 57 SNPs found robustly associated with insomnia by Lane *et al.*^48^ in an older GWAS on UK Biobank (n = 345 022 cases and 108 357 controls) were used in a post hoc one-sample MR Cox regression analysis using same methods as above. These SNPs represent crucial variants for the phenotype any insomnia symptoms (“sometimes”/“usually” as cases versus “never/rarely” as controls).
6. To evaluate the potential impact of winner’s curse, one-sample MR analysis was repeated using genetic variants that replicated at a genome-wide significance level (P < 5×10^−8^) in a large independent dataset for insomnia symptoms (23andMe, n = 944 477, highlighted in supplementary material, Table G1),^35^ and chronotype (23andMe, n = 240D098, highlighted in supplementary material, Table G2).^37^
7. To avoid potential bias due to arbitrary dichotomization and to maximize power, we performed a continuous factorial MR analysis using two GRS (for any combination of two sleep traits) as quantitative traits and their product term.^49^ Further, we calculated RERI as a test of interaction using the risk estimates for the quantitative GRS and their product term for each sleep trait combination where none of the HRs were less than 1 (i.e. preventive) for AF.^38,50^

All analyses were conducted using R version 3.6.3 (R Foundation for Statistical Computing, Vienna, Austria) and performed in line with Strengthening the Reporting of Observational Studies in Epidemiology guidelines for MR studies (STROBE-MR).^51,52^

## Results

In the observational analysis, 16 192 out of 287 352 UK Biobank participants had an AF during a mean (standard deviation (SD)) follow-up of 11.6 (2.1) years, and 3 627 out of 31 458 HUNT2 participants had an AF during a mean (SD) follow-up of 20.7 (6.7) years. Participants with an AF during follow-up in the UK Biobank and HUNT2 were more likely to be males, older, have used sleep medication(s), have higher BMI, systolic blood pressure and blood glucose levels, and be suffering from depression and chronic illness. They were less likely to consume alcohol, be physically active, have a tertiary education, work shifts and be employed compared to participants with no diagnosis of AF (Table 1). The HUNT2 participants with an AF were more likely to have higher serum cholesterol levels, but were less likely to be current smokers and suffering from anxiety when compared to participants with no diagnosis of AF, which is contrary to UK Biobank participants with an AF (Table 1). The baseline characteristics were similar among the study participants included in the MR analysis (see supplementary material, Table S2).

**Table 1:** Baseline characteristics of study participants included in observational analysis with and without an episode of atrial fibrillation (AF) during follow-up in UK Biobank and HUNT2.

|  | UK Biobank (N = 287 352) |  | HUNT2 (N = 31 458) |  |
| --- | --- | --- | --- | --- |
|  | No AF diagnosis | AF diagnosis | No AF diagnosis | AF diagnosis |
| **Total, % (n)** | 94.36 (271 160) | 5.63 (16 192) | 88.47 (27 831) | 11.53 (3 627) |
| **Variables, % (n)** |  |  |  |  |
| Male | 45.55 (123 526) | 64.56 (10 454) | 45.29 (12 604) | 61.54 (2 232) |
| Married | 74.07 (200 856) | 73.88 (11 962) | 59.81 (16 646) | 73.17 (2 654) |
| Weekly alcohol intake | 51.06 (138 465) | 46.93 (7 599) | 26.53 (7 383) | 25.72 (933) |
| Current smokers | 10.22 (27 721) | 10.63 (1 722) | 29.73 (8 273) | 24.21 (878) |
| Highly physically active | 40.61 (110 135) | 40.59 (6 572) | 38.18 (10 625) | 30.30 (1 099) |
| Tertiary education | 46.60 (126 367) | 42.37 (6 861) | 25.38 (7 063) | 18.97 (688) |
| Shift workers | 5.14 (13 936) | 3.59 (581) | 19.09 (5 313) | 9.98 (362) |
| Employed | 60.63 (164 408) | 37.96 (6 146) | 74.23 (20 660) | 51.17 (1 856) |
| Use of sleep medication(s) | 0.87 (2 360) | 1.35 (218) | 5.45 (1 518) | 9.73 (353) |
| Suffering from depression | 11.52 (31 245) | 13.94 (2 257) | - | - |
| Suffering from anxiety | 6.13 (16 630) | 9.20 (1 489) | - | - |
| Suffering from chronic illness | 29.62 (78 965) | 47.63 (7 559) | 26.80 (7 342) | 45.96 (1 636) |
| **Variables, mean (SD)** |  |  |  |  |
| Age, years | 56.09 (8.04) | 61.90 (6.06) | 44.34 (14.78) | 59.94 (12.68) |
| TDI | -1.56 (2.92) | -1.41 (3.02) | - | - |
| BMI, kg/m<sup>2</sup> | 27.17 (4.60) | 28.83 (5.25) | 25.97 (3.93) | 27.52 (4.19) |
| SBP, mmHg | 137.40 (18.44) | 143.40 (19.24) | 133.10 (19.02) | 145.90 (21.65) |
| Serum cholesterol, mmol/L | 5.74 (1.13) | 5.39 (1.18) | 5.70 (1.21) | 6.18 (1.20) |
| Blood glucose, mmol/L | 5.09 (1.16) | 5.31 (1.46) | 5.30 (1.30) | 5.68 (1.62) |
| HADS - D scores | - | - | 3.13 (2.88) | 3.83 (3.01) |
| HADS - A scores | - | - | 4.19 (3.26) | 4.01 (3.24) |

AF, atrial fibrillation; SD, standard deviation; TDI, Townsend deprivation index; BMI, body mass index; SBP, systolic blood pressure; HADS - D scores, Hospital Anxiety and Depression Scale - Depression scores; and HADS - A scores, Hospital Anxiety and Depression Scale - Anxiety scores.

The uwGRS for insomnia symptoms, 24-hour sleep duration, short sleep (≤6 h vs. 7-8 h), long sleep (≥9 h vs. 7-8 h), and morning preference chronotype had F-statistics of 1369.47, 1993.0, 558.61, 270.36, and 5176.33, respectively among UK Biobank participants. The corresponding variance explained (R^2^) was 0.41%, 0.60%, 0.18%, 0.11%, and 1.54% (see supplementary material, Table S3). The wGRS for insomnia symptoms, 24-hour sleep duration, short sleep and long sleep had F-statistics of 72.38, 42.63, 5.35, and 4.42, respectively among HUNT2 participants. The corresponding variance explained (R^2^) was 0.16%, 0.09%, 0.02%, and 0.01% (see supplementary material, Table S3).

The baseline characteristics were evenly distributed among the factorial groups for any two sleep traits (see supplementary material, Table S4-S10), suggesting that the study participants were randomly assigned to groups of similar sized based on their genetic risk for any two sleep traits.

### Individual sleep traits and the risk of AF

Based on the observational analysis, insomnia symptoms were associated with an increased risk of AF in both UK Biobank and HUNT2, with HRs of 1.15 (95% CI 1.06, 1.24) and 1.10 (95% CI 1.00, 1.21), respectively (Figure 2). No association was found for a per hour increase in sleep duration on the risk of AF in UK Biobank and HUNT2, where the corresponding HRs were 0.99 (95% CI 0.96, 1.03) and 0.99 (95% CI 0.96, 1.02). Moreover, short or long sleep duration was not clearly associated with the risk of AF in either cohort. The corresponding HRs in UK Biobank were 1.01 (95% CI 0.93, 1.11) and 1.01 (95% CI 0.88, 1.15), respectively; and in HUNT2 were 0.96 (95% CI 0.53, 1.74) and 1.08 (95% CI 0.99, 1.16), respectively. In addition, morning chronotype was not associated with the risk of AF in UK Biobank (HR 0.95; 95% CI 0.88, 1.02) (Figure 2).

**Figure 2:**
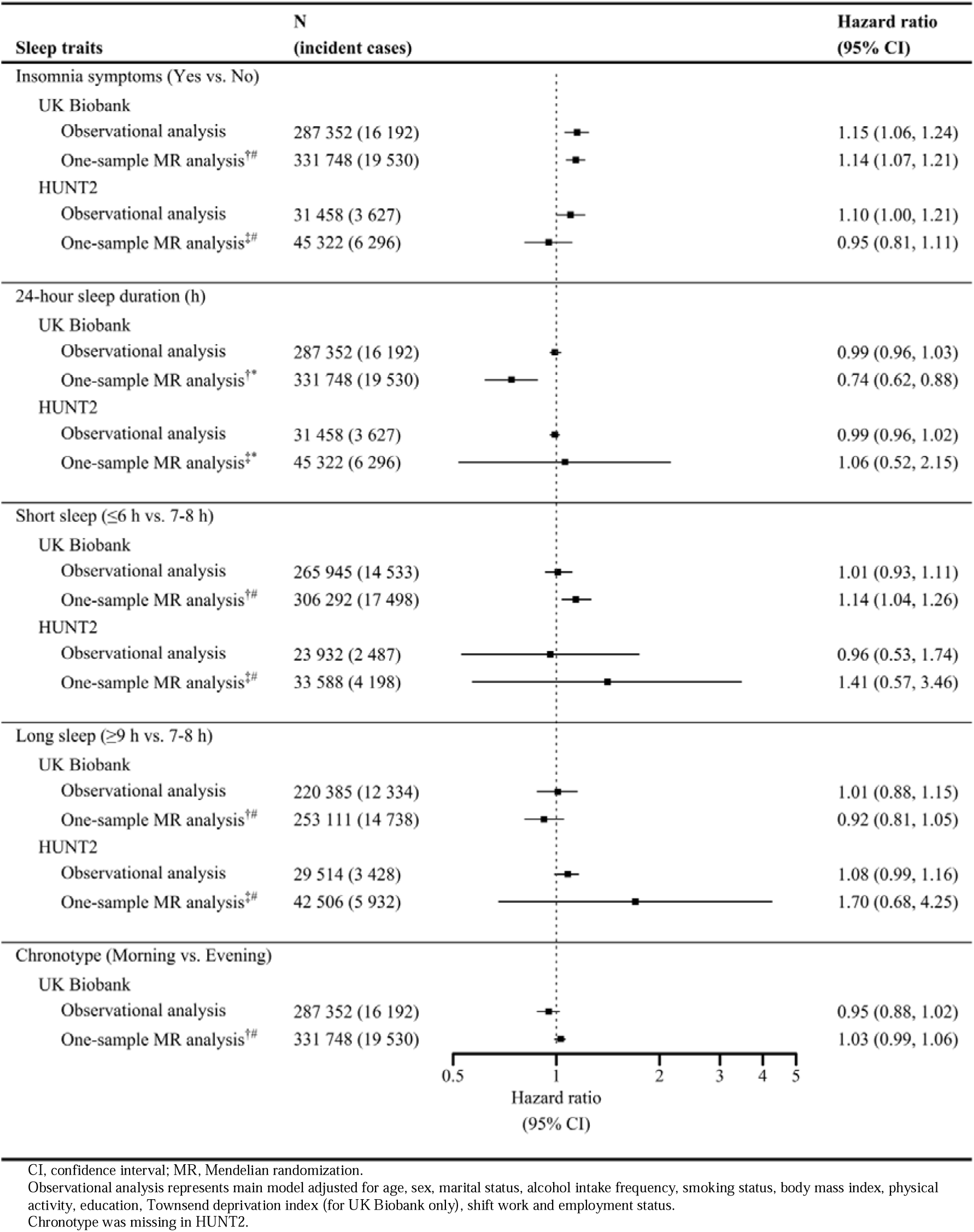
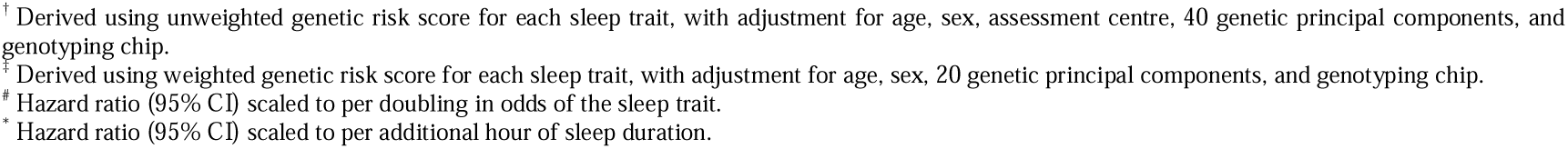
Observational and one-sample Mendelian randomization Cox regression analysis for incident atrial fibrillation in relation to individual self-reported sleep traits in UK Biobank and HUNT2.

Similar to the observational analysis, there was evidence that per doubling in odds of genetically-determined insomnia symptoms increased the incidence of AF in UK Biobank (HR 1.14; 95% CI 1.07, 1.21), however not in HUNT2 (HR 0.95; 95% CI 0.81, 1.11) (Figure 2). Unlike the observational analysis, each hour increase in genetically-determined sleep duration decreased the incidence of AF in UK Biobank (HR 0.74; 95% CI 0.62, 0.88), but not in HUNT2 (HR 1.06; 95% CI 0.52, 2.15). Further, there was evidence that per doubling in odds of genetically-determined short sleep duration increased the incidence of AF in UK Biobank (HR 1.14; 95% CI 1.04, 1.26), however the analysis was underpowered in HUNT2 (HR 1.41; 95% CI 0.57, 3.46). Similar to the observational analysis, genetically-determined long sleep duration was not associated with the incidence of AF in UK Biobank (HR 0.92; 95% CI 0.81, 1.05), and the analysis was underpowered in HUNT2 (HR 1.70; 95% CI 0.68, 4.25). Unlike the observational analysis, there was evidence that per doubling in odds of genetically-determined morning preference chronotype was weakly associated with an increased risk of AF in UK Biobank (HR 1.03; 95% CI 0.99, 1.06) (Figure 2).

### Combination of sleep traits and the risk of AF

#### Insomnia symptoms with short sleep duration

Based on the observational analysis, for UK Biobank participants who reported short sleep duration without insomnia symptoms, there was little evidence of a decreased risk of AF (HR 0.98; 95% CI 0.86, 1.11), while for those who reported short sleep duration with insomnia symptoms, there was weak evidence of an increased risk (HR 1.10; 95% CI 0.98, 1.25), and for those who reported normal sleep duration with insomnia symptoms, there was strong evidence of an increased risk of AF (HR 1.14; 95% CI 1.03, 1.26) when compared to those who reported normal sleep duration without insomnia symptoms (Figure 3). There was a similar pattern in HUNT2, however the corresponding HRs were imprecise (Figure 3).

**Figure 3:**
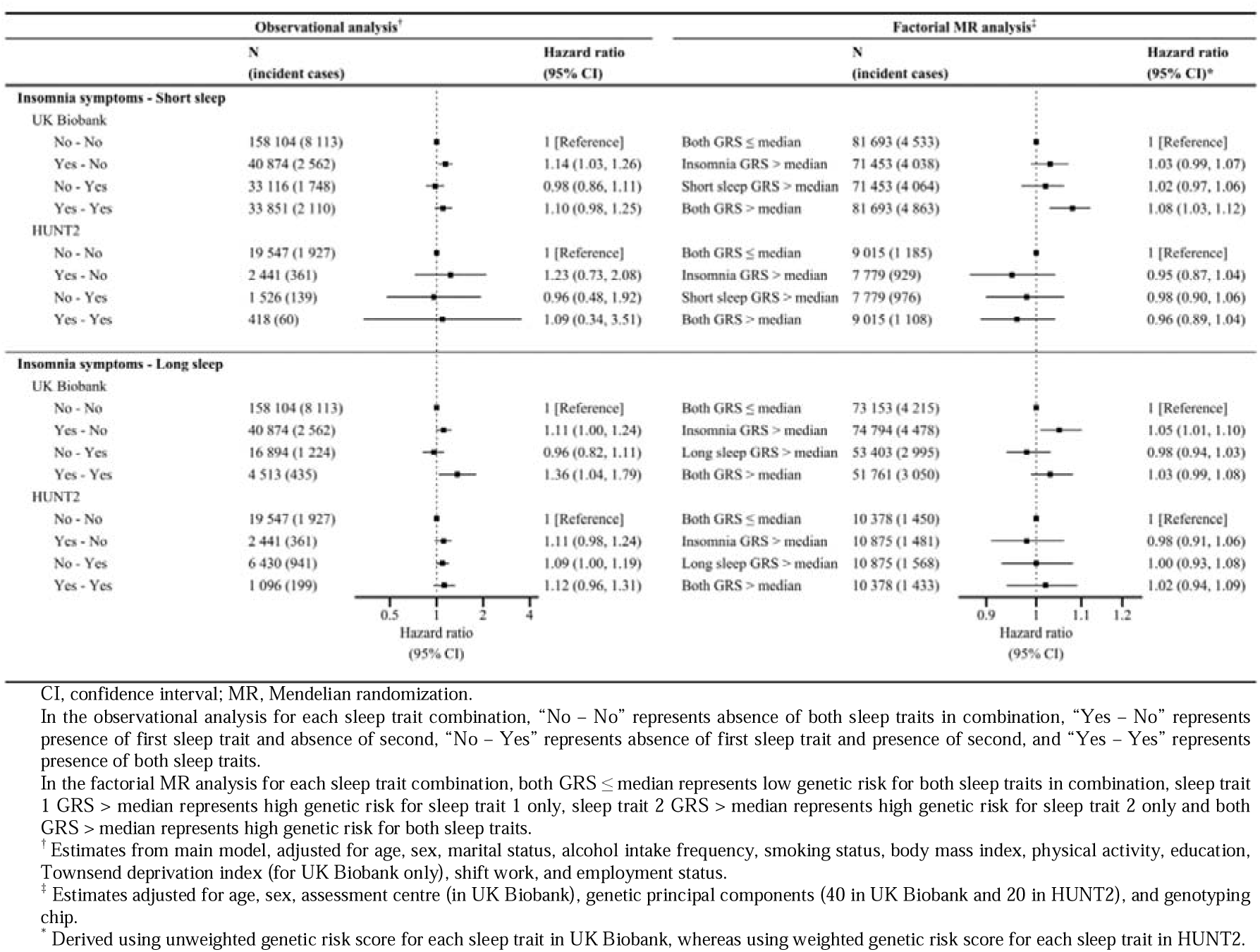
Observational and 2×2 factorial Mendelian randomization Cox regression analysis assessing the joint effects of insomnia symptoms and sleep duration on the risk of atrial fibrillation in UK Biobank and HUNT2.

In UK Biobank, participants with only a high genetic risk for insomnia symptoms or high genetic risk for short sleep duration had a slightly higher risk of AF (HR 1.03; 95% CI 0.99, 1.07 and HR 1.02; 95% CI 0.97, 1.06, respectively), whereas participants with high genetic risks for both traits had the highest risk (HR 1.08; 95% CI 1.03, 1.12) (Figure 3); however, there was no evidence of interaction (RERI 0.03; 95% CI −0.03, 0.09). This pattern was not consistent in HUNT2, showing imprecise estimates (Figure 3).

#### Insomnia symptoms with long sleep duration

Based on the observational analysis, for UK Biobank participants who reported long sleep duration without insomnia symptoms, there was little evidence of a decreased risk of AF (HR 0.96; 95% CI 0.82, 1.11), while for those who reported long sleep duration with insomnia symptoms, there was strong evidence of an increased risk (HR 1.36; 95% CI 1.04, 1.79) and for those who reported normal sleep duration with insomnia symptoms, there was slightly weaker evidence of an increased risk of AF (HR 1.11; 95% CI 1.00, 1.24) when compared to those who reported normal sleep duration without insomnia symptoms (Figure 3). However, the joint association of insomnia symptoms and long sleep duration on the risk of AF was inconclusive in HUNT2 (Figure 3).

The factorial MR analysis of genetic risks for insomnia symptoms and long sleep duration on risk of AF, was inconclusive in both UK Biobank and HUNT2 (Figure 3).

#### Chronotype with insomnia symptoms or short/long sleep duration

Based on the observational analysis, for UK Biobank participants who reported morning chronotype without insomnia symptoms, there was little evidence of a decreased risk of AF (HR 0.96; 95% CI 0.88, 1.05), while for those who reported morning chronotype with insomnia symptoms, there was weak evidence of an increased risk (HR 1.08; 95% CI 0.97, 1.21) and for those who reported evening chronotype with insomnia symptoms, there was strong evidence of an increased risk of AF (HR 1.18; 95% CI 1.04, 1.35) when compared to those who reported evening chronotype without insomnia symptoms (Figure 4). However, the joint associations of morning chronotype and short sleep duration, as well as morning chronotype and long sleep duration on the risk of AF were inconclusive in UK Biobank (Figure 4).

**Figure 4:**
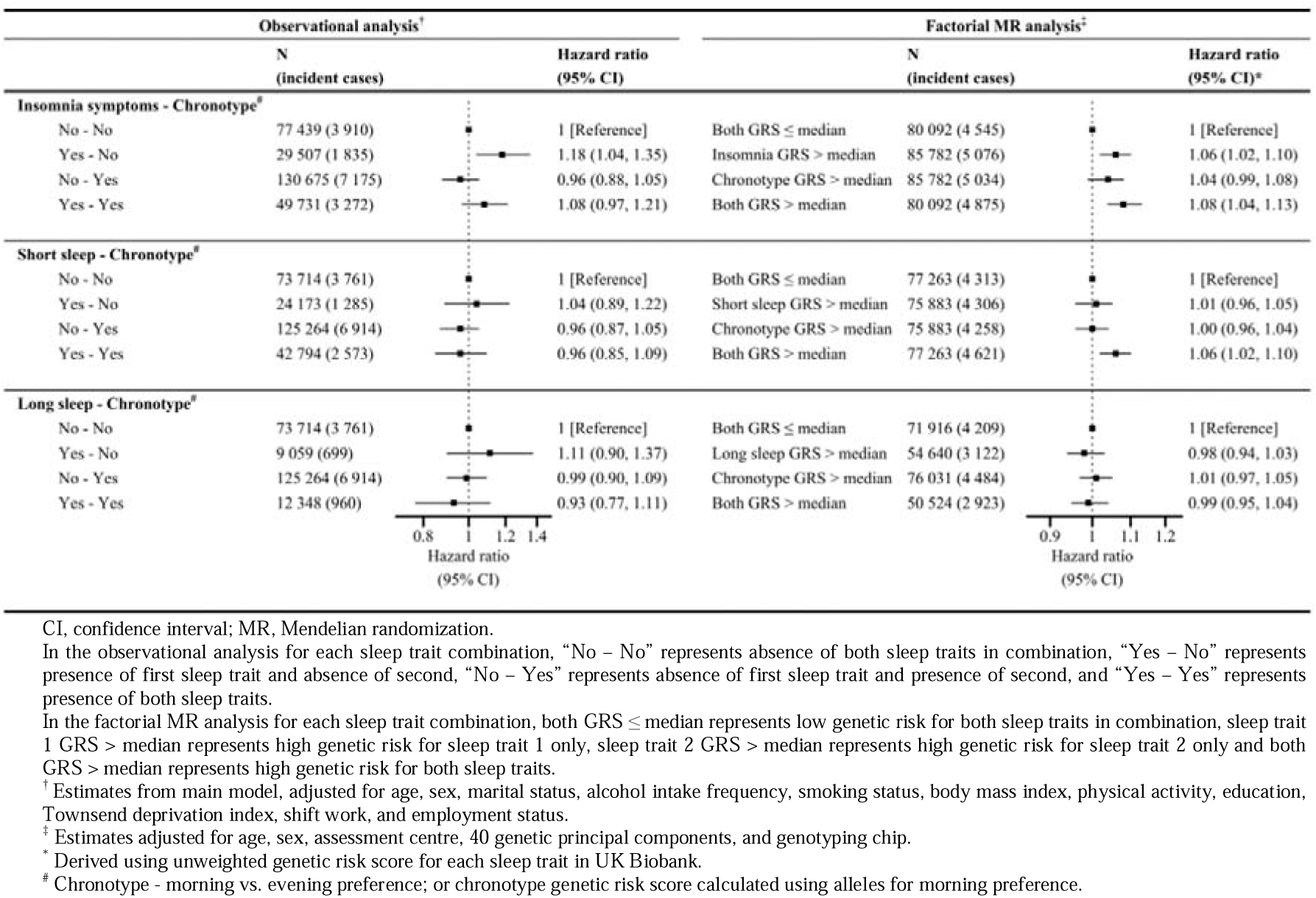
Observational and 2×2 factorial Mendelian randomization Cox regression analysis assessing the joint effects of insomnia symptoms/sleep duration and chronotype on the risk of incident atrial fibrillation in UK Biobank.

In addition, UK Biobank participants with only a high genetic risk for insomnia symptoms or high genetic risk for a morning chronotype had a slightly higher risk of AF (HR 1.06; 95% CI 1.02, 1.10 and HR 1.04; 95% CI 0.99, 1.08, respectively), whereas participants with high genetic risk for both sleep traits had the highest risk (HR 1.08; 95% CI 1.04, 1.13) (Figure 4); however, there was no evidence of interaction (RERI −0.01; 95% CI −0.07, 0.04). The UK Biobank participants with only a high genetic risk for short sleep duration or high genetic risk for a morning chronotype had no increased risk of AF (HR 1.01; 95% CI 0.96, 1.05 and HR 1.00; 95% CI 0.96, 1.04, respectively), whereas participants with high genetic risk for both sleep traits had an increased risk (HR 1.06; 95% CI 1.02, 1.10), although again there was no statistical evidence of interaction (RERI 0.06; 95% CI −0.01, 0.12). The factorial MR analysis of genetic risks for long sleep duration and morning chronotype on risk of AF in UK Biobank was inconclusive (Figure 4).

The proportionality of hazards assumption was held for all the observational and MR Cox regression analyses (see supplementary material, Tables S11-S12).

### Sensitivity analysis

1. Further adjustment of potential confounding variables did not change the results markedly when repeating the observational analysis with additional potential confounders (see supplementary material, Tables S13-S19).
2. Using the uwGRS for the sleep traits in HUNT2, the one-sample MR estimates changed markedly for short sleep duration (HR 1.97; 95% CI 0.68, 5.72 versus HR 1.41; 95% CI 0.57, 3.46 for wGRS) and long sleep duration (HR 1.19; 95% CI 0.64, 2.23 versus HR 1.70; 95% CI 0.68, 4.25 for wGRS), but only slightly for insomnia symptoms and 24-hour sleep duration (see supplementary material, Table S20). However, the 2×2 factorial MR estimates remained unchanged (see supplementary material, Figure S1).
3. Even after correction for multiple testing, several confounding factors were associated with the sleep trait GRS in both UK Biobank and HUNT2 which might indicate potential pleiotropy (see supplementary material, Tables S21 & S22). Further sensitivity analysis was undertaken adjusting for these potential confounders in the one-sample MR analysis, and the effect estimates for insomnia symptoms (HR 1.05; 95% CI 0.97, 1.13) and short sleep duration (HR 1.09; 95% CI 0.98, 1.22) attenuated slightly and were less precise. Moreover, adjusting for potential confounders, the effect estimate for 24-hour sleep duration attenuated slightly in UK Biobank (HR 0.82; 95% CI 0.67, 1.00) (see supplementary material, Table S23).
4. The causal estimates obtained using MR-Egger, weighted median- and weighted mode-based methods attenuated slightly and were less precise (see supplementary material, Figures S2-S6, Tables S24 & S25).
5. Further, the post hoc one-sample MR analysis using insomnia symptoms variants from Lane *et al.*^48^ gave similar results, where the TSPS estimates from UK Biobank and HUNT2 showed a suggestive adverse causal effect (HR 1.08; 95% CI 0.99, 1.18 and HR 1.11; 95% CI 0.87, 1.41, respectively) (see supplementary material, Figure S7 and Table S26).
6. The causal estimates attenuated only slightly when using the GRS consisting of 116 insomnia SNPs (one missing in the HUNT imputed dataset) and 72 chronotype SNPs which replicated at a genome-wide significance level (P < 5×10^−8^) in the independent 23andMe dataset (see supplementary material, Tables S27 & S28).
7. The estimates from the continuous factorial MR analysis using sleep trait GRS as quantitative traits (per standard deviation increase) and their product term inferred similar effects (see supplementary material, Figure S8). In UK Biobank, the GRS for insomnia symptoms and short sleep duration were associated with an increased risk of AF (HR 1.03; 95% CI 1.01, 1.04 and HR 1.02; 95% CI 1.00, 1.03, respectively), but there was no evidence of interaction (RERI 0.01; 95% CI −0.01, 0.02). Similarly, the GRS for insomnia symptoms and morning chronotype were associated with an increased risk of AF (HR 1.03; 95% CI 1.02, 1.05 and HR 1.01; 95% CI 1.00, 1.03, respectively), though there was no evidence of interaction (RERI −0.01; 95% CI −0.02, 0.01). Also, the GRS for short sleep duration and morning chronotype were associated with an increased risk of AF (HR 1.02; 95% CI 1.00, 1.04 and HR 1.01; 95% CI 1.00, 1.03, respectively), with no evidence of interaction (RERI 0.01; 95% CI −0.01, 0.02).

The STROBE-MR checklist was presented in the supplementary material (Table S29).

## Discussion

In the observational analysis, we found insomnia symptoms were associated with an increased risk of AF, whereas there was limited evidence of association for sleep duration (including short or long sleep) and chronotype in both cohorts. In the one-sample MR analysis, we found evidence of adverse effects of insomnia symptoms and short sleep duration, and a protective effect of sleep duration on the risk of AF in UK Biobank. However, these associations were either not supported or too imprecise in HUNT2 for validation, likely due to low power in this cohort.

There was no evidence of relative excess risk due to interaction, suggesting the joint effect of two sleep traits on AF does not exceed the effect of each sleep trait assessed independently. Although in the factorial MR analysis participants with high genetic risks for two sleep traits in certain combinations (i.e., insomnia symptoms with short sleep duration, insomnia symptoms with morning chronotype, and short sleep duration with morning chronotype) had higher risks of AF than those with high genetic risk for a single sleep trait, but in UK Biobank only. These results were not replicated in HUNT2, where the estimates were imprecise.

### Comparison of observational and MR findings across other studies

Our findings of an adverse effect of insomnia symptoms on risk of AF in all analyses performed in UK Biobank are consistent across much of the previous observational^10,53^ and MR studies.^23–25,54^ However, in HUNT2, our observational analysis found weak evidence of an increased risk of AF, whereas our one-sample MR analysis showed no effect of insomnia symptoms on the risk of AF. A recent observational study conducted using the third survey of the Trøndelag Health Study (HUNT3; n = 33 983) found no association between insomnia symptoms and the risk of AF.^55^ This lack of replication in HUNT2 and HUNT3 could be due to low statistical power from its smaller sample, where larger study sizes might have shown some associations. Moreover, the estimated effect of insomnia symptoms using fewer SNPs from Lane *et al.*^48^ showed the tendency towards an increased risk of AF in participants genetically predisposed to insomnia symptoms (HR 1.11; 95% CI 0.87, 1.41) in HUNT2.

Our findings of no association of 24-hour sleep duration on the risk of AF from observational analysis in both cohorts could be due to underlying U-shaped association, as evident from previous observational studies noting that both short and long sleep durations increase the risk of AF.^11,53,56–58^ However, surprisingly, we found no associations of short (≤6 h vs. 7-8 h) or long (≥9 h vs. 7-8 h) sleep durations when assessed separately in the observational analysis. On the other hand, one-sample MR findings in UK Biobank suggested a protective effect per hour increase in sleep duration and an adverse effect of short sleep duration on the risk of AF, which are in line with previous MR analyses.^26,27^ However, these findings were not replicated in HUNT2, which again may be due to smaller sample and/or weak instrument for short sleep duration.

Our observational findings of no association of chronotype on the risk of AF in the UK Biobank align with prior research.^53^ Also, our MR findings indicate no causal effect of chronotype on the risk of AF in UK Biobank.

Overall, there was no statistical evidence of interaction among different combinations of sleep traits, suggesting the joint effect of sleep traits on AF does not exceed the effect of each sleep trait assessed independently (i.e., no evidence of synergism). Although a prior observational study in UK Biobank found healthy sleep patterns (represented by morning chronotype, adequate sleep duration (7–8 h), no frequent insomnia symptoms, no snoring, and no frequent daytime sleepiness) were associated with a lower risk of AF.^53^ Interestingly, in our factorial MR analysis participants with high genetic risks for two sleep traits in certain combinations (i.e., insomnia symptoms with short sleep duration, insomnia symptoms with morning chronotype, and short sleep duration with morning chronotype) had higher risks of AF than those with high genetic risk for a single sleep trait in UK Biobank. However, this was not replicated in HUNT2, possibly due to low power from a smaller sample and/or weak instruments for short and long sleep durations.

### Potential mechanisms

The definitive mechanisms mediating detrimental effects of sleep disturbances on AF risk remain elusive. Nonetheless, it is postulated that insomnia symptoms and short sleep duration trigger a range of physiological processes, including autonomic dysregulation,^59–61^ activation of proinflammatory and oxidative stress pathways,^62^ dysregulation of hypothalamic-pituitary axis,^63^ and activation of renin-aldosterone-angiotensin system.^64^ These processes may contribute to atrial remodelling and fibrosis,^65–67^ resulting in the loss of atrial muscle mass and subsequent promotion of proarrhythmic conditions conducive to AF incidence. Given there was no evidence of relative excess risk due to interaction, we do not suspect these potential mechanisms to interplay with each other.

### Strengths and limitations

Our study is the first to explore the role of individual sleep traits and their joint effects on the incidence of AF using multiple approaches in two large cohorts – the UK Biobank and the HUNT study. certificates which reduced bias due to misclassification. Furthermore, this study benefitted from the MR analyses that use SNPs from three large GWASs for insomnia symptoms,^35^ sleep duration^36^ and chronotype.^37^

The present study employs various approaches that exhibit distinct advantages and disadvantages. In observational analyses, efforts were made to alleviate possible sources of bias, such as confounding and reverse causation, by utilizing multivariable Cox regression of incident AF and controlling for several potential confounders. However, to further minimize the likelihood of bias due to confounding, reverse causation and measurement error, we applied MR.

However, our study has several limitations. Self-reported measures of sleep traits were used in observational analyses, as well as to identify genetic variants for subsequent MR analysis. It remains unclear if self-reported sleep duration captures time in bed or actual sleep time. The insomnia questions used in both cohorts did not cover all components used in the framework for diagnosing insomnia.^31^ Our use of a simplified definition of insomnia considers people with some-to-severe insomnia in this study, which may underestimate our findings where previous studies indicate that the consequences of insomnia are related to insomnia severity.^68,69^ Chronotype was assessed from a single question about chronotype in UK Biobank, rather than considering validated tools.^70,71^ Also, the biological functions of the genetic variants employed in MR analysis to predict sleep traits, and the mechanistic pathways underlying their observed effects remain poorly understood.

We used sleep traits as binary exposures (except 24-hour sleep duration), which are likely coarsened approximations of the true latent exposure.^72^ This deliberate loss of information may increase the likelihood of false positive findings, but also opens up alternate pathways from the genetic instruments to the outcome, which may violate the exclusion restriction assumption of MR and result in biased effect estimates.^72^ Our investigations may have been impelled to identify any non-linear effects of the exposures due to their binary nature. However, we also acknowledge that application of non-linear MR methods is underdevelopment.^73,74^

The selection of participants may be a source of bias and may limit generalizability. We relied on our findings from UK Biobank due to its large sample, although it is not representative of the UK population owing to low participation (5.5% response rate),^75^ and may have led to bias in both the observational and MR estimates of association through selection bias.^76–79^ On the other hand, HUNT2 (69.5% response rate) is more representative of the general population and may be subject to less bias, which could explain why the estimates for the UK Biobank and HUNT2 differed. Moreover, the study sample restricting to participants from European ancestry in both cohorts may further limit generalizability of our findings to other ethnicities.

The limited sample size of the HUNT2 study may have resulted in inadequate statistical power, leading to the potential oversight of weak evidence of an effect. Moreover, the genetic instruments employed explained much less variance in the sleep traits, particularly for short and long sleep durations within the HUNT2 sample, raising concerns about weak instrument bias.^80^ A simulation study has shown that use of bootstrapping to obtain corrected standard errors is limited to the availability of strong instruments.^81^ Thus, we restricted the use of bootstrapping in our MR analyses owing to weak instruments for short and long sleep durations in HUNT2. Furthermore, the SNPs used to construct the GRS for short and long sleep durations were not assessed for replication in other independent cohorts,^36^ implying that the validity of the GRS for short and long sleep durations utilized in this study is not established in any other population.

Despite our considerable efforts to limit potential confounding in the observational analysis, we recognize that residual confounding due to measurement errors or misclassification of the confounders, as well as confounding due to factors not considered/collected still persists.^82^ An important example of such factor is obstructive sleep apnoea (OSA), which is a well-established risk factor for AF.^83^ We did not have information on OSA or other sleep disorders in our study. OSA changes intrathoracic pressure, leading to cyclic augmentation of atrial wall stress. This stress can further exacerbate autonomic dysregulation and inflammation pathways,^84^ which are potentially pathophysiological in the development of AF. For OSA to be able to substantially influence our estimates, it must be unrelated to other covariates controlled for in the models, while also being strongly associated with both the exposure and the outcome. OSA is known to be correlated with age, BMI, blood pressure and depression.^85,86^ By adjusting for these related variables in our observational analysis, we have partially accounted for some of the confounding that could arise from OSA. Thus, it seems unlikely that OSA alone could explain increased risk of AF observed among participants with insomnia symptoms and short sleep duration. Moreover, OSA is both underdiagnosed and under-recognized.^87^

In all MR analysis, we could not exclude the possibility of pleiotropy.^88^ However, we applied several robust methods (such as MR-Egger, weighted median and weighted mode based methods) as sensitivity analyses to test for horizontal pleiotropy.^44^ Although these methods produce less precise estimates, consistent results in the same direction strengthens confidence in identification of causal links. We also found that genetic risk for insomnia symptoms was strongly associated with BMI, smoking status, depression, and education among other covariates, which may be indicative of confounding, mediation or horizontal pleiotropy.^35^ However, our results remained fairly consistent across various MR robust methods. Additionally, previous studies have showed only mild attenuation of causal effects of insomnia symptoms on AF risk when adjusted for BMI, smoking, depression and education using multivariable Mendelian randomization.^23,24^

The SNP-exposure estimates were obtained from three GWASs on sleep traits, where UK Biobank formed part of the discovery cohort. This could lead to winner’s curse, when the magnitude of the effect sizes for genetic variants identified within a discovery cohort is likely to be exaggerated.^89^ In a one-sample MR analysis, the impact of winner’s curse of the SNP-exposure association can bias estimates towards the confounded observational estimate.^89^ We therefore used unweighted GRS for our sleep traits in UK Biobank to minimize any bias of winner’s curse.^40^ To further check the impact of winner’s curse in one-sample MR, we derived GRS for insomnia symptoms and chronotype composed of SNPs that replicated in an independent sample (i.e., 23andMe).^35,37^ However, we could not apply the same approach to check the impact of winner’s curse on sleep duration due to the limited sample size of the replication dataset, meaning that genetic associations might be imprecise.^36^ Moreover, we acknowledge availability of information from latest GWAS meta-analysis of insomnia conducted on 593 724Dcases and 1 771 286Dcontrols, including data from the UK Biobank and 23andMe.^90^ This analysis identified 554 risk loci, with 364 being novel for insomnia. Nonetheless, our study relied on individual-level genetic data to compute the GRS of insomnia, where SNP information was limited.

Although factorial MR is a promising approach for investigating the joint effects of multiple exposures on a particular outcome, one of the main concern is the underpowered nature of this approach, which can lead to false negative results.^49^ Nevertheless, this study addressed this limitation by utilizing the UK Biobank cohort, consisting of 331 748 participants, representing the largest factorial MR study conducted to date on sleep traits. Moreover, the good instrument strength observed in UK Biobank cohort further mitigate concerns due to underpowered factorial MR findings.^91^ However, this may also be a limitation due to sample overlap with the discovery cohort. Additionally, it is important to acknowledge that MR assumes that the exposures remain stable throughout an individual’s life course,^22^ and as such, any observed joint effects should be interpreted with caution. While factorial MR can effectively identify if two independent exposures interact and have a joint effect that holds public health significance,^91^ it is critical to consider the potential variations in exposure over time and how these fluctuations might affect the magnitude of observed effects.

Furthermore, direct comparison of the findings from observational and MR study designs must be done cautiously, as disparities in results may stem from methodological differences. MR estimates the effect of small differences in lifelong exposures, which might not be directly comparable to the effects of self-reported exposure differences over limited periods in conventional observational study designs. Also, it is crucial to acknowledge that potential interaction effects could be apparent only at certain thresholds. In our study, the use of sleep traits as binary exposures, we overlook whether the differences in sleep traits among groups in the 2×2 factorial analysis (dichotomized across median GRS cut-offs) correspond to an important distinction in exposures or if the differences between groups are too small to detect interaction effects.

## Conclusions

This study reveals that individuals with two sleep traits do not have a risk of AF that exceeds the additive risk from the individual sleep traits (i.e., no relative excess risk due to interaction), indicating the effect of each sleep traits on AF are likely to be independent of each other. Using both observational and MR study design this study also provides strong evidence for an adverse causal effect of insomnia symptoms on the incidence of AF. There is some evidence of an adverse causal effect of short sleep duration and a protective causal effect of an additional hour of sleep duration on the incidence of AF. Our findings support effective management of insomnia symptoms and short sleep duration in general population to mitigate the risk of AF and improve overall cardiovascular health.

## Data Availability

The data supporting the findings are available in the supplementary material and upon request. The UK Biobank data is available to researchers, subject to successful registration and application process via their website (https://www.ukbiobank.ac.uk/). The data from the HUNT Study are available from the HUNT Research Centre but restrictions apply to the availability of these data, which were used under license for the current study, and so are not publicly available. However, the data are available for export given approval of application to the HUNT Research Centre (http://www.ntnu.edu/hunt/data). The data on hospital records linkages to the HUNT Study participants are available from Nord-Trøndelag Hospital Trust and require permission. All other data used are publicly available and referenced according in the main text.

http://www.ntnu.edu/hunt/data

## Acknowledgements

This research has been conducted using data from UK Biobank, a major biomedical database (https://www.ukbiobank.ac.uk/), under application number 40135.

The Trøndelag Health Study (HUNT) is a collaboration between HUNT Research Centre (Faculty of Medicine and Health Sciences, Norwegian University of Science and Technology (NTNU)), Trøndelag County Council, Central Norway Regional Health Authority, and the Norwegian Institute of Public Health. We want to thank clinicians and other employees at Nord-Trøndelag Hospital Trust for their support and for contributing to data collection in this research project.

## Funding statement

This study was made possible with the financial support from the Norwegian Health Association (*Norwegian*: Nasjonalforeningen for folkehelsen; project number 19479), mobility grant funds from the Liaison Committee for Education, Research and Innovation in Central Norway (*Norwegian*: Samarbeidsorganet mellom NTNU og Helse Midt-Norge; project number 2023/34249) and top-up financing from the Department of Public Health and Nursing, Norwegian University of Science and Technology, Trondheim, Norway. The funders had no role in study design, data collection and analysis, decision to publish, or preparation of the manuscript.

Laxmi Bhatta received support from the Liaison Committee for Education, Research and Innovation in Central Norway (*Norwegian*: Samarbeidsorganet mellom NTNU og Helse Midt-Norge); and the Joint Research Committee between St. Olavs Hospital and the Faculty of Medicine and Health Sciences, NTNU.

## Disclosure of interest

The authors declare that there is no conflict of interest. The authors of this manuscript have certified that they comply with the principles of ethical publishing.

## Ethical approval and consent

UK Biobank received ethical approval from the National Health Service (NHS) Research Ethics Service (reference number 11/NW/0382). The HUNT Study was approved by the Data Inspectorate of Norway and recommended by the Regional Committee for Ethics in Medical Research (REK; reference number 152/95/AH/JGE). The ethical approval for conducting this study was also obtained from the Regional Committee for Ethics in Medical Research (REK nord; reference number 2020/47206).

Informed consent was obtained from all individual participants of both the cohorts included in this study.

## CRediT statement

Conceptualization: BMB, RCR, ESS, LBS, BOÅ; Data curation: NA, JPL, VM, RCR, LBS; Formal analysis: NA; Funding acquisition: NA, LBS; Methodology: NA, BMB, LB, RCR, LBS; Project administration: NA, LBS; Resources: BMB, RCR, LBS; Supervision: LB, RCR, ESS, LBS, BOÅ; Visualization: NA, BMB, RCR, LBS; Writing – original draft: NA; and Writing – review and editing: BMB, LB, JPL, VM, RCR, ESS, LBS, BOÅ.

## Notes

### Competing Interest Statement

The authors have declared no competing interest.

### Author Declarations

UK Biobank received ethical approval from the National Health Service (NHS) Research Ethics Service (reference number 11/NW/0382). The HUNT Study was approved by the Data Inspectorate of Norway and recommended by the Regional Committee for Ethics in Medical Research (REK; reference number 152/95/AH/JGE). The ethical approval for conducting this study was also obtained from the Regional Committee for Ethics in Medical Research (REK nord; reference number 2020/47206). Informed consent was obtained from all individual participants of both the cohorts included in this study.

### Summary of Updates

Manuscript revised - Some few revisions in introduction and discussion sections. Supplemental files updated.

