## Supplementary material for "A Mendelian randomization study investigating the role of sleep traits and their joint effects on the incidence of atrial fibrillation"

### Supplementary methods

### Detailed information on covariates

Information on several covariates as potential confounding factors such as sex, age at recruitment, marital status, alcohol intake, smoking status, physical activity, Townsend Deprivation Index (in UK Biobank only), education, shift work, and use of sleep medication(s) were collected through a self-administered questionnaire. Additionally, participants underwent clinical examinations and trained staff drew blood samples at examination stations.

In UK Biobank, participants were categorized as “married” if they live with their husband/wife/partner, and as “unmarried” if they don’t. In cases where the marital status information was unavailable, participants living alone were classified as “unmarried” based on the number of people living in their household. In HUNT2, participants were categorized as “married”, “unmarried”, and “separated/divorced/widowed”.

Both UK Biobank and HUNT2 participants were asked their alcohol intake frequency and were categorized as “never/rarely” for non-drinkers or those who only drink on special occasions, “monthly” for those who drink 1-3 times a month, “weekly” for those who drink 1-4 times a week, or “daily/almost daily” for those who drink more frequently. In the HUNT2, the information about participants who had never consumed alcohol was used to categorize them as “never/rarely” for observations having missing information on alcohol intake frequency. Hence, the recorded information was grouped as “never/rarely,” “monthly,” “weekly,” or “daily/almost daily” alcohol intake.

Both UK Biobank and HUNT2 participants were categorized as “never”, “previous” or “current” smoker for their smoking status.

In UK Biobank, physical activity (PA) was assessed using adapted questions from the validated short International Physical Activity Questionnaire (IPAQ),^1^ following the data processing guidelines published by IPAQ.^2^ Total physical activity, comprising walking, moderate, and vigorous PA over the previous 7 days was evaluated. Participants were classified into three mutually exclusive PA categories as performing “high PA” (≥ 1 hour of moderate PA or ≥ ½ hour of vigorous PA above the basal activity level on most days), “moderate PA” (≥ ½ hour of moderate PA above the basal activity level on most days), or “low PA/inactive” (anything else) using standard scoring criteria.^3^ The basal activity level was considered to be approximately 5000 steps per day. In HUNT2, PA was assessed based on self-reported leisure time light and hard PA in the past year. Light PA was defined as activity that did not cause sweating or shortness of breath, while hard PA was defined as activity that resulted in sweating or shortness of breath, including the commute to work as leisure time. The study participants were grouped into three mutually exclusive categories as performing “high PA” (defined as ≥ 1 hour of hard PA regardless of light PA or ≥ 3 hours of light PA with < 1 hour of hard PA), “moderate PA” (defined as ≥ 3 hours of light PA with no hard PA or < 3 hours of light PA with < 1 hour of hard PA), or “low PA/inactive” (for anything else). This categorization strategy for PA was previously used by Brumpton *et al.*^4^ The questions on PA from HUNT2 were reported to have acceptable reliability and validity for hard PA but poor for light PA.^5^

In UK Biobank, the Townsend Deprivation Index (TDI) was utilized to determine the socioeconomic status of participants. This index was derived from census data on housing, employment, car availability and social class based on postal codes of participants. Higher TDI values correspond to greater levels of deprivation. The validity of the TDI for use in a UK population has been established through validation studies.^6^

Both UK Biobank and HUNT2 participants were asked about their education attainment and were categorized as “10 years or less” (for primary and lower secondary school education), “11-13 years” (for upper secondary school education), or “14 years or more” (for university/college education).

The UK Biobank questionnaire contained separate questions about working night shifts or shift work, which were then combined to create a proxy variable and the highest response category was used as final. The resulting proxy variable was then dichotomized, where “usually” or “always” were classified as “yes” and all other responses as “no”. In HUNT2, information on working shifts/working at night/being on call was also dichotomized as “yes” or “no”. Additionally, in both UK Biobank and HUNT2, current employment/work status was used to categorize participants without paid employment or those who were self-employed as “no” for observations with missing information on working shifts/at night/on call.

In UK Biobank, participants’ use of sleep medications were ascertained by self-report. This was based on a list of sleep medication as used in a previous study by Daghlas *et al.*,^7^ along with five additional commonly used anxiolytics or sleep medications (Table S30). The responses were then categorized as “yes” indicating the use of sleep medication(s), and “no” indicating the absence of use. In HUNT2, participants were asked about their use of anxiolytics or sleep medications within the last month and categorized as “yes” if they reported using them daily or weekly, and “no” if they reported otherwise.

#### Clinical examination

In UK Biobank, weight was measured using the Tanita BC-418MA body composition analyzer to the nearest 0.1kg, while height was measured with a Seca 202 height measure. Meanwhile, in HUNT2, weight was measured to the nearest 0.5kg, and height was measured to the nearest 1cm. In both cohorts, participants were instructed to wear light clothing and no shoes during these measurements. The body mass index (BMI) was then calculated by dividing the weight (in kg) by the square of the height (in meter).

Blood pressure measurements were obtained using different methods and protocols in the UK Biobank and the HUNT2. In UK Biobank, automated (using Omron HEM-705 IT electronic blood pressure monitor) and/or manual (using manual sphygmomanometer) measurements were taken for systolic and diastolic blood pressure, with two sets of measurements taken one minute apart, and the average of these two was used in the analysis. In cases where automated readings were unavailable, manual readings were used instead. In HUNT2, systolic and diastolic measurements of blood pressure were recorded automatedly (using a Dinamap 845XT (Critikon) sphygmomanometer based on oscillometry), with three sets of measurements were taken one minute apart, and the average of the second and third measurements used in the analysis.

#### Laboratory measurements

In accordance with the standard operating procedures for the UK Biobank, a random (non-fasting) blood sample was collected from each participant and stored in refrigerators at temperatures ranging from 2 to 8°C. Fasting time was noted as the duration between the last consumption of food or drink and the blood sample collection. The samples were transported to a central laboratory for storage and analysis on a daily basis. Serum samples were centrifuged at 2000 RCF for 10 minutes, and the concentrations of glucose, total cholesterol, HDL-cholesterol, and triglycerides were determined using a Beckman Coulter AU5800 automated analyzer. Glucose was measured using hexokinase analysis, while total cholesterol, HDL-cholesterol and triglycerides were measured by CHO-POD analysis, enzyme immunoinhibition analysis and GPO-POD analysis, respectively.^8^

In HUNT2, a random (non-fasting) blood sample was collected from each participant. Within two hours of collection, the serum was separated from the blood by centrifugation and stored in a refrigerator at 4°C at the screening site. Time between the last meal and venipuncture was recorded. The samples were then sent to the central laboratory at Levanger Hospital, where they were analyzed using a Hitachi 911 Autoanalyzer (Hitachi, Mito, Japan). The samples were transported to the laboratory on the same day or within two to three days (for example on weekends). The serum concentrations of glucose, total cholesterol, HDL-cholesterol, and triglycerides were analyzed using reagents from Boehringer Mannheim (Mannheim, Germany). The day-to-day coefficients of variation were 1.3-2.0%, 1.3-1.9%, 2.4%, and 0.7-1.3%, respectively. Glucose was measured using an enzymatic hexokinase method, total cholesterol and HDL-cholesterol were measured using an enzymatic colorimetric cholesterol esterase method, and triglycerides were measured using an enzymatic colorimetric method.^9^

#### Depression and anxiety

Anxiety and depression episodes in UK Biobank were identified from hospital records using ICD-10 codes - F40 and F41 for anxiety, and F32, F33, F34, F38, and F39 for depression. This information was then used to create two binary proxy variables for each for anxiety and depression, categorized as "yes" or "no".

In HUNT2, the symptoms of anxiety and depression were evaluated using the Hospital Anxiety and Depression Scale (HADS). The questionnaire consisted of 14 Likert-scaled items (7 each for anxiety and depression) having a four-point scale ranging from 0 (not at all) to 3 (very often). The responses were summed to generate anxiety and depression scores ranging from 0 to 21, with higher scores indicating a greater likelihood of anxiety and depression.^10^ The HADS does not include items related to sleep difficulties or somatic symptoms. This assessment tool is useful in both primary care and hospital settings for measuring symptom severity of anxiety and depression,^11^ and its psychometric properties have been validated as part of the HUNT Study.^12^

### Assumptions of Mendelian randomization (MR)

There are three core assumptions of MR (as illustrated by the directed acyclic graph presented in Figure SM1).^13,14^ These assumptions state the following:

1.	The genetic instrument must be robustly associated with the exposure (relevance assumption).

2.	The genetic instrument should not be associated with any confounders of the exposure-outcome association (independence assumption).

3.	The genetic instrument should only affect the outcome via the exposure of interest, i.e., no independent pathway except through the exposure (exclusion restriction assumption).

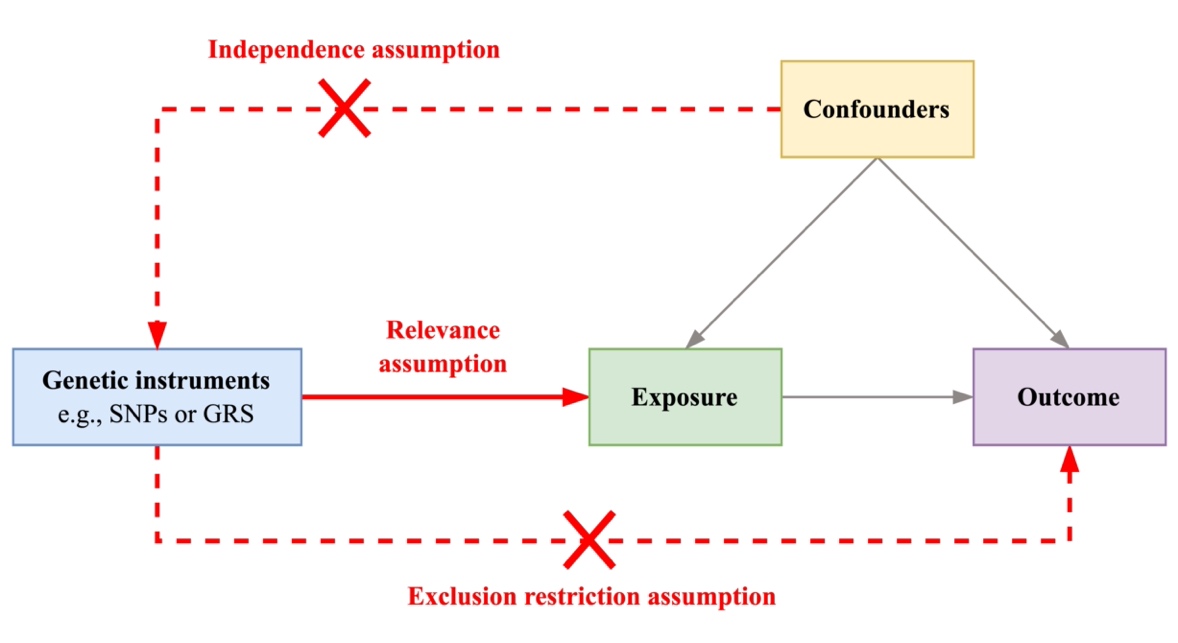

Figure SM1: Directed acyclic graph presenting Mendelian randomization assumptions.

Each of the summary-level MR methods used for the sensitivity analyses makes different assumptions about the genetic variants used, where the MR-Egger regression method gives a valid causal estimate under the instrument strength independent of direct effect (InSIDE) assumption and its intercept allows the size of any unbalanced pleiotropic effect to be determined,^15^ the weighted median method assumes that at least 50% of genetic variants are valid,^16^ and the weighted mode-based estimation method assumes that a plurality of genetic variants are valid.^17^

### Supplementary figures

Figure S1: 2x2 factorial Mendelian randomization Cox regression analysis assessing the joint effects of two sleep traits with risk of incident atrial fibrillation in HUNT2 using weighted and unweighted genetic risk scores for sleep traits

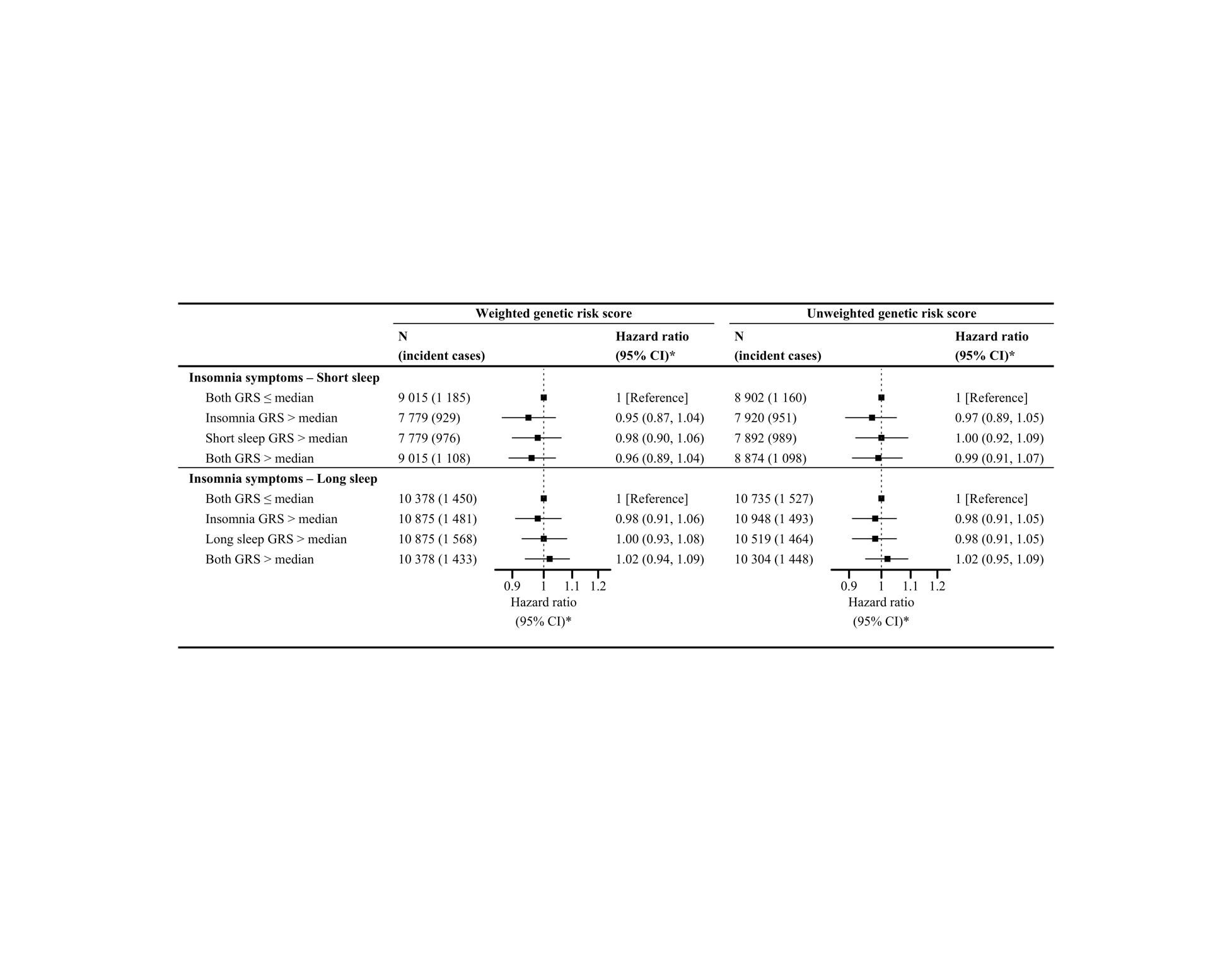

CI, confidence interval; and GRS, genetic risk score.

For each sleep trait combination, both GRS ≤ median represents low genetic risk for both sleep traits in combination, sleep trait 1 GRS > median represents high genetic risk for sleep trait 1 only, sleep trait 2 GRS > median represents high genetic risk for sleep trait 2 only and both GRS > median represents high genetic risk for both sleep traits.

**^*^** Adjusted for age, sex, 20 genetic principal components, and genotyping chip.

Figure S2: Association of insomnia SNPs from Jansen et al., 2019^18^ and atrial fibrillation (AF) within a) UK Biobank b) HUNT2. IVW, MR-Egger, simple median and weighted median estimates are indicated by the red, green, blue and purple lines respectively

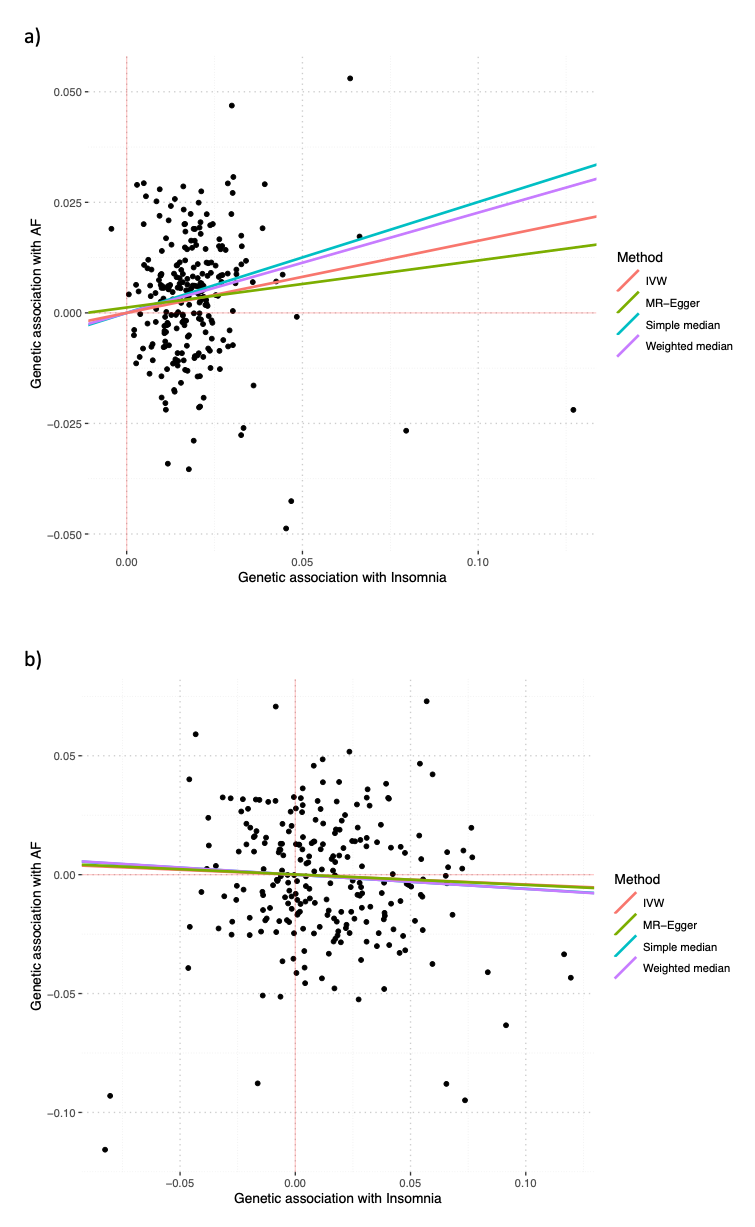

Figure S3: Association of 24-hour sleep duration SNPs from Dashti et al., 2019^19^ and atrial fibrillation (AF) within a) UK Biobank b) HUNT2. IVW, MR-Egger, simple median and weighted median estimates are indicated by the red, green, blue and purple lines respectively

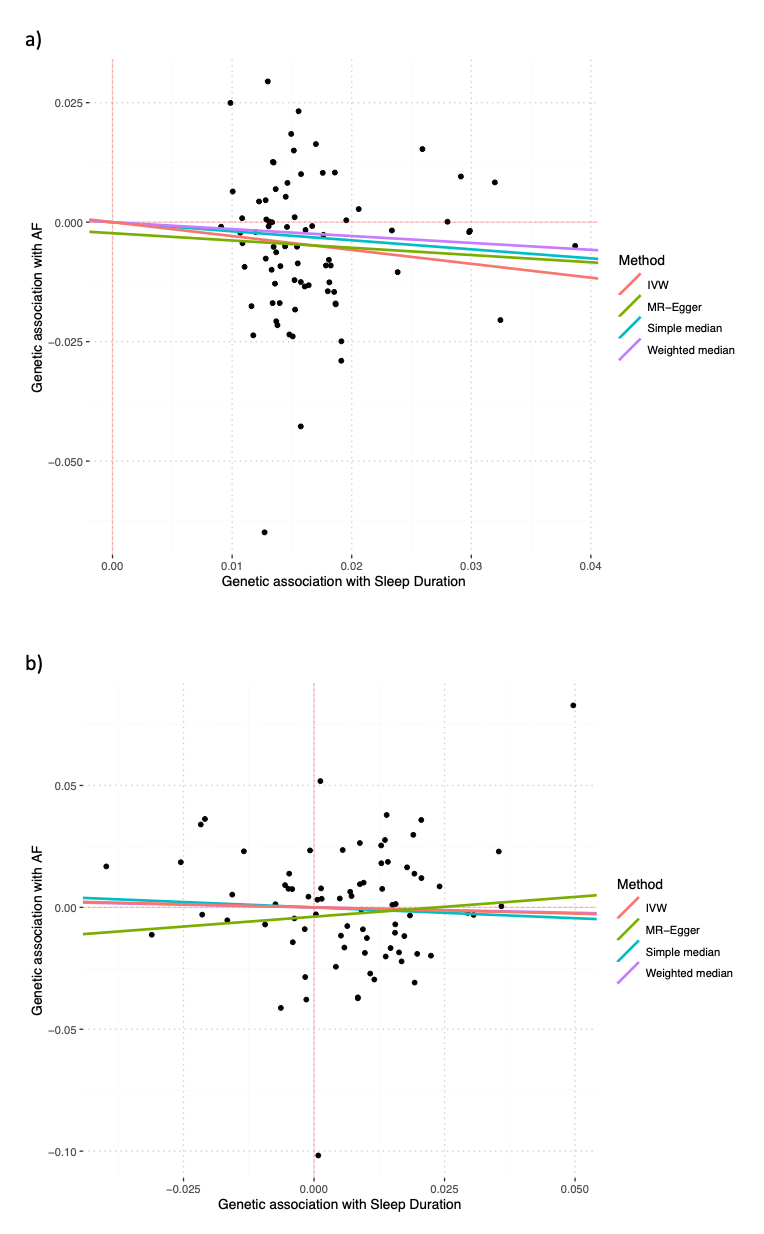

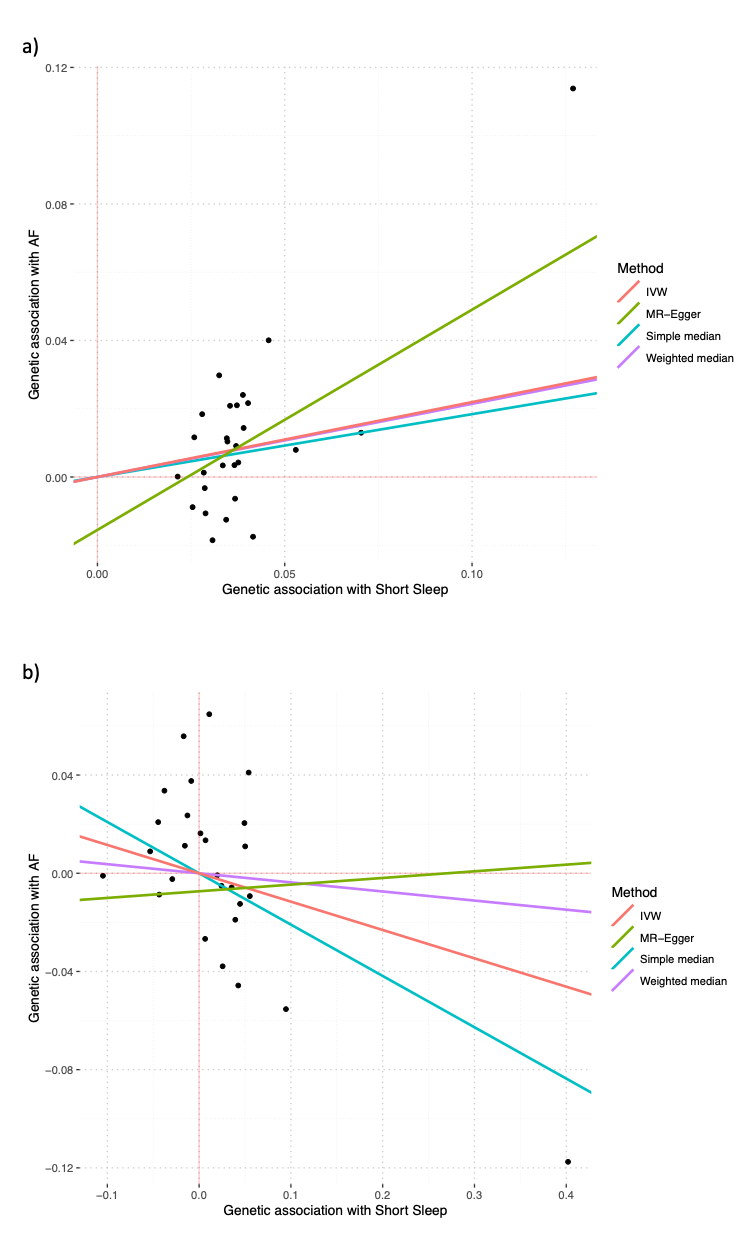
Figure S4: Association of short sleep duration SNPs from Dashti et al., 2019^19^ and atrial fibrillation (AF) within a) UK Biobank b) HUNT2. IVW, MR-Egger, simple median and weighted median estimates are indicated by the red, green, blue and purple lines respectively

Figure S5: Association of long sleep duration SNPs from Dashti et al., 2019^19^ and atrial fibrillation (AF) within a) UK Biobank b) HUNT2. IVW, MR-Egger, simple median and weighted median estimates are indicated by the red, green, blue and purple lines respectively

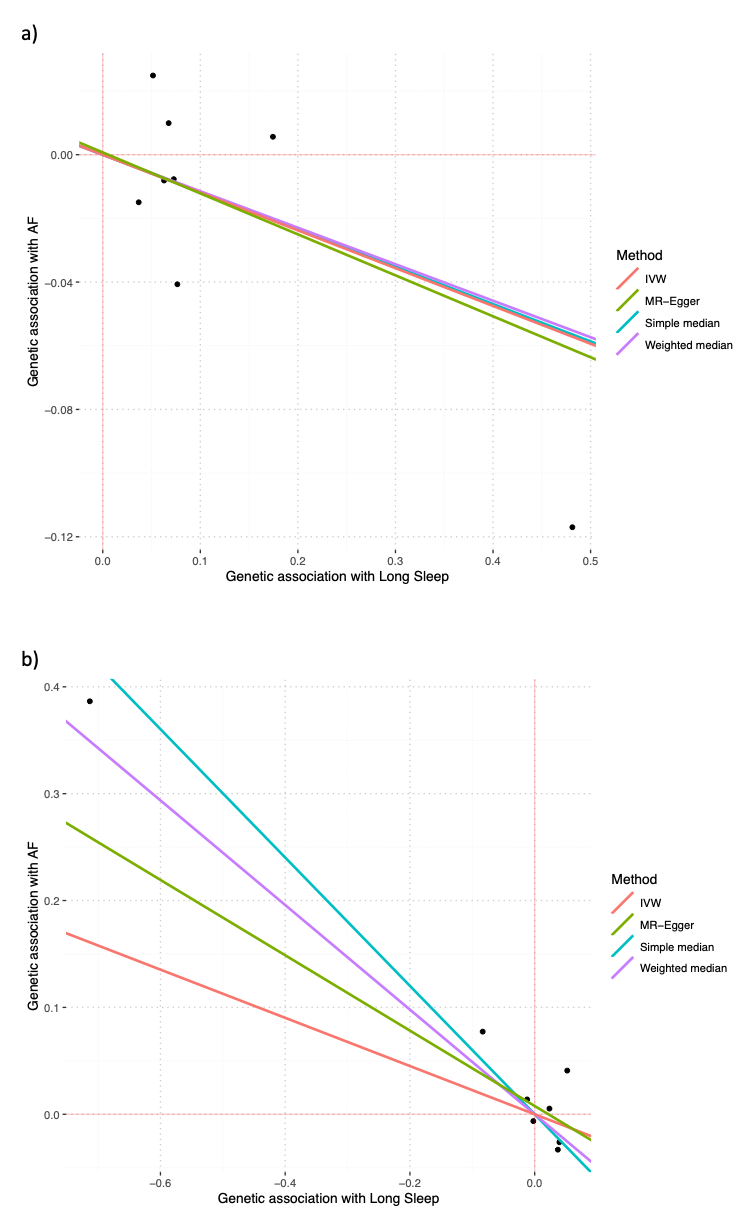

Figure S6: Association of chronotype SNPs from Jones et al., 2019^20^ and atrial fibrillation (AF) within UK Biobank. IVW, MR-Egger, simple median and weighted median estimates are indicated by the red, green, blue and purple lines respectively

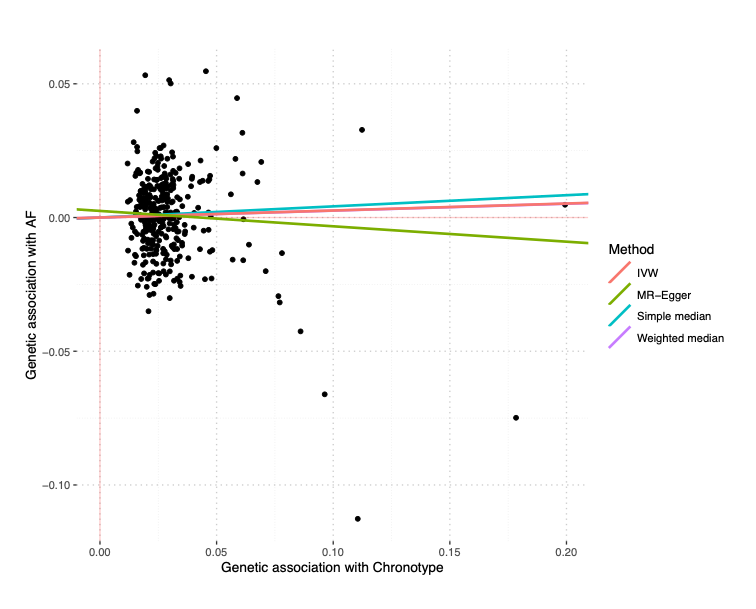

Figure S7: Association of insomnia SNPs from Lane et al., 2019^21^ and atrial fibrillation (AF) within a) UK Biobank b) HUNT2. IVW, MR-Egger, simple median and weighted median estimates are indicated by the red, green, blue and purple lines respectively

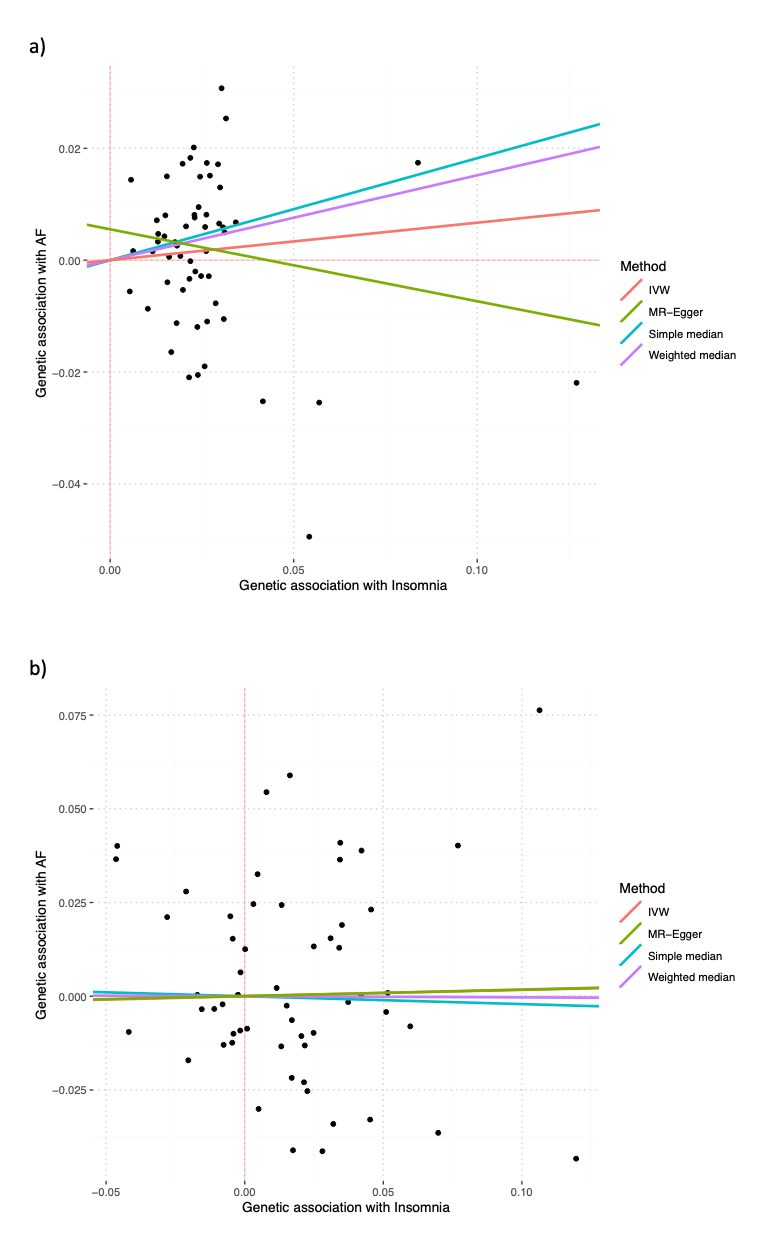

Figure S8: Continuous factorial Mendelian randomization analysis using genetic risk score as quantitative traits with their product term assessing the joint effects of two sleep traits with risk of incident atrial fibrillation in UK Biobank and HUNT2

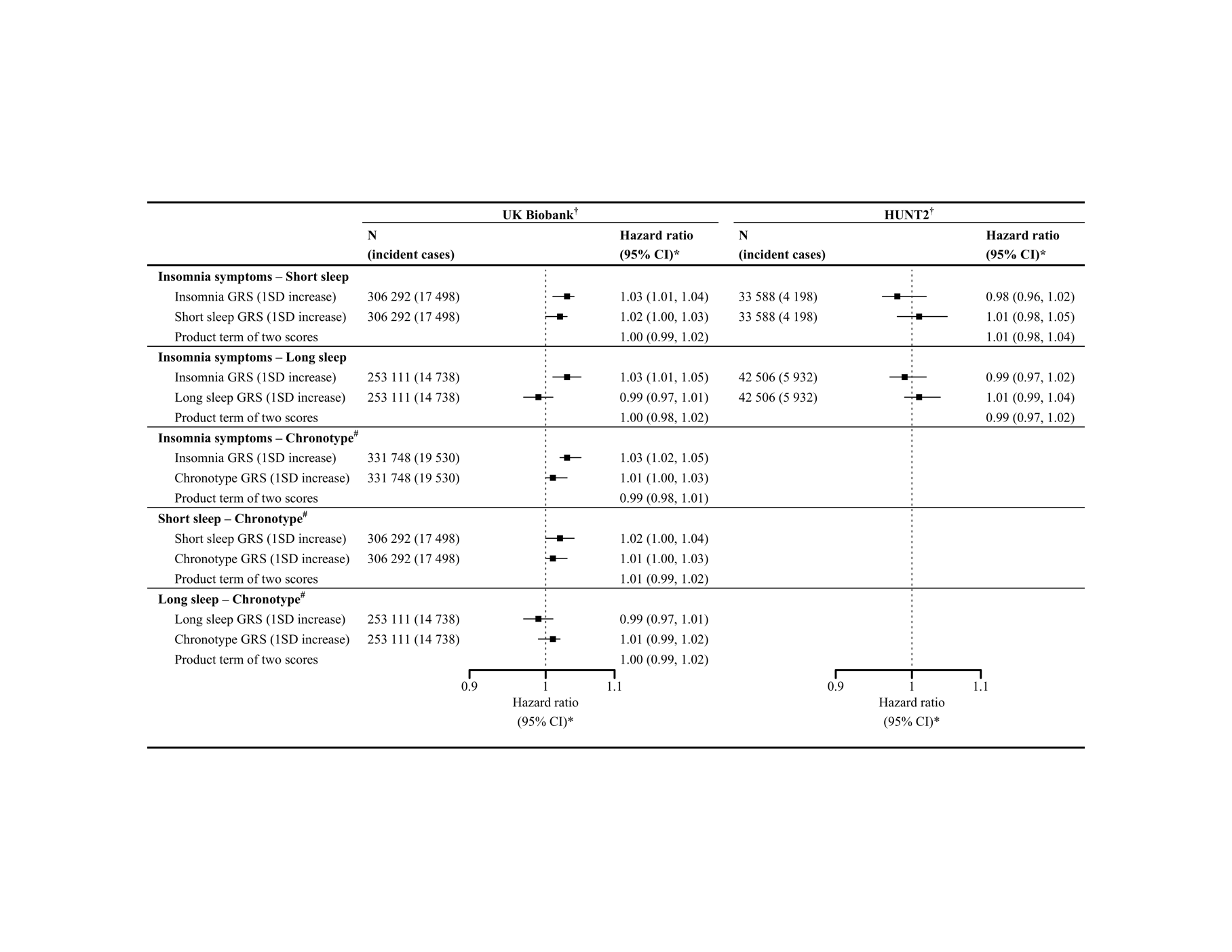

CI, confidence interval; and GRS, genetic risk score

**^†^** Derived using unweighted genetic risk score for each sleep trait in UK Biobank, whereas using weighted genetic risk score for each sleep trait in HUNT2.

**^*^** Adjusted for age, sex, assessment center (in UK Biobank), genetic principal components (40 in UK Biobank and 20 in HUNT2), and genotyping chip.

**^#^** Chronotype genetic risk score calculated using alleles for morning preference.

### Supplementary tables

Table S1: Summary of genome-wide significant genetic instruments of sleep traits in the discovery genome-wide association studies

| **Sleep traits** | **Discovery GWAS** | **PMID** | **N** | **Cohorts used by the discovery GWAS** | | **No. of SNPs identified** |
| --- | --- | --- | --- | --- | --- | --- |
|  |  |  |  | **UK Biobank** | **23andMe** |  |
| Insomnia symptoms | Jansen et al., 2019 ^18^ | **30804565** | 1 331 010 | 109 402 cases and  277 131 controls | 288 557 cases and  655 920 controls | 248 |
| 24-hour sleep duration (h) | Dashti et al., 2019 ^19^ | 30846698 | 446 118 | N.A. | Not included | 78 |
| Short sleep  (≤6 h vs. 7-8 h) | Dashti et al., 2019 ^19^ | 30846698 | 411 934 | 106 192 cases and  305 742 controls | Not included | 27 |
| Long sleep  (≥9 h vs. 7-8 h) | Dashti et al., 2019 ^19^ | 30846698 | 339 926 | 34 184 cases and  305 742 controls | Not included | 8 |
| Chronotype  (morning preference)^*^ | Jones et al., 2019 ^20^ | 30696823 | 651 295 | 252 287 cases and  150 908 controls | 120 478 cases and  127 622 controls | 351 |

GWAS, genome-wide association studies; N, sample size; SNPs, single nucleotide polymorphisms; and N.A., Not applicable.

^*^ In the discovery GWAS of chronotype, the chronotype increasing allele is morning preference.

Table S2: Baseline characteristics of study participants included in Mendelian randomization analysis with and without a diagnosis of atrial fibrillation (AF) during follow-up in UK Biobank and HUNT2

|  |  | | **UK Biobank (N = 331 748)** | | | |  | | **HUNT2 (N = 45 322)** | | |
| --- | --- | --- | --- | --- | --- | --- | --- | --- | --- | --- | --- |
|  |  |  | **No AF diagnosis** | | **AF diagnosis** | |  | | **No AF diagnosis** | | **AF diagnosis** |
|  | **Total, % (n)** | | 94.11 (312 218) | | 5.89 (19 530) | |  | | 86.11 (39 026) | | 13.89 (6 296) |
|  | **Variables, % (n)** | | | | | | | | | | |
|  | Male | | 43.63 (136 208) | | 61.56 (12 023) | |  | | 44.52 (17 374) | | 55.51 (3 495) |
|  | Missing, % (n) | | - | | - | |  | | - | | - |
|  | Married | | 74.19 (231 631) | | 73.47 (14 349) | |  | | 61.20 (23 885) | | 71.16 (4 480) |
| Missing, % (n) | | 0.46 (1 451) | | 0.87 (169) | |  | | 0.22 (86) | | 0.13 (8) | |
|  | Weekly alcohol intake | | 50.78 (158 531) | | 46.68 (9 116) | |  | | 22.58 (8 813) | | 19.46 (1 225) |
|  | Missing, % (n) | | 0.05 (147) | | 0.10 (19) | |  | | 7.09 (2 768) | | 10.32 (650) |
|  | Current smokers | | 10.20 (31 833) | | 11.02 (2 153) | |  | | 28.64 (11 178) | | 22.43 (1 412) |
|  | Missing, % (n) | | 0.28 (889) | | 0.44 (85) | |  | | 1.53 (599) | | 1.87 (118) |
|  | Highly physically active | | 33.45 (104 450) | | 32.42 (6 332) | |  | | 33.43 (13 046) | | 24.01 (1 511) |
|  | Missing, % (n) | | 17.62 (55 027) | | 19.17 (3 744) | |  | | 7.13 (2 782) | | 14.56 (917) |
|  | Tertiary education | | 43.31 (135 228) | | 38.83 (7 583) | |  | | 21.65 (8 449) | | 13.88 (874) |
|  | Missing, % (n) | | 0.74 (2 299) | | 1.00 (195) | |  | | 3.17 (1 239) | | 6.42 (404) |
|  | Shift workers | | 5.30 (16 536) | | 3.53 (689) | |  | | 16.28 (6 353) | | 7.70 (485) |
|  | Missing, % (n) | | 0.27 (847) | | 0.26 (50) | |  | | 7.20 (2 810) | | 6.97 (439) |
|  | Employed | | 58.40 (182 336) | | 35.92 (7 016) | |  | | 69.18 (26 999) | | 45.20 (2 846) |
|  | Missing, % (n) | | 0.24 (741) | | 0.19 (38) | |  | | 0.95 (371) | | 0.59 (37) |
|  | Use of sleep medication(s) | | 0.94 (2 926) | | 1.58 (309) | |  | | 6.01 (2 345) | | 10.44 (657) |
|  | Missing, % (n) | | - | | - | |  | | 9.38 (3 659) | | 10.15 (639) |
|  | Suffering from depression | | 12.01 (37 460) | | 14.51 (2 833) | |  | | - | | - |
|  | Missing, % (n) | | - | | - | |  | | - | | - |
|  | Suffering from anxiety | | 6.51 (20 316) | | 9.72 (1 898) | |  | | - | | - |
|  | Missing, % (n) | | - | | - | |  | | - | | - |
|  | Suffering from chronic illness | | 30.13 (94 065) | | 47.48 (9 272) | |  | | 30.05 (11 727) | | 47.73 (3 005) |
|  | Missing, % (n) | | 2.01 (6 291) | | 2.38 (464) | |  | | 2.95 (1 150) | | 4.67 (294) |
|  | **Variables, mean (SD)** | | | | | | | | | | |
|  | Age, *years* | | 56.55 (7.93) | | 62.13 (5.90) | |  | | 47.22 (15.98) | | 62.09 (12.39) |
|  | Missing, % (n) | | - | | - | |  | | - | | - |
|  | TDI | | -1.60 (2.91) | | -1.38 (3.04) | |  | | - | | - |
|  | Missing, % (n) | | 0.12 (381) | | 0.08 (16) | |  | | - | | - |
|  | BMI, *kg/m^2^* | | 27.30 (4.69) | | 28.96 (5.36) | |  | | 26.11 (3.99) | | 27.60 (4.21) |
|  | Missing, % (n) | | 0.29 (904) | | 0.53 (104) | |  | | 0.50 (194) | | 0.48 (30) |
|  | SBP, *mmHg* | | 138.10 (18.54) | | 143.80 (19.25) | |  | | 135.10 (20.28) | | 147.80 (22.47) |
|  | Missing, % (n) | | 0.09 (272) | | 0.11 (21) | |  | | 0.11 (41) | | 0.10 (6) |
|  | Serum cholesterol, *mmol/L* | | 5.75 (1.14) | | 5.43 (1.18) | |  | | 5.81 (1.24) | | 6.26 (1.21) |
|  | Missing, % (n) | | 4.55 (14 214) | | 4.50 (878) | |  | | 0.13 (49) | | 0.03 (2) |
|  | Blood glucose, *mmol/L* | | 5.10 (1.17) | | 5.32 (1.50) | |  | | 5.38 (1.41) | | 5.75 (1.71) |
|  | Missing, % (n) | | 12.73 (39 730) | | 12.57 (2 454) | |  | | 0.17 (68) | | 0.10 (6) |
|  | HADS - D scores | | - | | - | |  | | 3.30 (2.98) | | 4.01 (3.12) |
|  | Missing, % (n) | | - | | - | |  | | 6.42 (2 507) | | 10.32 (650) |
|  | HADS - A scores | | - | | - | |  | | 4.19 (3.26) | | 4.01 (3.24) |
|  | Missing, % (n) | | - | | - | |  | | 12.80 (4 997) | | 21.98 (1 384) |

AF, atrial fibrillation; SD, standard deviation; TDI, Townsend deprivation index; BMI, body mass index; SBP, systolic blood pressure; HADS – D scores, Hospital Anxiety and Depression Scale – Depression scores; and HADS – A scores, Hospital Anxiety and Depression Scale – Anxiety scores.

Table S3: Summary of genetic instruments showing their strength applying to UK Biobank and HUNT2

| **UK Biobank** | | | | | | |
| --- | --- | --- | --- | --- | --- | --- |
| **Sleep traits** | **N** | **No. of SNPs to generate the uwGRS** | **Mean (SD) no. of increasing allele** | **Association of uwGRS with sleep trait ^†^** | | |
|  |  |  |  | **Coefficient (SE)** | **R^2^ ^§^** | **F-statistics** **^§§^** |
| Insomnia symptoms | 331 748 | 248 | 245.17 (10.27) | 0.1561 (0.0039) | 0.41% | 1369.47 |
| 24-hour sleep duration (h) | 331 748 | 78 | 76.26 (5.43) | 0.0832 (0.0019) | 0.60% | 1993.0 |
| Short sleep  (≤6 h vs. 7-8 h) | 306 292 | 27 | 26.34 (3.15) | 0.1045 (0.0042) | 0.18% | 558.61 |
| Long sleep  (≥9 h vs. 7-8 h) | 253 111 | 8 | 4.10 (1.42) | 0.0874 (0.0066) | 0.11% | 270.36 |
| Chronotype  (morning preference)^*^ | 331 748 | 341 ^#^ | 334.12 (11.64) | 0.2995 (0.0037) | 1.54% | 5176.33 |
| **HUNT2** | | | | | | |
| **Sleep traits** | **N** | **No. of SNPs to generate the wGRS** | **Mean (SD) no. of increasing allele** | **Association of wGRS with sleep trait ^‡^** | | |
|  |  |  |  | **Coefficient (SE)** | **R^2 §^** | **F-statistics ^§§^** |
| Insomnia symptoms | 45 322 | 244 ^#^ | 240.40 (10.16) | 0.1037 (0.0136) | 0.16% | 72.38 |
| 24-hour sleep duration (h) | 45 322 | 78 | 77.25 (5.35) | 0.0376 (0.0058) | 0.09% | 42.63 |
| Short sleep  (≤6 h vs. 7-8 h) | 33 588 | 27 | 26.12 (3.12) | 0.0346 (0.0197) | 0.02% | 5.35 |
| Long sleep  (≥9 h vs. 7-8 h) | 42 506 | 8 | 4.18 (1.39) | 0.0247 (0.0108) | 0.01% | 4.42 |

SNPs, single nucleotide polymorphisms; SD, standard deviation; uwGRS, unweighted genetic risk score; wGRS, weighted genetic risk score; and SE, standard error

**^†^** Adjusted for age, sex, assessment center, 40 principal components, and genotyping chip.

**^‡^** Adjusted for age, sex, 20 principal components, and genotyping chip.

**^§^** McFadden R^2^ statistics for sleep traits – insomnia symptoms, short sleep, long sleep and chronotype.

**^§§^** F-statistics was calculated using F = (R^2^/K) / ((1 – R^2^) / (N-K-1)); where R^2^ = McFadden R^2^ statistics, K = 1, and N = sample size.

^*^ Morning preference vs. Evening preference

^#^ rs146820337, rs112201801, rs10610420, rs9991917, rs67169439, rs34125199, rs60521023, rs3747463, rs213462, and rs7060620 were absent in the imputed UK Biobank genetic data; and rs1264419, rs138678612, rs238869 and rs3131638 were absent in the imputed HUNT genetic data.

Table S4: Baseline characteristics of participants across groups categorized by dichotomizing to the median genetic risk scores for insomnia symptoms and short sleep in UK Biobank

|  | **UK Biobank (N = 306 292)** | | | |
| --- | --- | --- | --- | --- |
|  | **Both GRS**  **≤ median** | **Insomnia GRS**  **> median** | **Short sleep GRS**  **> median** | **Both GRS**  **> median** |
| **Total, % (n)** | 26.67 (81 693) | 23.33 (71 453) | 23.33 (71 453) | 26.67 (81 693) |
| **Variables, % (n)** | | | | |
| Male | 44.81 (36 608) | 44.55 (31 832) | 44.95 (32 116) | 45.09 (36 838) |
| Missing, % (n) | - | - | - | - |
| Married | 74.51 (60 870) | 74.06 (52 921) | 74.32 (53 101) | 73.97 (60 427) |
| Missing, % (n) | 0.45 (365) | 0.50 (358) | 0.45 (325) | 0.49 (401) |
| Weekly alcohol intake | 51.48 (42 053) | 50.78 (36 282) | 51.27 (36 636) | 50.60 (41 334) |
| Missing, % (n) | 0.04 (35) | 0.06 (42) | 0.04 (27) | 0.05 (42) |
| Current smokers | 9.52 (7 779) | 10.50 (7 501) | 9.67 (6 908) | 10.76 (8 788) |
| Missing, % (n) | 0.28 (228) | 0.28 (198) | 0.31 (219) | 0.30 (244) |
| Highly physically active | 34.01 (27 784) | 33.41 (23 874) | 33.67 (24 061) | 33.49 (27 361) |
| Missing, % (n) | 17.19 (14 041) | 17.76 (12 693) | 17.21 (12 294) | 18.07 (14 760) |
| Tertiary education | 45.15 (36 883) | 43.22 (30 883) | 43.78 (31 283) | 42.27 (34 534) |
| Missing, % (n) | 0.69 (562) | 0.76 (540) | 0.71 (505) | 0.77 (630) |
| Shift workers | 5.17 (4 224) | 5.36 (3 829) | 5.28 (3 771) | 5.44 (4 445) |
| Missing, % (n) | 0.25 (201) | 0.24 (169) | 0.29 (205) | 0.29 (237) |
| Employed | 59.55 (48 646) | 59.15 (42 266) | 58.83 (42 038) | 58.72 (47 974) |
| Missing, % (n) | 0.22 (177) | 0.21 (151) | 0.25 (176) | 0.25 (201) |
| Use of sleep medication(s) | 0.78 (639) | 0.94 (673) | 0.88 (632) | 1.02 (836) |
| Missing, % (n) | - | - | - | - |
| Suffering from depression | 10.59 (8 650) | 12.15 (8 681) | 10.85 (7 751) | 12.34 (10 077) |
| Missing, % (n) | - | - | - | - |
| Suffering from anxiety | 6.11 (4 994) | 6.76 (4 832) | 6.20 (4 433) | 6.79 (5 551) |
| Missing, % (n) | - | - | - | - |
| Suffering from chronic illness | 28.40 (23 204) | 31.11 (22 227) | 28.97 (20 700) | 31.94 (26 089) |
| Missing, % (n) | 1.91 (1 561) | 2.13 (1 524) | 1.96 (1 397) | 2.15 (1 755) |
| **Variables, mean (SD)** | | | | |
| Age, *years* | 56.72 (7.92) | 56.65 (7.94) | 56.81 (7.90) | 56.67 (7.94) |
| Missing, % (n) | - | - | - | - |
| TDI | -1.66 (2.87) | -1.57 (2.92) | -1.64 (2.89) | -1.54 (2.94) |
| Missing, % (n) | 0.11 (92) | 0.11 (76) | 0.13 (95) | 0.12 (102) |
| BMI, *kg/m^2^* | 27.16 (4.62) | 27.43 (4.77) | 27.23 (4.65) | 27.53 (4.80) |
| Missing, % (n) | 0.26 (213) | 0.30 (217) | 0.30 (214) | 0.29 (240) |
| SBP, *mmHg* | 138.0 (18.64) | 138.30 (18.60) | 138.20 (18.55) | 138.40 (18.58) |
| Missing, % (n) | 0.08 (69) | 0.10 (70) | 0.07 (52) | 0.08 (62) |
| Serum cholesterol, *mmol/L* | 5.75 (1.13) | 5.73 (1.13) | 5.74 (1.14) | 5.72 (1.14) |
| Missing, % (n) | 4.53 (3 700) | 4.59 (3 281) | 4.57 (3 262) | 4.44 (3 626) |
| Blood glucose, *mmol/L* | 5.09 (1.12) | 5.12 (1.21) | 5.10 (1.14) | 5.11 (1.19) |
| Missing, % (n) | 12.69 (10 366) | 12.76 (9 120) | 12.74 (9 101) | 12.68 (10 361) |

GRS, genetic risk score; SD, standard deviation; TDI, Townsend deprivation index; BMI, body mass index; and SBP, systolic blood pressure.

Table S5: Baseline characteristics of participants across groups categorized by dichotomizing to the median genetic risk scores for insomnia symptoms and short sleep in HUNT2

|  | **HUNT2 (N = 33 588)** | | | |
| --- | --- | --- | --- | --- |
|  | **Both GRS**  **≤ median** | **Insomnia GRS**  **> median** | **Short sleep GRS**  **> median** | **Both GRS**  **> median** |
| **Total, % (n)** | 26.84 (9 015) | 23.16 (7 779) | 23.16 (7 779) | 26.84 (9 015) |
| **Variables, % (n)** | | | | |
| Male | 48.57 (4 379) | 47.77 (3 716) | 48.01 (3 735) | 46.84 (4 223) |
| Missing, % (n) | - | - | - | - |
| Married | 64.35 (5 801) | 64.07 (4 984) | 62.82 (4 887) | 62.88 (5 669) |
| Missing, % (n) | - | - | - | - |
| Weekly alcohol intake | 23.69 (2 136) | 23.83 (1 854) | 23.46 (1 825) | 23.02 (2 075) |
| Missing, % (n) | 6.84 (617) | 7.02 (546) | 7.04 (548) | 7.34 (662) |
| Current smokers | 27.54 (2 483) | 29.95 (2 330) | 27.15 (2 112) | 29.66 (2 674) |
| Missing, % (n) | 1.32 (119) | 1.49 (116) | 1.32 (103) | 1.42 (128) |
| Highly physically active | 35.67 (3 216) | 34.35 (2 672) | 34.86 (2 712) | 33.43 (3 014) |
| Missing, % (n) | 6.47 (583) | 6.45 (502) | 6.83 (531) | 7.31 (659) |
| Tertiary education | 22.91 (2 065) | 22.80 (1 774) | 22.14 (1 722) | 21.86 (1 971) |
| Missing, % (n) | 2.78 (251) | 2.73 (212) | 3.01 (234) | 3.16 (285) |
| Shift workers | 15.93 (1 436) | 16.93 (1 317) | 16.54 (1 287) | 16.54 (1 491) |
| Missing, % (n) | 7.68 (692) | 7.60 (591) | 7.84 (610) | 7.68 (692) |
| Employed | 73.84 (6 657) | 74.21 (5 773) | 72.90 (5 671) | 72.66 (6 550) |
| Missing, % (n) | 0.85 (77) | 0.90 (70) | 0.75 (58) | 0.93 (84) |
| Use of sleep medication(s) | 4.50 (406) | 5.01 (390) | 4.76 (370) | 5.56 (501) |
| Missing, % (n) | 9.33 (841) | 9.31 (724) | 9.74 (758) | 9.01 (811) |
| Suffering from chronic illness | 27.86 (2 512) | 28.31 (2 202) | 26.48 (2 060) | 29.83 (2 689) |
| Missing, % (n) | 2.85 (257) | 2.84 (221) | 2.98 (232) | 3.11 (280) |
| **Variables, mean (SD)** | | | | |
| Age, *years* | 47.72 (14.99) | 47.10 (14.87) | 47.28 (15.14) | 47.38 (15.07) |
| Missing, % (n) | - | - | - | - |
| BMI, *kg/m^2^* | 26.13 (3.92) | 26.24 (3.90) | 26.16 (3.96) | 26.32 (4.07) |
| Missing, % (n) | 0.19 (17) | 0.30 (23) | 0.28 (22) | 0.28 (25) |
| SBP, *mmHg* | 135.60 (20.18) | 135.00 (19.95) | 135.60 (19.93) | 135.30 (20.10) |
| Missing, % (n) | 0.06 (5) | 0.09 (7) | 0.15 (12) | 0.12 (11) |
| Serum cholesterol, *mmol/L* | 5.82 (1.21) | 5.81 (1.24) | 5.80 (1.21) | 5.80 (1.22) |
| Missing, % (n) | 0.04 (4) | 0.18 (14) | 0.15 (12) | 0.10 (9) |
| Blood glucose, *mmol/L* | 5.34 (1.25) | 5.36 (1.28) | 5.35 (1.32) | 5.38 (1.34) |
| Missing, % (n) | 0.08 (7) | 0.21 (16) | 0.17 (13) | 0.16 (14) |
| HADS - D scores | 3.25 (2.87) | 3.28 (2.94) | 3.20 (2.89) | 3.29 (2.95) |
| Missing, % (n) | 6.12 (552) | 5.39 (419) | 6.20 (482) | 6.46 (582) |
| HADS - A scores | 4.11 (3.18) | 4.20 (3.28) | 4.03 (3.11) | 4.22 (3.27) |
| Missing, % (n) | 12.21 (1 101) | 12.10 (941) | 12.32 (958) | 12.86 (1 159) |

GRS, genetic risk score; SD, standard deviation; BMI, body mass index; SBP, systolic blood pressure; HADS – D scores, Hospital Anxiety and Depression Scale – Depression scores; and HADS – A scores, Hospital Anxiety and Depression Scale – Anxiety scores.

Table S6: Baseline characteristics of participants across groups categorized by dichotomizing to the median genetic risk scores for insomnia symptoms and long sleep in UK Biobank

|  | **UK Biobank (N = 253 111)** | | | |
| --- | --- | --- | --- | --- |
|  | **Both GRS**  **≤ median** | **Insomnia GRS**  **> median** | **Long sleep GRS**  **> median** | **Both GRS**  **> median** |
| **Total, % (n)** | 28.90 (73 153) | 29.55 (74 794) | 21.10 (53 403) | 20.45 (51 761) |
| **Variables, % (n)** | | | | |
| Male | 44.65 (32 664) | 44.67 (33 410) | 43.82 (23 401) | 44.37 (22 967) |
| Missing, % (n) | - | - | - | - |
| Married | 76.12 (55 682) | 75.71 (56 630) | 75.65 (40 400) | 75.61 (39 134) |
| Missing, % (n) | 0.42 (304) | 0.41 (310) | 0.39 (208) | 0.48 (247) |
| Weekly alcohol intake | 51.59 (37 736) | 50.95 (38 105) | 51.31 (27 399) | 50.88 (26 334) |
| Missing, % (n) | 0.03 (25) | 0.04 (29) | 0.04 (23) | 0.06 (32) |
| Current smokers | 8.96 (6 552) | 9.94 (7 432) | 9.28 (4 956) | 10.04 (5 198) |
| Missing, % (n) | 0.30 (219) | 0.27 (201) | 0.26 (137) | 0.25 (129) |
| Highly physically active | 33.83 (24 747) | 33.51 (25 067) | 33.61 (17 950) | 33.01 (17 085) |
| Missing, % (n) | 16.74 (12 248) | 17.37 (12 988) | 16.94 (9044) | 17.60 (9 111) |
| Tertiary education | 45.18 (33 050) | 43.61 (32 620) | 44.83 (23 942) | 43.39 (22 457) |
| Missing, % (n) | 0.67 (489) | 0.74 (554) | 0.64 (341) | 0.69 (356) |
| Shift workers | 4.53 (3 316) | 4.58 (3 429) | 4.38 (2 338) | 4.55 (2 356) |
| Missing, % (n) | 0.27 (200) | 0.26 (193) | 0.26 (139) | 0.26 (133) |
| Employed | 56.31 (41 196) | 56.35 (42 149) | 56.19 (30 006) | 56.12 (29 048) |
| Missing, % (n) | 0.24 (172) | 0.22 (166) | 0.22 (119) | 0.22 (115) |
| Use of sleep medication(s) | 0.67 (493) | 0.82 (611) | 0.77 (409) | 0.85 (435) |
| Missing, % (n) | - | - | - | - |
| Suffering from depression | 10.93 (7 993) | 12.21 (9 132) | 10.65 (5 685) | 12.03 (6 229) |
| Missing, % (n) | - | - | - | - |
| Suffering from anxiety | 5.98 (4 374) | 6.59 (4 929) | 6.10 (3 260) | 6.71 (3 474) |
| Missing, % (n) | - | - | - | - |
| Suffering from chronic illness | 28.36 (20 745) | 31.13 (23 281) | 28.18 (15 047) | 30.91 (15 997) |
| Missing, % (n) | 1.75 (1 277) | 1.92 (1 436) | 1.88 (1 005) | 2.05 (1 061) |
| **Variables, mean (SD)** | | | | |
| Age, *years* | 56.98 (7.96) | 56.85 (8.02) | 56.93 (8.01) | 56.84 (8.04) |
| Missing, % (n) | - | - | - | - |
| TDI | -1.74 (2.83) | -1.65 (2.86) | -1.73 (2.84) | -1.66 (2.88) |
| Missing, % (n) | 0.13 (98) | 0.11 (80) | 0.10 (56) | 0.13 (67) |
| BMI, *kg/m^2^* | 27.17 (4.60) | 27.44 (4.75) | 27.01 (4.50) | 27.27 (4.64) |
| Missing, % (n) | 0.28 (205) | 0.31 (234) | 0.30 (162) | 0.26 (132) |
| SBP, *mmHg* | 138.30 (18.69) | 138.50 (18.74) | 138.10 (18.77) | 138.40 (18.72) |
| Missing, % (n) | 0.07 (49) | 0.09 (68) | 0.09 (47) | 0.09 (46) |
| Serum cholesterol, *mmol/L* | 5.74 (1.14) | 5.71 (1.14) | 5.75 (1.14) | 5.73 (1.14) |
| Missing, % (n) | 4.52 (3 308) | 4.50 (3 367) | 4.66 (2 487) | 4.59 (2 376) |
| Blood glucose, *mmol/L* | 5.10 (1.17) | 5.12 (1.20) | 5.09 (1.13) | 5.11 (1.18) |
| Missing, % (n) | 12.61 (9 227) | 12.63 (9 449) | 12.93 (6 907) | 12.86 (6 656) |

GRS, genetic risk score; SD, standard deviation; TDI, Townsend deprivation index; BMI, body mass index; and SBP, systolic blood pressure.

Table S7: Baseline characteristics of participants across groups categorized by dichotomizing to the median genetic risk scores for insomnia symptoms and long sleep in HUNT2

|  | **HUNT2 (N = 42 506)** | | | |
| --- | --- | --- | --- | --- |
|  | **Both GRS**  **≤ median** | **Insomnia GRS**  **> median** | **Long sleep GRS**  **> median** | **Both GRS**  **> median** |
| **Total, % (n)** | 24.42 (10 378) | 25.58 (10 875) | 25.58 (10 875) | 24.42 (10 378) |
| **Variables, % (n)** | | | | |
| Male | 45.68 (4 741) | 45.53 (4 951) | 45.77 (4 977) | 44.59 (4 628) |
| Missing, % (n) | - | - | - | - |
| Married | 63.06 (6 544) | 63.57 (6 913) | 63.19 (6 872) | 62.01 (6 435) |
| Missing, % (n) | - | - | - | - |
| Weekly alcohol intake | 22.14 (2 298) | 21.91 (2 383) | 22.17 (2 411) | 22.37 (2 322) |
| Missing, % (n) | 7.17 (744) | 7.52 (818) | 7.65 (832) | 7.77 (806) |
| Current smokers | 26.39 (2 739) | 27.74 (3 017) | 26.03 (2 831) | 28.85 (2 994) |
| Missing, % (n) | 1.64 (170) | 1.52 (165) | 1.63 (177) | 1.53 (159) |
| Highly physically active | 33.00 (3 425) | 31.51 (3 427) | 32.16 (3 497) | 31.56 (3 275) |
| Missing, % (n) | 7.94 (824) | 8.13 (884) | 8.24 (896) | 8.32 (863) |
| Tertiary education | 20.71 (2 149) | 20.50 (2 229) | 21.42 (2 329) | 21.00 (2 179) |
| Missing, % (n) | 3.65 (379) | 3.49 (380) | 3.64 (396) | 3.69 (383) |
| Shift workers | 15.11 (1 568) | 14.72 (1 601) | 14.10 (1 533) | 15.25 (1 583) |
| Missing, % (n) | 7.03 (730) | 6.90 (750) | 7.33 (797) | 7.22 (749) |
| Employed | 66.01 (6 851) | 65.02 (7 071) | 64.75 (7 042) | 66.01 (6 851) |
| Missing, % (n) | 0.83 (86) | 0.94 (102) | 0.88 (96) | 0.95 (99) |
| Use of sleep medication(s) | 6.24 (648) | 7.25 (788) | 6.25 (680) | 6.93 (719) |
| Missing, % (n) | 9.73 (1 010) | 9.21 (1 002) | 9.66 (1 051) | 9.18 (953) |
| Suffering from chronic illness | 30.98 (3 215) | 33.70 (3 665) | 32.31 (3 514) | 32.77 (3 401) |
| Missing, % (n) | 3.13 (325) | 3.06 (333) | 3.12 (339) | 3.24 (336) |
| **Variables, mean (SD)** | | | | |
| Age, *years* | 49.44 (16.43) | 49.45 (16.30) | 49.85 (16.49) | 49.06 (16.39) |
| Missing, % (n) | - | - | - | - |
| BMI, *kg/m^2^* | 26.27 (4.05) | 26.40 (4.09) | 26.20 (3.98) | 26.33 (4.07) |
| Missing, % (n) | 0.50 (52) | 0.53 (58) | 0.52 (57) | 0.45 (47) |
| SBP, *mmHg* | 137.10 (21.21) | 136.80 (21.08) | 137.30 (21.25) | 136.80 (20.98) |
| Missing, % (n) | 0.08 (8) | 0.12 (13) | 0.12 (13) | 0.11 (11) |
| Serum cholesterol, *mmol/L* | 5.89 (1.25) | 5.88 (1.25) | 5.91 (1.25) | 5.86 (1.25) |
| Missing, % (n) | 0.12 (12) | 0.15 (16) | 0.09 (10) | 0.10 (10) |
| Blood glucose, *mmol/L* | 5.42 (1.47) | 5.45 (1.58) | 5.43 (1.42) | 5.42 (1.39) |
| Missing, % (n) | 0.17 (18) | 0.18 (20) | 0.14 (15) | 0.14 (15) |
| HADS - D scores | 3.33 (2.98) | 3.41 (3.02) | 3.34 (2.95) | 3.37 (2.98) |
| Missing, % (n) | 6.91 (717) | 6.74 (733) | 7.29 (793) | 6.76 (702) |
| HADS - A scores | 4.08 (3.17) | 4.18 (3.29) | 4.05 (3.18) | 4.18 (3.27) |
| Missing, % (n) | 13.92 (1 445) | 14.26 (1 551) | 14.11 (1 535) | 13.85 (1 437) |

GRS, genetic risk score; SD, standard deviation; BMI, body mass index; SBP, systolic blood pressure; HADS – D scores, Hospital Anxiety and Depression Scale – Depression scores; and HADS – A scores, Hospital Anxiety and Depression Scale – Anxiety scores.

Table S8: Baseline characteristics of participants across groups categorized by dichotomizing to the median genetic risk scores for insomnia symptoms and chronotype (morning preference) in UK Biobank

|  | **UK Biobank (N = 331 748)** | | | |
| --- | --- | --- | --- | --- |
|  | **Both GRS**  **≤ median** | **Insomnia GRS**  **> median** | **Chronotype GRS**  **> median** | **Both GRS**  **> median** |
| **Total, % (n)** | 24.14 (80 092) | 25.86 (85 782) | 25.86 (85 782) | 24.14 (80 092) |
| **Variables, % (n)** | | | | |
| Male | 44.73 (35 824) | 44.41 (38 092) | 44.68 (38 325) | 44.94 (35 990) |
| Missing, % (n) | - | - | - | - |
| Married | 73.99 (59 263) | 73.58 (63 115) | 74.71 (64 086) | 74.31 (59 516) |
| Missing, % (n) | 0.47 (377) | 0.52 (446) | 0.47 (406) | 0.49 (391) |
| Weekly alcohol intake | 50.82 (40 699) | 50.01 (42 891) | 50.96 (43 716) | 50.37 (40 341) |
| Missing, % (n) | 0.04 (33) | 0.07 (58) | 0.05 (44) | 0.04 (31) |
| Current smokers | 9.83 (7 873) | 10.96 (9 405) | 9.60 (8 238) | 10.58 (8 470) |
| Missing, % (n) | 0.29 (235) | 0.29 (245) | 0.30 (258) | 0.29 (236) |
| Highly physically active | 33.17 (26 566) | 32.73 (28 080) | 34.01 (29 169) | 33.67 (26 967) |
| Missing, % (n) | 17.67 (14 156) | 18.18 (15 597) | 17.05 (14 628) | 17.97 (14 390) |
| Tertiary education | 44.14 (35 355) | 42.14 (36 150) | 43.73 (37 516) | 42.19 (33 790) |
| Missing, % (n) | 0.69 (549) | 0.73 (629) | 0.75 (646) | 0.84 (670) |
| Shift workers | 5.15 (4 126) | 5.38 (4 613) | 5.07 (4 348) | 5.17 (4 138) |
| Missing, % (n) | 0.28 (226) | 0.26 (226) | 0.26 (227) | 0.27 (218) |
| Employed | 57.23 (45 837) | 57.08 (48 963) | 57.11 (48 991) | 56.89 (45 561) |
| Missing, % (n) | 0.24 (192) | 0.22 (192) | 0.24 (202) | 0.24 (193) |
| Use of sleep medication(s) | 0.87 (697) | 1.06 (908) | 0.90 (773) | 1.07 (857) |
| Missing, % (n) | - | - | - | - |
| Suffering from depression | 11.39 (9 124) | 12.96 (11 115) | 11.37 (9 752) | 12.86 (10 302) |
| Missing, % (n) | - | - | - | - |
| Suffering from anxiety | 6.38 (5 106) | 6.99 (6 000) | 6.40 (5 488) | 7.02 (5 620) |
| Missing, % (n) | - | - | - | - |
| Suffering from chronic illness | 29.96 (23 994) | 32.86 (28 190) | 29.41 (25 226) | 32.37 (25 927) |
| Missing, % (n) | 1.98 (1 582) | 2.17 (1 860) | 1.91 (1 639) | 2.09 (1 674) |
| **Variables, mean (SD)** | | | | |
| Age, *years* | 56.91 (7.93) | 56.80 (7.97) | 56.95 (7.91) | 56.85 (7.93) |
| Missing, % (n) | - | - | - | - |
| TDI | -1.59 (2.92) | -1.51 (2.96) | -1.67 (2.87) | -1.56 (2.92) |
| Missing, % (n) | 0.10 (82) | 0.13 (109) | 0.14 (116) | 0.11 (90) |
| BMI, *kg/m^2^* | 27.23 (4.65) | 27.54 (4.82) | 27.26 (4.69) | 27.54 (4.83) |
| Missing, % (n) | 0.30 (239) | 0.30 (259) | 0.29 (251) | 0.32 (259) |
| SBP, *mmHg* | 138.30 (18.70) | 138.50 (18.72) | 138.20 (18.57) | 138.40 (18.54) |
| Missing, % (n) | 0.08 (66) | 0.10 (84) | 0.09 (75) | 0.08 (68) |
| Serum cholesterol, *mmol/L* | 5.75 (1.14) | 5.72 (1.14) | 5.75 (1.14) | 5.72 (1.14) |
| Missing, % (n) | 4.52 (3 623) | 4.46 (3 825) | 4.60 (3 942) | 4.62 (3 702) |
| Blood glucose, *mmol/L* | 5.11 (1.19) | 5.13 (1.22) | 5.10 (1.13) | 5.12 (1.23) |
| Missing, % (n) | 12.75 (10 212) | 12.56 (10 775) | 12.70 (10 892) | 12.87 (10 305) |

GRS, genetic risk score; SD, standard deviation; TDI, Townsend deprivation index; BMI, body mass index; and SBP, systolic blood pressure.

Table S9: Baseline characteristics of participants across groups categorized by dichotomizing to the median genetic risk scores for short sleep and chronotype (morning preference) in UK Biobank

|  | **UK Biobank (N = 306 292)** | | | |
| --- | --- | --- | --- | --- |
|  | **Both GRS**  **≤ median** | **Short sleep GRS**  **> median** | **Chronotype GRS**  **> median** | **Both GRS**  **> median** |
| **Total, % (n)** | 25.23 (77 263) | 24.77 (75 883) | 24.77 (75 883) | 25.23 (77 263) |
| **Variables, % (n)** | | | | |
| Male | 44.55 (34 423) | 44.94 (34 100) | 44.83 (34 017) | 45.11 (34 854) |
| Missing, % (n) | - | - | - | - |
| Married | 74.05 (57 213) | 73.70 (55 927) | 74.56 (56 578) | 74.55 (57 601) |
| Missing, % (n) | 0.48 (371) | 0.48 (363) | 0.46 (352) | 0.47 (363) |
| Weekly alcohol intake | 51.03 (39 427) | 50.81 (38 554) | 51.27 (38 908) | 51.02 (39 416) |
| Missing, % (n) | 0.06 (47) | 0.05 (35) | 0.04 (30) | 0.04 (34) |
| Current smokers | 10.12 (7 819) | 10.45 (7 927) | 9.83 (7 461) | 10.06 (7 769) |
| Missing, % (n) | 0.28 (213) | 0.31 (232) | 0.28 (213) | 0.30 (231) |
| Highly physically active | 33.27 (25 709) | 33.17 (25 174) | 34.20 (25 949) | 33.97 (26 248) |
| Missing, % (n) | 17.72 (13 690) | 17.84 (13 541) | 17.19 (13 044) | 17.49 (13 513) |
| Tertiary education | 44.20 (34 149) | 43.215 (32 743) | 44.30 (33 617) | 42.81 (33 074) |
| Missing, % (n) | 0.68 (522) | 0.71 (539) | 0.76 (580) | 0.77 (596) |
| Shift workers | 5.31 (4 100) | 5.43 (4 121) | 5.21 (3 953) | 5.30 (4 095) |
| Missing, % (n) | 0.23 (179) | 0.30 (227) | 0.25 (191) | 0.28 (215) |
| Employed | 59.50 (45 968) | 58.85 (44 657) | 59.23 (44 944) | 58.70 (45 355) |
| Missing, % (n) | 0.20 (153) | 0.25 (188) | 0.23 (175) | 0.24 (189) |
| Use of sleep medication(s) | 0.83 (641) | 0.97 (734) | 0.88 (671) | 0.95 (734) |
| Missing, % (n) | - | - | - | - |
| Suffering from depression | 11.35 (8 772) | 11.75 (8 915) | 11.28 (8 559) | 11.54 (8 913) |
| Missing, % (n) | - | - | - | - |
| Suffering from anxiety | 6.50 (5 023) | 6.43 (4 876) | 6.33 (4 803) | 6.61 (5 108) |
| Missing, % (n) | - | - | - | - |
| Suffering from chronic illness | 29.97 (23 153) | 30.80 (23 372) | 29.36 (22 278) | 30.31 (23 417) |
| Missing, % (n) | 2.05 (1 583) | 2.09 (1 589) | 1.98 (1 502) | 2.02 (1 563) |
| **Variables, mean (SD)** | | | | |
| Age, *years* | 56.64 (7.95) | 56.73 (7.93) | 56.73 (7.91) | 56.74 (7.92) |
| Missing, % (n) | - | - | - | - |
| TDI | -1.58 (2.91) | -1.55 (2.94) | -1.66 (2.88) | -1.62 (2.89) |
| Missing, % (n) | 0.11 (83) | 0.12 (94) | 0.11 (85) | 0.13 (103) |
| BMI, *kg/m^2^* | 27.30 (4.69) | 27.38 (4.73) | 27.28 (4.69) | 27.39 (4.74) |
| Missing, % (n) | 0.29 (227) | 0.28 (216) | 0.27 (203) | 0.31 (238) |
| SBP, *mmHg* | 138.20 (18.71) | 138.40 (18.66) | 138.10 (18.54) | 138.20 (18.47) |
| Missing, % (n) | 0.09 (70) | 0.08 (59) | 0.09 (69) | 0.07 (55) |
| Serum cholesterol, *mmol/L* | 5.74 (1.13) | 5.73 (1.14) | 5.74 (1.13) | 5.73 (1.13) |
| Missing, % (n) | 4.48 (3 465) | 4.48 (3 397) | 4.63 (3 516) | 4.52 (3 491) |
| Blood glucose, *mmol/L* | 5.10 (1.18) | 5.11 (1.18) | 5.10 (1.15) | 5.10 (1.15) |
| Missing, % (n) | 12.71 (9 817) | 12.63 (9 583) | 12.74 (9 669) | 12.79 (9 879) |

GRS, genetic risk score; SD, standard deviation; TDI, Townsend deprivation index; BMI, body mass index; and SBP, systolic blood pressure.

Table S10: Baseline characteristics of participants across groups categorized by dichotomizing to the median genetic risk scores for long sleep and chronotype (morning preference) in UK Biobank

|  | **UK Biobank (N = 253 111)** | | | |
| --- | --- | --- | --- | --- |
|  | **Both GRS**  **≤ median** | **Long sleep GRS**  **> median** | **Chronotype GRS**  **> median** | **Both GRS**  **> median** |
| **Total, % (n)** | 28.41 (71 916) | 21.59 (54 640) | 30.04 (76 031) | 19.96 (50 524) |
| **Variables, % (n)** | | | | |
| Male | 44.56 (32 044) | 43.94 (24 007) | 44.76 (34 030) | 44.26 (22 361) |
| Missing, % (n) | - | - | - | - |
| Married | 75.49 (54 289) | 75.43 (41 216) | 76.31 (58 023) | 75.84 (38 318) |
| Missing, % (n) | 0.41 (296) | 0.45 (246) | 0.42 (318) | 0.41 (209) |
| Weekly alcohol intake | 51.11 (36 754) | 50.94 (27 834) | 51.41 (39 087) | 51.26 (25 899) |
| Missing, % (n) | 0.04 (28) | 0.06 (33) | 0.03 (26) | 0.04 (22) |
| Current smokers | 9.59 (6 895) | 9.81 (5 361) | 9.32 (7 089) | 9.49 (4 793) |
| Missing, % (n) | 0.28 (203) | 0.26 (143) | 0.29 (217) | 0.24 (123) |
| Highly physically active | 33.19 (23 868) | 33.04 (18 055) | 34.13 (25 946) | 33.61 (16 980) |
| Missing, % (n) | 17.29 (12 436) | 17.53 (9 581) | 16.84 (12 800) | 16.97 (8 574) |
| Tertiary education | 44.47 (31 981) | 44.10 (24 095) | 44.31 (33 689) | 44.15 (22 304) |
| Missing, % (n) | 0.69 (497) | 0.63 (342) | 0.72 (546) | 0.70 (355) |
| Shift workers | 4.66 (3 350) | 4.53 (2 473) | 4.47 (3 395) | 4.40 (2 221) |
| Missing, % (n) | 0.28 (198) | 0.27 (145) | 0.26 (195) | 0.25 (127) |
| Employed | 56.43 (40 582) | 56.09 (30 645) | 56.24 (42 763) | 56.23 (28 409) |
| Missing, % (n) | 0.23 (163) | 0.23 (125) | 0.23 (175) | 0.22 (109) |
| Use of sleep medication(s) | 0.75 (540) | 0.79 (430) | 0.74 (564) | 0.83 (418) |
| Missing, % (n) | - | - | - | - |
| Suffering from depression | 11.66 (8 386) | 11.39 (6 226) | 11.49 (8 739) | 11.26 (5 688) |
| Missing, % (n) | - | - | - | - |
| Suffering from anxiety | 6.40 (4 604) | 6.31 (3 448) | 6.18 (4 699) | 6.50 (3 286) |
| Missing, % (n) | - | - | - | - |
| Suffering from chronic illness | 30.15 (21 683) | 29.74 (16 248) | 29.39 (22 343) | 29.29 (14 796) |
| Missing, % (n) | 1.90 (1 363) | 2.03 (1 110) | 1.78 (1 350) | 1.89 (956) |
| **Variables, mean (SD)** | | | | |
| Age, *years* | 56.92 (7.99) | 56.87 (8.05) | 56.91 (7.99) | 56.90 (8.00) |
| Missing, % (n) | - | - | - | - |
| TDI | -1.67 (2.87) | -1.66 (2.88) | -1.73 (2.82) | -1.74 (2.84) |
| Missing, % (n) | 0.13 (91) | 0.10 (54) | 0.11 (87) | 0.14 (69) |
| BMI, *kg/m^2^* | 27.30 (4.67) | 27.12 (4.55) | 27.31 (4.69) | 27.15 (4.60) |
| Missing, % (n) | 0.29 (205) | 0.28 (153) | 0.31 (234) | 0.28 (141) |
| SBP, *mmHg* | 138.40 (18.83) | 138.40 (18.77) | 138.30 (18.61) | 138.20 (18.72) |
| Missing, % (n) | 0.09 (62) | 0.08 (45) | 0.07 (55) | 0.10 (48) |
| Serum cholesterol, *mmol/L* | 5.73 (1.14) | 5.75 (1.14) | 5.73 (1.14) | 5.74 (1.14) |
| Missing, % (n) | 4.45 (3 203) | 4.55 (2 484) | 4.57 (3 472) | 4.71 (2 379) |
| Blood glucose, *mmol/L* | 5.12 (1.20) | 5.11 (1.18) | 5.10 (1.16) | 5.09 (1.13) |
| Missing, % (n) | 12.52 (9 001) | 12.71 (6 943) | 12.73 (9 675) | 13.10 (6 620) |

GRS, genetic risk score; SD, standard deviation; TDI, Townsend deprivation index; BMI, body mass index; and SBP, systolic blood pressure.

Table S11: Statistical test of the proportional hazard assumption for observational and one-sample Mendelian randomization (MR) Cox regression models

|  | **Observational analysis ^*^** | | | | |  | **One-sample MR analysis** | | | | | | | |
| --- | --- | --- | --- | --- | --- | --- | --- | --- | --- | --- | --- | --- | --- | --- |
| **Sleep trait** | **UK Biobank** | |  | **HUNT2** | |  | **UK Biobank ^†^** | |  | | **HUNT2 ^‡^** | | | |
|  | **Correlation coefficient ^#^** | **P** |  | **Correlation coefficient ^#^** | **P** |  | **Correlation coefficient ^#^** | **P** | |  | **Correlation coefficient ^#^** | | **P** | |
| Insomnia symptoms | 1.7693 | 0.180 |  | 0.2823 | 0.595 |  | -0.0153 | 0.316 | |  | | -0.0048 | | 0.702 |
| 24-hour sleep duration (h) | 0.0626 | 0.801 |  | 0.0030 | 0.957 |  | 0.0158 | 0.287 | |  | | -0.0081 | | 0.516 |
| Short sleep  (≤6 h vs. 7-8 h) | 0.0506 | 0.820 |  | 0.2945 | 0.587 |  | -0.0049 | 0.515 | |  | | 0.0134 | | 0.385 |
| Long sleep  (≥9 h vs. 7-8 h) | 0.5543 | 0.460 |  | 0.0001 | 0.992 |  | 0.0067 | 0.406 | |  | | -0.0021 | | 0.871 |
| Chronotype  (morning preference) | 0.62754 | 0.430 |  | - | - |  | 0.0071 | 0.313 | |  | | - | | - |

^*^ Observational analysis adjusted for covariates in the main model.

**^†^** Using unweighted genetic risk score for each sleep trait in the MR Cox regression model, with adjustment for age, sex, assessment center, 40 genetic principal components, and genotyping chip.

**^‡^** Using weighted genetic risk score for each sleep trait in the MR Cox regression model, with adjustment for age, sex, 20 genetic principal components, and genotyping chip.

^#^ Values represent the Pearson’s correlation coefficient between the first scaled Schoenfeld residual in the Cox regression and the rank-normalized natural logarithm of follow-up time.

Table S12: Statistical test of the proportional hazard assumption for observational and 2x2 factorial Mendelian randomization (MR) Cox regression models

|  | **Observational analysis ^*^** | | | | |  | | **One-sample MR analysis** | | | | | | |
| --- | --- | --- | --- | --- | --- | --- | --- | --- | --- | --- | --- | --- | --- | --- |
| **Sleep trait combination** | **UK Biobank** | |  | **HUNT2** | | |  | **UK Biobank ^†^** | |  | | **HUNT2 ^‡^** | | |
|  | **Correlation coefficient ^#^** | **P** |  | **Correlation coefficient ^#^** | **P** | |  | **Correlation coefficient ^#^** | **P** | |  | | **Correlation coefficient ^#^** | **P** |
| Insomnia symptoms – Short sleep | 2.6636 | 0.450 |  | 4.750 | 0.191 | |  | -0.0103 | 0.170 | |  | | -0.0161 | 0.292 |
| Insomnia symptoms – Long sleep | 2.3939 | 0.490 |  | 0.557 | 0.906 | |  | -0.0028 | 0.736 | |  | | 0.0016 | 0.904 |
| Insomnia symptoms – Chronotype | 2.4518 | 0.480 |  | - | - | |  | -0.0020 | 0.776 | |  | | - | - |
| Short sleep – Chronotype | 2.4920 | 0.480 |  | - | - | |  | -0.0049 | 0.517 | |  | | - | - |
| Long sleep – Chronotype | 1.6624 | 0.650 |  | - | - | |  | 0.0075 | 0.362 | |  | | - | - |

^*^ Observational analysis adjusted for covariates in the main model.

**^†^** Using unweighted genetic risk score for the sleep traits in the factorial MR Cox regression model, with adjustment for age, sex, assessment center, 40 genetic principal components, and genotyping chip.

**^‡^** Using weighted genetic risk score for each sleep trait in the factorial MR Cox regression model, with adjustment for age, sex, 20 genetic principal components, and genotyping chip.

^#^ Values represent the Pearson’s correlation coefficient between the first scaled Schoenfeld residual in the factorial MR Cox regression and the rank-normalized natural logarithm of follow-up time.

Table S13: Hazard ratios (95% confidence intervals) from observational Cox regression analysis for incident atrial fibrillation (AF) in relation to individual self-reported sleep traits (insomnia symptoms, short sleep, long sleep and chronotype) in UK Biobank and HUNT2

|  |  | **No insomnia symptoms** | **Insomnia symptoms** |
| --- | --- | --- | --- |
| **UK Biobank**  (n = 287 352) | AF events/Person-years | 11 085/2 420 927 | 5 107/912 170 |
|  | Crude model | Reference | 1.20 (1.16, 1.24) |
|  | Main model | Reference | 1.15 (1.06, 1.24) |
|  | Additional model | Reference | 1.10 (1.06, 1.13) |
| **HUNT2**  (n = 31 458) | AF events/Person-years | 3 007/575 945 | 620/76 159 |
|  | Crude model | Reference | 1.13 (1.03, 1.24) |
|  | Main model | Reference | 1.10 (1.00, 1.21) |
|  | Additional model | Reference | 1.08 (0.98, 1.19) |
|  |  | **Normal sleep duration** | **Short sleep duration** |
| **UK Biobank**  (n = 265 945) | AF events/Person-years | 10 675/2 316 522 | 3 858/774 470 |
|  | Crude model | Reference | 1.11 (1.07, 1.15) |
|  | Main model | Reference | 1.01 (0.93, 1.11) |
|  | Additional model | Reference | 1.02 (0.99, 1.06) |
| **HUNT2**  (n = 23 932) | AF events/Person-years | 2 288/469 729 | 199/41 411 |
|  | Crude model | Reference | 1.16 (0.99, 1.36) |
|  | Main model | Reference | 0.96 (0.53, 1.74) |
|  | Additional model | Reference | 0.94 (0.51, 1.72) |
|  |  | **Normal sleep duration** | **Long sleep duration** |
| **UK Biobank**  (n = 220 385) | AF events/Person-years | 10 675/2 316 522 | 1 659/242 106 |
|  | Crude model | Reference | 1.21 (1.15, 1.27) |
|  | Main model | Reference | 1.01 (0.88, 1.15) |
|  | Additional model | Reference | 1.06 (1.01, 1.12) |
| **HUNT2**  (n = 29 514) | AF events/Person-years | 2 288/469 729 | 1 140/140 964 |
|  | Crude model | Reference | 1.09 (1.01, 1.18) |
|  | Main model | Reference | 1.08 (0.99, 1.16) |
|  | Additional model | Reference | 1.07 (0.99, 1.16) |
|  |  | **Evening chronotype** | **Morning chronotype** |
| **UK Biobank**  (n = 287 352) | AF events/Person-years | 5 745/1 240 830 | 10 447/2 092 267 |
|  | Crude model | Reference | 0.96 (0.93, 0.99) |
|  | Main model | Reference | 0.95 (0.88, 1.02) |
|  | Additional model | Reference | 1.01 (0.98, 1.04) |

Crude model adjusted for age and sex.

Main model adjusted for age, sex, marital status, alcohol intake frequency, smoking status, body mass index, physical activity, education, Townsend deprivation index (for UK Biobank only), shift work and employment status.

Additional model adjusted for covariates in the main model plus systolic blood pressure, serum cholesterol level, blood glucose level, time since last meal, use of sleep medication(s), depression and anxiety.

Table S14: Hazard ratios (95% confidence intervals) from observational Cox regression analysis for incident atrial fibrillation (AF) in relation to 24-hour sleep duration in UK Biobank and HUNT2

|  |  | **24-hour sleep duration** |
| --- | --- | --- |
| **UK Biobank**  (n = 287 352) | AF events/Person-years | 16 192/3 333 097 |
|  | Crude model | 1.00 (0.99, 1.01) |
|  | Main model | 0.99 (0.96, 1.03) |
|  | Additional model | 1.00 (0.98, 1.01) |
| **HUNT2**  (n = 31 458) | AF events/Person-years | 3 627/652 104 |
|  | Crude model | 0.99 (0.96, 1.02) |
|  | Main model | 0.99 (0.96, 1.02) |
|  | Additional model | 0.99 (0.96, 1.02) |

Crude model adjusted for age and sex.

Main model adjusted for age, sex, marital status, alcohol intake frequency, smoking status, body mass index, physical activity, education, Townsend deprivation index (for UK Biobank only), shift work and employment status.

Additional model adjusted for covariates in the main model plus systolic blood pressure, serum cholesterol level, blood glucose level, time since last meal, use of sleep medication(s), depression and anxiety.

Table S15: Hazard ratios (95% confidence intervals) from observational Cox regression analysis for incident atrial fibrillation (AF) in relation to combination of insomnia symptoms and short sleep duration in UK Biobank and HUNT2

|  |  | **Normal sleep duration** | | |  | **Short sleep duration** | |
| --- | --- | --- | --- | --- | --- | --- | --- |
|  |  | **Insomnia symptoms** | | | ___ | **Insomnia symptoms** | |
|  |  | **No** | **Yes** | |  | **No** | **Yes** |
| **UK Biobank**  (n = 265 945) | AF events/Person-years | 8 113/1 844 350 | 2 562/472 172 | |  | 1 748/384 679 | 2 110/389 790 |
|  | Crude model | Reference | 1.16 (1.11, 1.22) | |  | 1.04 (0.99, 1.09) | 1.25 (1.19, 1.31) |
|  | Main model | Reference | 1.14 (1.03, 1.26) | |  | 0.98 (0.86, 1.11) | 1.10 (0.98, 1.25) |
|  | Additional model | Reference | 1.08 (1.03, 1.13) | |  | 0.98 (0.93, 1.03) | 1.10 (1.05, 1.16) |
| **HUNT2**  (n = 23 932) | AF events/Person-years | 1 927/420 370 | 361/49 359 | |  | 139/33 123 | 60/8 288 |
|  | Crude model | Reference | 1.16 (1.02, 1.31) | |  | 1.15 (0.96, 1.38) | 1.28 (0.96, 1.70) |
|  | Main model | Reference | 1.23 (0.73, 2.08) | |  | 0.96 (0.48, 1.92) | 1.09 (0.34, 3.51) |
|  | Additional model | Reference | 1.21 (0.68, 2.17) | |  | 0.94 (0.46, 1.91) | 1.05 (0.31, 3.55) |

Crude model adjusted for age and sex.

Main model adjusted for age, sex, marital status, alcohol intake frequency, smoking status, body mass index, physical activity, education, Townsend deprivation index (for UK Biobank only), shift work and employment status.

Additional model adjusted for covariates in the main model plus systolic blood pressure, serum cholesterol level, blood glucose level, time since last meal, use of sleep medication(s), depression and anxiety.

Table S16: Hazard ratios (95% confidence intervals) from observational Cox regression analysis for incident atrial fibrillation (AF) in relation to combination of insomnia symptoms and long sleep duration in UK Biobank and HUNT2

|  |  | **Normal sleep duration** | | |  | **Long sleep duration** | |
| --- | --- | --- | --- | --- | --- | --- | --- |
|  |  | **Insomnia symptoms** | | | ___ | **Insomnia symptoms** | |
|  |  | **No** | **Yes** | |  | **No** | **Yes** |
| **UK Biobank**  (n = 220 385) | AF events/Person-years | 8 113/1 844 350 | 2 562/472 172 | |  | 1 224/191 898 | 435/50 208 |
|  | Crude model | Reference | 1.16 (1.11, 1.22) | |  | 1.17 (1.10, 1.24) | 1.55 (1.40, 1.70) |
|  | Main model | Reference | 1.11 (1.00, 1.24) | |  | 0.96 (0.82, 1.11) | 1.36 (1.04, 1.79) |
|  | Additional model | Reference | 1.08 (1.03, 1.13) | |  | 1.04 (0.98, 1.11) | 1.22 (1.10, 1.34) |
| **HUNT2**  (n = 29 514) | AF events/Person-years | 1 927/420 370 | 361/49 359 | |  | 941/ 122 452 | 199/18 512 |
|  | Crude model | Reference | 1.14 (1.01, 1.28) | |  | 1.10 (1.00, 1.20) | 1.17 (1.00, 1.37) |
|  | Main model | Reference | 1.11 (0.98, 1.24) | |  | 1.09 (1.00, 1.19) | 1.12 (0.96, 1.31) |
|  | Additional model | Reference | 1.09 (0.97, 1.24) | |  | 1.08 (1.00, 1.19) | 1.09 (0.92, 1.28) |

Crude model adjusted for age and sex.

Main model adjusted for age, sex, marital status, alcohol intake frequency, smoking status, body mass index, physical activity, education, Townsend deprivation index (for UK Biobank only), shift work and employment status.

Additional model adjusted for covariates in the main model plus systolic blood pressure, serum cholesterol level, blood glucose level, time since last meal, use of sleep medication(s), depression and anxiety.

Table S17: Hazard ratios (95% confidence intervals) for atrial fibrillation (AF) according to the observational joint association of self-reported insomnia symptoms and chronotype in UK Biobank

|  |  | **Evening chronotype** | | |  | **Morning chronotype** | |
| --- | --- | --- | --- | --- | --- | --- | --- |
|  |  | **Insomnia symptoms** | | | ___ | **Insomnia symptoms** | |
|  |  | **No** | **Yes** | |  | **No** | **Yes** |
| **UK Biobank**  (n = 287 352) | AF events/Person-years | 3 910/901 427 | 1 835/339 402 | |  | 7 175/1 519 500 | 3 272/572 767 |
|  | Crude model | Reference | 1.24 (1.17, 1.31) | |  | 0.98 (0.94, 1.02) | 1.15 (1.10, 1.21) |
|  | Main model | Reference | 1.18 (1.04, 1.35) | |  | 0.96 (0.88, 1.05) | 1.08 (0.97, 1.21) |
|  | Additional model | Reference | 1.11 (1.05, 1.18) | |  | 1.02 (0.98, 1.06) | 1.10 (1.05, 1.16) |

Crude model adjusted for age and sex.

Main model adjusted for age, sex, marital status, alcohol intake frequency, smoking status, body mass index, physical activity, education, Townsend deprivation index, shift work and employment status.

Additional model adjusted for covariates in the main model plus systolic blood pressure, serum cholesterol level, blood glucose level, time since last meal, use of sleep medication(s), depression and anxiety.

Table S18: Hazard ratios (95% confidence intervals) for atrial fibrillation (AF) according to the observational joint association of self-reported short sleep duration and chronotype in UK Biobank

|  |  | **Evening chronotype** | | |  | **Morning chronotype** | |
| --- | --- | --- | --- | --- | --- | --- | --- |
|  |  | **Sleep duration** | | | ___ | **Sleep duration** | |
|  |  | **Normal** | **Short** | |  | **Normal** | **Short** |
| **UK Biobank**  (n =265 945) | AF events/Person-years | 3 761/858 963 | 1 285/279 864 | |  | 6 914/1 457 559 | 2 573/494 606 |
|  | Crude model | Reference | 1.12 (1.06, 1.20) | |  | 0.97 (0.94, 1.01) | 1.07 (1.02, 1.12) |
|  | Main model | Reference | 1.04 (0.89, 1.22) | |  | 0.96 (0.87, 1.05) | 0.96 (0.85, 1.09) |
|  | Additional model | Reference | 1.03 (0.97, 1.10) | |  | 1.01 (0.97, 1.06) | 1.03 (0.98, 1.09) |

Crude model adjusted for age and sex.

Main model adjusted for age, sex, marital status, alcohol intake frequency, smoking status, body mass index, physical activity, education, Townsend deprivation index, shift work and employment status.

Additional model adjusted for covariates in the main model plus systolic blood pressure, serum cholesterol level, blood glucose level, time since last meal, use of sleep medication(s), depression and anxiety.

Table S19: Hazard ratios (95% confidence intervals) for atrial fibrillation (AF) according to the observational joint association of self-reported long sleep duration and chronotype in UK Biobank

|  |  | **Evening chronotype** | | |  | **Morning chronotype** | |
| --- | --- | --- | --- | --- | --- | --- | --- |
|  |  | **Sleep duration** | | | ___ | **Sleep duration** | |
|  |  | **Normal** | **Long** | |  | **Normal** | **Long** |
| **UK Biobank**  (n =220 385) | AF events/Person-years | 3 761/858 963 | 699/102 003 | |  | 6 914/1 457 559 | 960/140 103 |
|  | Crude model | Reference | 1.24 (1.14, 1.34) | |  | 0.97 (0.94, 1.01) | 1.15 (1.07, 1.24) |
|  | Main model | Reference | 1.11 (0.90, 1.37) | |  | 0.99 (0.90, 1.09) | 0.93 (0.77, 1.11) |
|  | Additional model | Reference | 1.07 (0.99, 1.16) | |  | 1.02 (0.98, 1.06) | 1.07 (1.00, 1.15) |

Crude model adjusted for age and sex.

Main model adjusted for age, sex, marital status, alcohol intake frequency, smoking status, body mass index, physical activity, education, Townsend deprivation index, shift work and employment status.

Additional model adjusted for covariates in the main model plus systolic blood pressure, serum cholesterol level, blood glucose level, time since last meal, use of sleep medication(s), depression and anxiety.

Table S20: One-sample Mendelian randomization Cox regression analysis for risk of incident atrial fibrillation associated with sleep traits in HUNT2 using weighted and unweighted genetic risk scores for sleep traits

| **Sleep trait** | **Weighted genetic risk score** | |  | **Unweighted genetic risk score** | |
| --- | --- | --- | --- | --- | --- |
|  | **N**  **(incident cases)** | **Hazard ratio**  **(95% CI)*** |  | **N**  **(incident cases)** | **Hazard ratio**  **(95% CI)*** |
| Insomnia symptoms ^#^ | 45 322  (6 296) | 0.95  (0.81, 1.11) |  | 45 322  (6 296) | 0.98  (0.83, 1.15) |
| 24-hour sleep duration (h) | 45 322  (6 296) | 1.06  (0.52, 2.15) |  | 45 322  (6 296) | 0.97  (0.49, 1.90) |
| Short sleep ^#^  (≤6 h vs. 7-8 h) | 33 588  (4 198) | 1.41  (0.57, 3.46) |  | 33 588  (4 198) | 1.97  (0.68, 5.72) |
| Long sleep ^#^  (≥9 h vs. 7-8 h) | 42 506  (5 932) | 1.70  (0.68, 4.25) |  | 42 506  (5 932) | 1.19  (0.64, 2.23) |

CI, confidence interval

**^*^** Adjusted for age, sex, 20 genetic principal components, and genotyping chip.

**^#^** Hazard ratio (95% CI) scaled to per doubling in odds of the sleep trait.

Table S21: Associations between genetic risk scores and potential confounders in UK Biobank

| **Instrument** | **Confounder** | **Coefficient (Beta)** | **SE** | **N** | **P** |  |
| --- | --- | --- | --- | --- | --- | --- |
| Insomnia uwGRS | Marital status | 0.064 | 0.051 | 230 006 | 0.212 |  |
|  | Alcohol intake | -0.078 | 0.023 | 230 006 | 7.22e-04 |  |
|  | Smoking status | 0.283 | 0.033 | 230 006 | <2e-16 | * |
|  | BMI | 0.055 | 0.005 | 230 006 | <2e-16 | * |
|  | Physical activity | -0.002 | 0.030 | 230 006 | 0.935 |  |
|  | TDI | 0.030 | 0.008 | 230 006 | 9.55e-05 | * |
|  | Education | -0.212 | 0.023 | 230 006 | <2e-16 | * |
|  | Shift work | 0.052 | 0.102 | 230 006 | 0.612 |  |
|  | Employment status | 0.141 | 0.046 | 230 006 | 0.002 |  |
|  | SBP | 8.59e-04 | 0.001 | 230 006 | 0.481 |  |
|  | Fasting time | -0.003 | 0.009 | 230 006 | 0.739 |  |
|  | Serum cholesterol | -0.054 | 0.019 | 230 006 | 0.005 |  |
|  | Blood glucose | 0.023 | 0.019 | 230 006 | 0.213 |  |
|  | Depression | 0.622 | 0.071 | 230 006 | <2e-16 | * |
|  | Anxiety | 0.294 | 0.092 | 230 006 | 0.001 |  |
|  | Sleep medication | 0.409 | 0.232 | 230 006 | 0.078 |  |
|  | Chronic illness | 0.679 | 0.049 | 230 006 | <2e-16 | * |
| Sleep duration uwGRS | Marital status | -0.010 | 0.027 | 230 006 | 0.715 |  |
|  | Alcohol intake | 0.001 | 0.012 | 230 006 | 0.909 |  |
|  | Smoking status | -0.010 | 0.017 | 230 006 | 0.555 |  |
|  | BMI | -0.016 | 0.003 | 230 006 | 4.58e-10 | * |
|  | Physical activity | -0.008 | 0.016 | 230 006 | 0.588 |  |
|  | TDI | -0.004 | 0.004 | 230 006 | 0.325 |  |
|  | Education | 0.045 | 0.012 | 230 006 | 3.02e-04 | * |
|  | Shift work | 0.003 | 0.054 | 230 006 | 0.958 |  |
|  | Employment status | -0.066 | 0.024 | 230 006 | 0.007 |  |
|  | SBP | 0.001 | 0.001 | 230 006 | 0.020 |  |
|  | Fasting time | 6.86e-04 | 0.005 | 230 006 | 0.888 |  |
|  | Serum cholesterol | -0.003 | 0.010 | 230 006 | 0.786 |  |
|  | Blood glucose | 0.001 | 0.010 | 230 006 | 0.914 |  |
|  | Depression | 0.067 | 0.038 | 230 006 | 0.073 |  |
|  | Anxiety | 0.026 | 0.049 | 230 006 | 0.595 |  |
|  | Sleep medication | -0.215 | 0.123 | 230 006 | 0.080 |  |
|  | Chronic illness | -0.067 | 0.026 | 230 006 | 0.010 |  |
| Short sleep uwGRS | Marital status | 0.002 | 0.016 | 212 880 | 0.882 |  |
|  | Alcohol intake | -0.028 | 0.007 | 212 880 | 2.03e-04 | * |
|  | Smoking status | 0.015 | 0.011 | 212 880 | 0.161 |  |
|  | BMI | 0.006 | 0.002 | 212 880 | 2.26e-04 | * |
|  | Physical activity | -0.007 | 0.009 | 212 880 | 0.436 |  |
|  | TDI | 0.002 | 0.002 | 212 880 | 0.442 |  |
|  | Education | -0.060 | 0.007 | 212 880 | 6.55e-16 | * |
|  | Shift work | 4.25e-04 | 0.032 | 212 880 | 0.989 |  |
|  | Employment status | -0.001 | 0.015 | 212 880 | 0.962 |  |
|  | SBP | 0.001 | 3.90e-04 | 212 880 | 0.137 |  |
|  | Fasting time | 0.001 | 0.003 | 212 880 | 0.821 |  |
|  | Serum cholesterol | -0.009 | 0.006 | 212 880 | 0.138 |  |
|  | Blood glucose | -0.008 | 0.006 | 212 880 | 0.204 |  |
|  | Depression | 0.038 | 0.023 | 212 880 | 0.098 |  |
|  | Anxiety | 0.004 | 0.029 | 212 880 | 0.895 |  |
|  | Sleep medication | 0.103 | 0.077 | 212 880 | 0.179 |  |
|  | Chronic illness | 0.042 | 0.016 | 212 880 | 0.008 |  |
| Long sleep uwGRS | Marital status | -0.021 | 0.008 | 177 072 | 0.012 |  |
|  | Alcohol intake | 0.003 | 0.004 | 177 072 | 0.434 |  |
|  | Smoking status | 0.002 | 0.005 | 177 072 | 0.724 |  |
|  | BMI | -0.007 | 7.89e-04 | 177 072 | <2e-16 | * |
|  | Physical activity | -0.019 | 0.005 | 177 072 | 4.53e-05 | * |
|  | TDI | 5.36e-04 | 0.001 | 177 072 | 0.671 |  |
|  | Education | -0.005 | 0.004 | 177 072 | 0.181 |  |
|  | Shift work | -0.007 | 0.017 | 177 072 | 0.697 |  |
|  | Employment status | -0.004 | 0.007 | 177 072 | 0.536 |  |
|  | SBP | 2.28e-04 | 1.92e-04 | 177 072 | 0.235 |  |
|  | Fasting time | 0.002 | 0.001 | 177 072 | 0.136 |  |
|  | Serum cholesterol | 0.009 | 0.003 | 177 072 | 0.005 |  |
|  | Blood glucose | 9.79e-04 | 0.003 | 177 072 | 0.742 |  |
|  | Depression | -0.015 | 0.011 | 177 072 | 0.197 |  |
|  | Anxiety | 0.029 | 0.015 | 177 072 | 0.050 |  |
|  | Sleep medication | 0.021 | 0.041 | 177 072 | 0.600 |  |
|  | Chronic illness | -0.002 | 0.008 | 177 072 | 0.765 |  |
| Chronotype (morning preference) uwGRS | Marital status | 0.135 | 0.058 | 230 006 | 0.021 |  |
|  | Alcohol intake | -0.056 | 0.026 | 230 006 | 0.033 |  |
|  | Smoking status | -0.047 | 0.037 | 230 006 | 0.211 |  |
|  | BMI | 0.013 | 0.006 | 230 006 | 0.017 |  |
|  | Physical activity | 0.198 | 0.034 | 230 006 | 3.87e-09 | * |
|  | TDI | -0.049 | 0.009 | 230 006 | 4.44e-08 | * |
|  | Education | -0.066 | 0.027 | 230 006 | 0.012 |  |
|  | Shift work | -0.098 | 0.115 | 230 006 | 0.398 |  |
|  | Employment status | -0.029 | 0.052 | 230 006 | 0.575 |  |
|  | SBP | -0.001 | 0.001 | 230 006 | 0.309 |  |
|  | Fasting time | -0.018 | 0.010 | 230 006 | 0.078 |  |
|  | Serum cholesterol | -0.047 | 0.022 | 230 006 | 0.033 |  |
|  | Blood glucose | -0.066 | 0.021 | 230 006 | 0.002 |  |
|  | Depression | -0.024 | 0.081 | 230 006 | 0.767 |  |
|  | Anxiety | 0.059 | 0.105 | 230 006 | 0.573 |  |
|  | Sleep medication | -0.032 | 0.263 | 230 006 | 0.903 |  |
|  | Chronic illness | -0.207 | 0.056 | 230 006 | 2.12e-04 | * |

uwGRS, unweighted genetic risk score; SE, standard error; BMI, body mass index; TDI, Townsend deprivation index; SBP, systolic blood pressure.

Coefficients are in terms of an average-SNP increase in the allele score per unit/level increase in confounder.

* Associations surpassing multiple-testing corrected p-value threshold of 0.05/85 = 5.88e-04.

Table S22: Associations between genetic risk scores and potential confounders in HUNT2

| **Instrument** | **Confounder** | **Coefficient (Beta)** | **SE** | **N** | **P** |  |
| --- | --- | --- | --- | --- | --- | --- |
| Insomnia wGRS | Marital status | 0.005 | 0.005 | 27 631 | 0.293 |  |
|  | Alcohol intake | -3.38e-04 | 0.004 | 27 631 | 0.927 |  |
|  | Smoking status | 0.015 | 0.003 | 27 631 | 9.17e-06 | * |
|  | BMI | 0.003 | 0.001 | 27 631 | 4.22e-05 | * |
|  | Physical activity | -0.007 | 0.003 | 27 631 | 0.030 |  |
|  | Education | -4.83e-04 | 0.004 | 27 631 | 0.909 |  |
|  | Shift work | 0.008 | 0.007 | 27 631 | 0.286 |  |
|  | Employment status | 0.003 | 0.007 | 27 631 | 0.663 |  |
|  | SBP | -1.70e-04 | 1.60e-04 | 27 631 | 0.287 |  |
|  | Fasting time | 0.002 | 0.001 | 27 631 | 0.178 |  |
|  | Serum cholesterol | -0.010 | 0.002 | 27 631 | 3.91e-05 | * |
|  | Blood glucose | -6.56e-04 | 0.002 | 27 631 | 0.758 |  |
|  | HADS - Depression | -0.003 | 0.001 | 27 631 | 0.005 |  |
|  | HADS - Anxiety | 0.004 | 0.001 | 27 631 | 2.33e-04 | * |
|  | Sleep medication | 0.035 | 0.012 | 27 631 | 0.006 |  |
|  | Chronic illness | 0.011 | 0.007 | 27 631 | 0.109 |  |
| Sleep duration wGRS | Marital status | -0.992 | 3.371 | 27 631 | 0.769 |  |
|  | Alcohol intake | 3.772 | 2.598 | 27 631 | 0.146 |  |
|  | Smoking status | -3.423 | 2.389 | 27 631 | 0.152 |  |
|  | BMI | -0.746 | 0.513 | 27 631 | 0.145 |  |
|  | Physical activity | 0.313 | 2.338 | 27 631 | 0.893 |  |
|  | Education | -2.877 | 2.951 | 27 631 | 0.330 |  |
|  | Shift work | -3.724 | 5.135 | 27 631 | 0.468 |  |
|  | Employment status | 2.505 | 4.883 | 27 631 | 0.608 |  |
|  | SBP | -0.042 | 0.112 | 27 631 | 0.707 |  |
|  | Fasting time | -0.802 | 1.017 | 27 631 | 0.431 |  |
|  | Serum cholesterol | -1.109 | 1.751 | 27 631 | 0.527 |  |
|  | Blood glucose | 0.154 | 1.490 | 27 631 | 0.918 |  |
|  | HADS - Depression | -0.198 | 0.831 | 27 631 | 0.812 |  |
|  | HADS - Anxiety | 0.095 | 0.734 | 27 631 | 0.897 |  |
|  | Sleep medication | 3.179 | 8.787 | 27 631 | 0.718 |  |
|  | Chronic illness | -1.720 | 4.652 | 27 631 | 0.712 |  |
| Short sleep wGRS | Marital status | 5.05e-04 | 0.001 | 21 211 | 0.697 |  |
|  | Alcohol intake | -3.24e-04 | 0.001 | 21 211 | 0.745 |  |
|  | Smoking status | 0.001 | 0.001 | 21 211 | 0.098 |  |
|  | BMI | 5.03e-04 | 2.00e-04 | 21 211 | 0.012 |  |
|  | Physical activity | -0.001 | 20.001 | 21 211 | 0.200 |  |
|  | Education | -1.04e-04 | 0.001 | 21 211 | 0.927 |  |
|  | Shift work | -0.001 | 0.002 | 21 211 | 0.437 |  |
|  | Employment status | -0.003 | 0.002 | 21 211 | 0.124 |  |
|  | SBP | 1.75e-05 | 4.44e-05 | 21 211 | 0.694 |  |
|  | Fasting time | 5.14e-04 | 3.87e-04 | 21 211 | 0.184 |  |
|  | Serum cholesterol | -0.002 | 6.80e-04 | 21 211 | 0.014 |  |
|  | Blood glucose | 2.43e-04 | 6.33e-04 | 21 211 | 0.701 |  |
|  | HADS - Depression | -3.27e-04 | 3.26e-04 | 21 211 | 0.315 |  |
|  | HADS - Anxiety | -1.66e-04 | 2.84e-04 | 21 211 | 0.559 |  |
|  | Sleep medication | -1.23e-04 | 0.004 | 21 211 | 0.974 |  |
|  | Chronic illness | 6.30e-04 | 0.002 | 21 211 | 0.730 |  |
| Long sleep wGRS | Marital status | -7.73e-04 | 0.001 | 25 942 | 0.455 |  |
|  | Alcohol intake | 5.01e-04 | 7.94e-04 | 25 942 | 0.528 |  |
|  | Smoking status | 2.14e-04 | 7.33e-04 | 25 942 | 0.770 |  |
|  | BMI | -3.93e-04 | 1.57e-04 | 25 942 | 0.012 |  |
|  | Physical activity | -5.69e-04 | 7.17e-04 | 25 942 | 0.427 |  |
|  | Education | 0.002 | 9.01e-04 | 25 942 | 0.072 |  |
|  | Shift work | 1.97e-05 | 0.002 | 25 942 | 0.901 |  |
|  | Employment status | -0.001 | 0.001 | 25 942 | 0.439 |  |
|  | SBP | 2.03e-05 | 3.42e-05 | 25 942 | 0.552 |  |
|  | Fasting time | -2.42e-04 | 3.13e-04 | 25 942 | 0.439 |  |
|  | Serum cholesterol | 2.60e-04 | 5.35e-04 | 25 942 | 0.627 |  |
|  | Blood glucose | -1.03e-04 | 4.53e-04 | 25 942 | 0.820 |  |
|  | HADS - Depression | 3.36e-04 | 2.56e-04 | 25 942 | 0.190 |  |
|  | HADS - Anxiety | -1.92e-04 | 2.26e-04 | 25 942 | 0.395 |  |
|  | Sleep medication | -0.002 | 0.003 | 25 942 | 0.412 |  |
|  | Chronic illness | 0.001 | 0.001 | 25 942 | 0.713 |  |

wGRS, weighted genetic risk score; SE, standard error; BMI, body mass index; SBP, systolic blood pressure; HADS, Hospital Anxiety and Depression Scale.

Coefficients are in terms of an average-SNP increase in the allele score per unit/level increase in confounder.

* Associations surpassing multiple-testing corrected p-value threshold of 0.05/64 = 7.81e-04

Table S23: One-sample Mendelian randomization analysis for risk of incident atrial fibrillation associated with sleep traits with and without adjustment for potential confounders in UK Biobank and HUNT2

| **Sleep trait** | **UK Biobank** | | | | |  | **HUNT2** | | | | |
| --- | --- | --- | --- | --- | --- | --- | --- | --- | --- | --- | --- |
|  | **MR estimates ^†^** | |  | **MR estimates adjusted for potential confounders ^*^** | |  | **MR estimates ^‡^** | |  | **MR estimates adjusted for potential confounders ^**^** | |
|  | **N**  **(incident cases)** | **Hazard ratio**  **(95% CI)** |  | **N**  **(incident cases)** | **Hazard ratio (95% CI)** |  | **N**  **(incident cases)** | **Hazard ratio (95% CI)** |  | **N**  **(incident cases)** | **Hazard ratio**  **(95% CI)** |
| Insomnia symptoms ^#^ | 331 748  (19 530) | 1.14  (1.07, 1.21) |  | 265 290  (15 226) | 1.05  (0.97, 1.13) |  | 45 322  (6 296) | 0.95  (0.81, 1.11) |  | 38 293  (4 834) | 0.87  (0.70, 1.09) |
| 24-hour sleep duration (h) | 331 748  (19 530) | 0.74  (0.62, 0.88) |  | 265 290  (15 226) | 0.82  (0.67, 1.00) |  | 45 322  (6 296) | 1.06  (0.52, 2.15) |  | 38 293  (4 834) | 1.39  (0.66, 2.91) |
| Short sleep ^#^  (≤6 h vs. 7-8 h) | 306 292  (17 498) | 1.14  (1.04, 1.26) |  | 245 501  (13 668) | 1.09  (0.98, 1.22) |  | 33 588  (4 198) | 1.41  (0.57, 3.46) |  | 29 032  (3 326) | 0.67  (0.24, 1.85) |
| Long sleep ^#^  (≥9 h vs. 7-8 h) | 253 111  (14 738) | 0.92  (0.81, 1.05) |  | 204 188  (11 626) | 1.00  (0.87, 1.15) |  | 42 506  (5 932) | 1.70  (0.68, 4.25) |  | 35 929  (4 561) | 2.01  (0.57, 7.08) |
| Chronotype ^#^  (morning preference) | 331 748  (19 530) | 1.03  (0.99, 1.06) |  | 265 290  (15 226) | 1.02  (0.98, 1.06) |  | - | - |  | - | - |

CI, confidence interval.

**^†^** Derived using unweighted genetic risk score for each sleep trait, with adjustment for age, sex, assessment center, 40 genetic principal components, and genotyping chip.

**^‡^** Derived using weighted genetic risk score for each sleep trait, with adjustment for age, sex, 20 genetic principal components, and genotyping chip.

**^*^** Additionally adjusted for alcohol intake frequency, smoking status, body mass index, physical activity, Townsend deprivation index, education, depression, and chronic illness.

**^**^** Additionally adjusted for smoking status, body mass index, serum cholesterol levels, and anxiety.

**^#^** Hazard ratio (95% CI) scaled to per doubling in odds of the sleep trait.

Table S24: Sensitivity analysis for risk of incident atrial fibrillation associated with sleep traits in UK Biobank

| **Sleep trait** | **UK Biobank ^†^** | | | | | |
| --- | --- | --- | --- | --- | --- | --- |
|  | **N**  **(incident cases)** | **TSPS**  **HR (95% CI) ^§^** | **IVW**  **HR (95% CI)** | **MR-Egger**  **HR (95% CI)** | **Weighted Median**  **HR (95% CI)** | **Weighted Mode-based**  **HR (95% CI)** |
| Insomnia symptoms ^#^ | 331 748  (19 530) | 1.14  (1.07, 1.21) | 1.12  (1.06, 1.18) | 1.08  (0.95, 1.22);  *Int: 0.001*  *(-0.002, 0.003)* | 1.17  (1.08, 1.27) | 1.21  (1.02, 1.44) |
| 24-hour sleep duration (h) | 331 748  (19 530) | 0.74  (0.62, 0.88) | 0.75  (0.61, 0.91) | 0.86  (0.42, 1.74);  *Int: -0.002*  *(0.006, -0.014)* | 0.87  (0.67, 1.12) | 0.85  (0.53, 1.38) |
| Short sleep ^#^  (≤6 h vs. 7-8 h) | 306 292  (17 498) | 1.14  (1.04, 1.26) | 1.16  (1.05, 1.29) | 1.56  (1.04, 2.35);  *Int: -0.011*  *(-0.025, 0.004)* | 1.16  (1.01, 1.33) | 1.18  (0.93, 1.50) |
| Long sleep ^#^  (≥9 h vs. 7-8 h) | 253 111  (14 738) | 0.92  (0.81, 1.05) | 0.92  (0.79, 1.07) | 0.91  (0.60, 1.39);  *Int: 0.001*  *(-0.028, 0.029)* | 0.92  (0.78, 1.10) | 0.95  (0.71, 1.27) |
| Chronotype ^#^  (morning preference) | 331 748  (19 530) | 1.03  (0.99, 1.06) | 1.02  (0.98, 1.06) | 0.96  (0.86, 1.07);  *Int: 0.002*  *(-0.001, 0.005)* | 1.02  (0.98, 1.08) | 1.04  (0.85, 1.29) |

TSPS, two-stage predictor substitution; IVW, inverse variance weighted; HR, hazard ratio; CI, confidence interval; and Int, intercept.

**^†^** Adjusted for age, sex, assessment center, 40 genetic principal components, and genotyping chip.

**^§^** Derived using unweighted genetic risk score for each sleep trait

**^#^** Hazard ratio (95% CI) scaled to per doubling in odds of the sleep trait

Table S25: Sensitivity analysis for risk of incident atrial fibrillation associated with sleep traits in HUNT2

| **Sleep trait** | **HUNT2 ^†^** | | | | | |
| --- | --- | --- | --- | --- | --- | --- |
|  | **N**  **(incident cases)** | **TSPS**  **HR (95% CI) ^§^** | **IVW**  **HR (95% CI)** | **MR-Egger**  **HR (95% CI)** | **Weighted Median**  **HR (95% CI)** | **Weighted Mode-based**  **HR (95% CI)** |
| Insomnia symptoms ^#^ | 45 322  (6 296) | 0.95  (0.81, 1.11) | 0.97  (0.90, 1.04) | 0.97  (0.87, 1.08);  *Int: 0.001*  *(-0.003, 0.003)* | 0.96  (0.87, 1.06) | 1.04  (0.81, 1.33) |
| 24-hour sleep duration (h) | 45 322  (6 296) | 1.06  (0.52, 2.15) | 0.95  (0.69, 1.31) | 1.18  (0.67, 2.05);  *Int: -0.004*  *(-0.012, 0.004)* | 0.96  (0.59, 1.56) | 1.04  (0.54, 2.01) |
| Short sleep ^#^  (≤6 h vs. 7-8 h) | 33 588  (4 198) | 1.41  (0.57, 3.46) | 0.92  (0.78, 1.09) | 1.02  (0.75, 1.38);  *Int: -0.005*  *(-0.018, 0.008)* | 0.97  (0.76, 1.26) | 0.95  (0.75, 1.20) |
| Long sleep ^#^  (≥9 h vs. 7-8 h) | 42 506  (5 932) | 1.70  (0.68, 4.25) | 0.86  (0.61, 1.20) | 0.78  (0.47, 1.30);  *Int: 0.005*  *(-0.016, 0.027)* | 0.71  (0.36, 1.40) | 0.64  (0.38, 1.07) |

TSPS, two-stage predictor substitution; IVW, inverse variance weighted; HR, hazard ratio; CI, confidence interval; and Int, intercept.

**^†^** Adjusted for age, sex, 20 genetic principal components, and genotyping chip.

**^§^** Derived using weighted genetic risk score for each sleep trait.

**^#^** Hazard ratio (95% CI) scaled to per doubling in odds of the sleep trait.

Table S26: One-sample Mendelian randomization Cox regression analysis for risk of incident atrial fibrillation associated with insomnia symptoms using instruments from Lane et al., 2019^21^ in UK Biobank and HUNT2

|  | **N**  **(incident cases)** | **TSPS**  **HR (95% CI) ^§^** | **IVW**  **HR (95% CI)** | **MR-Egger**  **HR (95% CI)** | **Weighted Median**  **HR (95% CI)** | **Weighted Mode-based**  **HR (95% CI)** |
| --- | --- | --- | --- | --- | --- | --- |
| **UK Biobank ^†^** | 331 748  (19 530) | 1.08  (0.99, 1.18) | 1.05  (0.96, 1.14) | 0.91  (0.75, 1.12);  *Int: 0.004*  *(-0.001, 0.009)* | 1.11  (0.98, 1.26) | 1.10  (0.88, 1.38) |
| **HUNT2 ^‡^** | 45 322  (6 296) | 1.11  (0.87, 1.41) | 1.01  (0.88, 1.18) | 1.01  (0.80, 1.28);  *Int: 0.0004*  *(-0.007, 0.007)* | 1.00  (0.82, 1.22) | 1.08  (0.86, 1.37) |

TSPS, two-stage predictor substitution; IVW, inverse variance weighted; HR, hazard ratio; CI, confidence interval; and Int, intercept.

Hazard ratio (95% CI) scaled to per doubling in odds of the sleep trait.

**^§^** Derived using unweighted genetic risk score for insomnia symptoms in UK Biobank and weighted genetic risk score for insomnia symptoms in HUNT2.

**^†^** Adjusted for age, sex, assessment center, 40 genetic principal components, and genotyping chip.

**^‡^** Adjusted for age, sex, 20 genetic principal components, and genotyping chip.

Table S27: Sensitivity analysis for risk of incident atrial fibrillation associated with insomnia symptoms and chronotype in UK Biobank using genetic variants genome-wide significant in 23andMe

| **Sleep trait** | **UK Biobank ^†^** | | | | | | |
| --- | --- | --- | --- | --- | --- | --- | --- |
|  | **N**  **(incident cases)** | **TSPS**  **HR (95% CI) ^§^** | **TSPS**  **HR (95% CI) *** | **IVW**  **HR (95% CI)** | **MR-Egger**  **HR (95% CI)** | **Weighted Median**  **HR (95% CI)** | **Weighted Mode-based**  **HR (95% CI)** |
| Insomnia symptoms ^#^ | 331 748  (19 530) | 1.23  (1.11, 1.35) | 1.25  (1.13, 1.39) | 1.18  (1.08, 1.29) | 1.08  (0.91, 1.27);  *Int: 0.002*  *(-0.001, 0.005)* | 1.24  (1.09, 1.42) | 1.30  (1.06, 1.59) |
| Chronotype ^#^  (morning preference) | 331 748  (19 530) | 0.99  (0.93, 1.04) | 0.99  (0.93, 1.05) | 0.99  (0.92, 1.06) | 0.98  (0.83, 1.14);  *Int: 0.001*  *(-0.005, 0.006)* | 0.98  (0.90, 1.07) | 0.91  (0.71, 1.15) |

TSPS, two-stage predictor substitution; IVW, inverse variance weighted; HR, hazard ratio; CI, confidence interval; and Int, intercept.

**^†^** Adjusted for age, sex, assessment center, 40 genetic principal components, and genotyping chip.

**^§^** Derived using weighted genetic risk score for each sleep trait.

**^*^** Derived using unweighted genetic risk score for each sleep trait.

**^#^** Hazard ratio (95% CI) scaled to per doubling in odds of the sleep trait

Table S28: Sensitivity analysis for risk of incident atrial fibrillation associated with insomnia symptoms in HUNT2 using genetic variants genome-wide significant in 23andMe

| **Sleep trait** | **HUNT2 ^†^** | | | | | | |
| --- | --- | --- | --- | --- | --- | --- | --- |
|  | **N**  **(incident cases)** | **TSPS**  **HR (95% CI) ^§^** | **TSPS**  **HR (95% CI) *** | **IVW**  **HR (95% CI)** | **MR-Egger**  **HR (95% CI)** | **Weighted Median**  **HR (95% CI)** | **Weighted Mode-based**  **HR (95% CI)** |
| Insomnia symptoms ^#^ | 45 322  (6 296) | 0.85  (0.69, 1.05) | 0.86  (0.69, 1.09) | 0.92  (0.85, 1.01) | 0.96  (0.84, 1.10);  *Int: -0.001*  *(-0.006, 0.003)* | 0.96  (0.85, 1.09) | 0.93  (0.76, 1.13) |

TSPS, two-stage predictor substitution; IVW, inverse variance weighted; HR, hazard ratio; CI, confidence interval; and Int, intercept.

**^†^** Adjusted for age, sex, 20 genetic principal components, and genotyping chip.

**^§^** Derived using weighted genetic risk score for insomnia symptoms.

**^*^** Derived using unweighted genetic risk score for insomnia symptoms.

**^#^** Hazard ratio (95% CI) scaled to per doubling in odds of the sleep trait.

Table S29: STROBE-MR checklist of recommended items to address in reports of Mendelian randomization studies^22,23^

| **Item No.** | **Section** | **Checklist item** | **Relevant text from manuscript** |
| --- | --- | --- | --- |
| 1 | **TITLE and ABSTRACT** | Indicate Mendelian randomization (MR) as the study’s design in the title and/or the abstract if that is a main purpose of the study | Detailed in the Title and Abstract sections |
|  | **INTRODUCTION** |  |  |
| 2 | **Background** | Explain the scientific background and rationale for the reported study. What is the exposure? Is a potential causal relationship between exposure and outcome plausible? Justify why MR is a helpful method to address the study question | Detailed in the Introduction section |
| 3 | **Objectives** | State specific objectives clearly, including pre-specified causal hypotheses (if any). State that MR is a method that, under specific assumptions, intends to estimate causal effects | Detailed in the Introduction section |
|  | **METHODS** |  |  |
| 4 | **Study design and data sources** | Present key elements of the study design early in the article. Consider including a table listing sources of data for all phases of the study. For each data source contributing to the analysis, describe the following: |  |
|  | a) | Setting: Describe the study design and the underlying population, if possible. Describe the setting, locations, and relevant dates, including periods of recruitment, exposure, follow-up, and data collection, when available. | Detailed in the Methods section: Study design and population, Sleep traits, Ascertainment of atrial fibrillation, and Supplementary methods |
|  | b) | Participants: Give the eligibility criteria, and the sources and methods of selection of participants. Report the sample size, and whether any power or sample size calculations were carried out prior to the main analysis | Detailed in the Methods section: Study design and population, Sleep traits, Ascertainment of atrial fibrillation, Confounding variables, and Supplementary methods |
|  | c) | Describe measurement, quality control and selection of genetic variants | Detailed in the Methods section: Genetic variants, and Supplementary tables: Table S1 |
|  | d) | For each exposure, outcome, and other relevant variables, describe methods of assessment and diagnostic criteria for diseases | Detailed in the Methods sections: Sleep traits, Ascertainment of atrial fibrillation, Confounding variables, and Supplementary methods |
|  | e) | Provide details of ethics committee approval and participant informed consent, if relevant | Detailed in the Methods section: Study design and population |
| 5 | **Assumptions** | Explicitly state the three core IV assumptions for the main analysis (relevance, independence and exclusion restriction) as well assumptions for any additional or sensitivity analysis | Detailed in the Methods section: Statistical analysis, and Supplementary methods |
| 6 | **Statistical methods: main analysis** | Describe statistical methods and statistics used |  |
|  | a) | Describe how quantitative variables were handled in the analyses (i.e., scale, units, model) | Detailed in the Methods section: Sleep traits, Ascertainment of atrial fibrillation, Confounding variables, and Statistical analysis |
|  | b) | Describe how genetic variants were handled in the analyses and, if applicable, how their weights were selected | Detailed in the Methods section: Genetic variants, and Statistical analysis |
|  | c) | Describe the MR estimator (e.g. two-stage least squares, Wald ratio) and related statistics. Detail the included covariates and, in case of two-sample MR, whether the same covariate set was used for adjustment in the two samples | Detailed in the Methods section: Statistical analysis - MR analysis |
|  | d) | Explain how missing data were addressed | Detailed in the Methods sections: Sleep traits, Ascertainment of atrial fibrillation, Confounding variables, and Supplementary methods |
|  | e) | If applicable, indicate how multiple testing was addressed | Not applicable |
| 7 | **Assessment of assumptions** | Describe any methods or prior knowledge used to assess the assumptions or justify their validity | Detailed in the Methods sections: Statistical analysis |
| 8 | **Sensitivity analyses and additional analyses** | Describe any sensitivity analyses or additional analyses performed (e.g. comparison of effect estimates from different approaches, independent replication, bias analytic techniques, validation of instruments, simulations) | Detailed in the Methods sections: Statistical analysis – Sensitivity analyses |
| 9 | **Software and pre-registration** |  |  |
|  | a) | Name statistical software and package(s), including version and settings used | Detailed in the Methods sections: Statistical analysis |
|  | b) | State whether the study protocol and details were pre-registered (as well as when and where) | Detailed in the Methods section: Study design and population |
|  | **RESULTS** |  |  |
| 10 | **Descriptive data** |  |  |
|  | a) | Report the numbers of individuals at each stage of included studies and reasons for exclusion. Consider use of a flow diagram | Figure 1, and Detailed in the Results section |
|  | b) | Report summary statistics for phenotypic exposure(s), outcome(s), and other relevant variables (e.g. means, SDs, proportions) | Table 1, Supplementary tables: Table S2, and S4-S10 |
|  | c) | If the data sources include meta-analyses of previous studies, provide the assessments of heterogeneity across these studies | Not applicable |
|  | d) | For two-sample MR:  i.  Provide justification of the similarity of the genetic variant-exposure associations between the exposure and outcome samples  ii.  Provide information on the number of individuals who overlap between the exposure and outcome studies | Not applicable |
| 11 | **Main results** |  |  |
|  | a) | Report the associations between genetic variant and exposure, and between genetic variant and outcome, preferably on an interpretable scale | Detailed in the Results section, and Supplementary tables: Table S3 |
|  | b) | Report MR estimates of the relationship between exposure and outcome, and the measures of uncertainty from the MR analysis, on an interpretable scale, such as odds ratio or relative risk per SD difference | Figure 2 and 3, and Detailed in the Results section |
|  | c) | If relevant, consider translating estimates of relative risk into absolute risk for a meaningful time period | Not relevant |
|  | d) | Consider plots to visualize results (e.g. forest plot, scatterplot of associations between genetic variants and outcome versus between genetic variants and exposure) | Figure 2 and 3 |
| 12 | **Assessment of assumptions** |  |  |
|  | a) | Report the assessment of the validity of the assumptions | Detailed in the Results section, and Supplementary tables: Table S3 |
|  | b) | Report any additional statistics (e.g., assessments of heterogeneity across genetic variants, such as *I^2^*, Q statistic or E-value) | Detailed in the Results section, and Supplementary tables: Table S3 |
| 13 | **Sensitivity analyses and additional analyses** |  |  |
|  | a) | Report any sensitivity analyses to assess the robustness of the main results to violations of the assumptions | Detailed in the Results section: Sensitivity analysis |
|  | b) | Report results from other sensitivity analyses or additional analyses | Detailed in the Results section: Sensitivity analysis |
|  | c) | Report any assessment of direction of causal relationship (e.g., bidirectional MR) | Not relevant |
|  | d) | When relevant, report and compare with estimates from non-MR analyses | Detailed in the Results section |
|  | e) | Consider additional plots to visualize results (e.g., leave-one-out analyses) | Supplementary figures: Figure S1-S8, and Supplementary tables: Table S11-S28 |
|  | **DISCUSSION** |  |  |
| 14 | **Key results** | Summarize key results with reference to study objectives | Detailed in the Discussion section |
| 15 | **Limitations** | Discuss limitations of the study, taking into account the validity of the IV assumptions, other sources of potential bias, and imprecision. Discuss both direction and magnitude of any potential bias and any efforts to address them | Detailed in the Discussion section: Strengths and limitations |
| 16 | **Interpretation** |  |  |
|  | a) | Meaning: Give a cautious overall interpretation of results in the context of their limitations and in comparison with other studies | Detailed in the Discussion section |
|  | b) | Mechanism: Discuss underlying biological mechanisms that could drive a potential causal relationship between the investigated exposure and the outcome, and whether the gene-environment equivalence assumption is reasonable. Use causal language carefully, clarifying that IV estimates may provide causal effects only under certain assumptions | Detailed in the Discussion section: Potential mechanisms |
|  | c) | Clinical relevance: Discuss whether the results have clinical or public policy relevance, and to what extent they inform effect sizes of possible interventions | Detailed in the Discussion section: Strengths and limitations, and the Conclusion section |
| 17 | **Generalizability** | Discuss the generalizability of the study results (a) to other populations, (b) across other exposure periods/timings, and (c) across other levels of exposure | Detailed in the Discussion section: Strengths and limitations |
|  | **OTHER INFORMATION** |  |  |
| 18 | **Funding** | Describe sources of funding and the role of funders in the present study and, if applicable, sources of funding for the databases and original study or studies on which the present study is based | Detailed in the Declarations section |
| 19 | **Data and data sharing** | Provide the data used to perform all analyses or report where and how the data can be accessed, and reference these sources in the article. Provide the statistical code needed to reproduce the results in the article, or report whether the code is publicly accessible and if so, where | Detailed in the Declarations section |
| 20 | **Conflicts of Interest** | All authors should declare all potential conflicts of interest | Detailed in the Declarations section |

Table S30: List of medications used to define the sleep medication covariate in UK Biobank

| **Sleep medication** | **Treatment/medication code**  (UK Biobank field ID: 20003) |
| --- | --- |
| Oxazepam | 1140863442 |
| Meprobamate | 1140863378 |
| Medazepam | 1140863372 |
| Bromazepam | 1140863318 |
| Lorazepam | 1140863302 |
| Clobazam | 1140863268 |
| Chlormezanone | 1140863262, 1140868274 |
| Temazepam | 1140863202 |
| Nitrazepam | 1140863182, 1140863104 |
| Lormetazepam | 1140863176 |
| Diazepam | 1140863152, 1141157496 |
| Zopiclone | 1140863144 |
| Triclofos sodium | 1140863140 |
| Methyprylone | 1140856040 |
| Prazepam | 1140855944 |
| Triazolam | 1140855914 |
| Ketazolam | 1140855860 |
| Dichloralphenazone | 1140855824 |
| Clomethiazole | 1140909798 |
| Zaleplon | 1141171404 |
| Butobarbital | 1141180444 |
| Clonazepam | 1140872150 |
| Flurazepam | 1140863110 |
| Loprazolam | 1140863120 |
| Alprazolam | 1140863308 |
| Butobarbitone | 1140882090 |
