## Supplementary material for "A Mendelian randomization study investigating the role of sleep traits and their joint effects on the incidence of atrial fibrillation"

**Genetic variants**

*Table G1: Summary information of genetic variants identified for insomnia symptoms.*

| Sr. No. | rsID | Effect Allele | Other Allele | Beta | Standard Error | P | Effect Allele Frequency | N | Replication in 23andMe |
| --- | --- | --- | --- | --- | --- | --- | --- | --- | --- |
| 1 | rs113851554 | T | G | 0.2062 | 0.014 | 1.56E-51 | 0.0506 | 1 331 010 | Yes |
| 2 | rs62149809 | A | G | 0.1476 | 0.025 | 5.71E-09 | 0.986 | 1 331 010 | No |
| 3 | rs1064939 | A | T | 0.1302 | 0.02 | 2.16E-10 | 0.9784 | 1 331 010 | No |
| 4 | rs79204944 | A | G | 0.0788 | 0.014 | 4.24E-08 | 0.0449 | 1 331 010 | No |
| 5 | rs72899452 | T | C | 0.0742 | 0.012 | 1.00E-09 | 0.0648 | 1 331 010 | Yes |
| 6 | rs55972276 | A | C | 0.0733 | 0.009 | 4.19E-17 | 0.1366 | 1 331 010 | Yes |
| 7 | rs77641763 | T | C | 0.0714 | 0.009 | 6.53E-15 | 0.122 | 1 331 010 | Yes |
| 8 | rs138014720 | A | T | 0.0695 | 0.013 | 3.46E-08 | 0.9406 | 1 331 010 | No |
| 9 | rs2286729 | A | G | 0.0695 | 0.011 | 5.37E-11 | 0.0862 | 1 331 010 | Yes |
| 10 | rs118166957 | T | C | 0.0677 | 0.008 | 1.95E-16 | 0.1591 | 1 331 010 | Yes |
| 11 | rs11650304 | C | G | 0.0667 | 0.012 | 1.23E-08 | 0.9308 | 1 331 010 | No |
| 12 | rs62158170 | A | G | 0.0658 | 0.007 | 1.20E-19 | 0.7856 | 1 331 010 | Yes |
| 13 | rs62264767 | A | C | 0.0649 | 0.008 | 1.63E-14 | 0.8531 | 1 331 010 | Yes |
| 14 | rs7168238 | C | G | 0.0639 | 0.011 | 1.80E-08 | 0.0743 | 1 331 010 | No |
| 15 | rs699844 | A | G | 0.0602 | 0.011 | 4.11E-08 | 0.9195 | 1 331 010 | Yes |
| 16 | rs7566062 | T | C | 0.0592 | 0.007 | 1.37E-16 | 0.2248 | 1 331 010 | Yes |
| 17 | rs28611339 | T | G | 0.0583 | 0.009 | 8.46E-11 | 0.1282 | 1 331 010 | No |
| 18 | rs1015438 | A | G | 0.0583 | 0.008 | 2.51E-14 | 0.1882 | 1 331 010 | Yes |
| 19 | rs6465151 | T | C | 0.0564 | 0.009 | 1.90E-09 | 0.1134 | 1 331 010 | No |
| 20 | rs2815757 | T | C | 0.0554 | 0.008 | 2.24E-13 | 0.8089 | 1 331 010 | Yes |
| 21 | rs16903122 | T | C | 0.0554 | 0.007 | 9.04E-16 | 0.2487 | 1 331 010 | Yes |
| 22 | rs2792990 | C | G | 0.0545 | 0.008 | 1.15E-10 | 0.8553 | 1 331 010 | Yes |
| 23 | rs62590551 | A | G | 0.0545 | 0.01 | 4.61E-09 | 0.9029 | 1 331 010 | Yes |
| 24 | rs34490907 | C | G | 0.0535 | 0.009 | 1.76E-08 | 0.8878 | 1 331 010 | No |
| 25 | rs670501 | T | C | 0.0526 | 0.007 | 7.40E-13 | 0.2133 | 1 331 010 | Yes |
| 26 | rs1927902 | T | C | 0.0526 | 0.007 | 1.15E-14 | 0.2542 | 1 331 010 | Yes |
| 27 | rs1620977 | A | G | 0.0516 | 0.007 | 2.27E-14 | 0.2696 | 1 331 010 | Yes |
| 28 | rs75452188 | A | G | 0.0516 | 0.009 | 1.58E-08 | 0.878 | 1 331 010 | No |
| 29 | rs71575448 | A | G | 0.0507 | 0.009 | 3.38E-09 | 0.8602 | 1 331 010 | No |
| 30 | rs11756035 | C | G | 0.0507 | 0.009 | 1.29E-08 | 0.1281 | 1 331 010 | No |
| 31 | rs62429521 | A | C | 0.0507 | 0.008 | 1.78E-09 | 0.1456 | 1 331 010 | No |
| 32 | rs7992992 | A | G | 0.0507 | 0.009 | 1.15E-08 | 0.1286 | 1 331 010 | Yes |
| 33 | rs908668 | T | C | 0.0497 | 0.007 | 1.41E-11 | 0.2081 | 1 331 010 | Yes |
| 34 | rs2491124 | T | C | 0.0488 | 0.006 | 8.81E-16 | 0.5758 | 1 331 010 | Yes |
| 35 | rs35322724 | A | C | 0.0488 | 0.006 | 3.75E-16 | 0.5774 | 1 331 010 | Yes |
| 36 | rs62068188 | T | C | 0.0488 | 0.008 | 1.18E-09 | 0.8339 | 1 331 010 | No |
| 37 | rs9931543 | T | C | 0.0478 | 0.007 | 1.11E-12 | 0.7359 | 1 331 010 | Yes |
| 38 | rs3902952 | T | C | 0.0478 | 0.008 | 2.55E-10 | 0.1881 | 1 331 010 | Yes |
| 39 | rs4790076 | T | C | 0.0478 | 0.008 | 1.76E-09 | 0.1742 | 1 331 010 | No |
| 40 | rs45453598 | A | T | 0.0469 | 0.008 | 4.42E-09 | 0.169 | 1 331 010 | Yes |
| 41 | rs17223714 | A | G | 0.0459 | 0.007 | 2.44E-10 | 0.7885 | 1 331 010 | Yes |
| 42 | rs9316619 | T | C | 0.0459 | 0.008 | 5.50E-09 | 0.8249 | 1 331 010 | No |
| 43 | rs429358 | T | C | 0.0459 | 0.008 | 2.13E-08 | 0.8458 | 1 331 010 | No |
| 44 | rs12310246 | A | G | 0.0450 | 0.007 | 4.74E-11 | 0.2489 | 1 331 010 | No |
| 45 | rs830716 | C | G | 0.0450 | 0.007 | 8.68E-12 | 0.7133 | 1 331 010 | Yes |
| 46 | rs67501351 | C | G | 0.0450 | 0.007 | 5.36E-11 | 0.7451 | 1 331 010 | No |
| 47 | rs34214423 | A | C | 0.0450 | 0.008 | 3.18E-09 | 0.8087 | 1 331 010 | No |
| 48 | rs12614369 | A | G | 0.0440 | 0.008 | 7.21E-09 | 0.8158 | 1 331 010 | No |
| 49 | rs116466468 | T | C | 0.0440 | 0.007 | 2.11E-10 | 0.7593 | 1 331 010 | No |
| 50 | rs2903385 | A | G | 0.0431 | 0.006 | 4.53E-13 | 0.4844 | 1 331 010 | Yes |
| 51 | rs6606731 | A | T | 0.0431 | 0.008 | 1.51E-08 | 0.1923 | 1 331 010 | No |
| 52 | rs742760 | A | T | 0.0431 | 0.008 | 2.48E-08 | 0.8155 | 1 331 010 | Yes |
| 53 | rs10800992 | T | C | 0.0421 | 0.006 | 3.84E-12 | 0.4431 | 1 331 010 | Yes |
| 54 | rs55772859 | A | C | 0.0421 | 0.006 | 4.82E-11 | 0.3106 | 1 331 010 | Yes |
| 55 | rs10865954 | T | C | 0.0421 | 0.006 | 1.92E-11 | 0.3344 | 1 331 010 | No |
| 56 | rs17005118 | A | G | 0.0421 | 0.007 | 6.13E-10 | 0.264 | 1 331 010 | No |
| 57 | rs35539975 | A | G | 0.0421 | 0.007 | 4.49E-09 | 0.7785 | 1 331 010 | No |
| 58 | rs12666306 | A | G | 0.0421 | 0.006 | 2.24E-12 | 0.5017 | 1 331 010 | Yes |
| 59 | rs2867690 | T | C | 0.0421 | 0.008 | 3.70E-08 | 0.1819 | 1 331 010 | No |
| 60 | rs12030482 | A | T | 0.0411 | 0.007 | 8.16E-09 | 0.2201 | 1 331 010 | No |
| 61 | rs17025198 | A | G | 0.0411 | 0.007 | 2.19E-08 | 0.2043 | 1 331 010 | No |
| 62 | rs56133505 | A | G | 0.0411 | 0.006 | 5.59E-12 | 0.5369 | 1 331 010 | Yes |
| 63 | rs7486418 | T | G | 0.0411 | 0.006 | 6.84E-11 | 0.657 | 1 331 010 | No |
| 64 | rs715338 | A | G | 0.0411 | 0.006 | 7.85E-12 | 0.5776 | 1 331 010 | Yes |
| 65 | rs4643373 | T | C | 0.0411 | 0.007 | 1.58E-10 | 0.7006 | 1 331 010 | No |
| 66 | rs56097173 | T | C | 0.0402 | 0.006 | 2.69E-10 | 0.6808 | 1 331 010 | No |
| 67 | rs12991815 | C | G | 0.0402 | 0.006 | 3.02E-11 | 0.4241 | 1 331 010 | Yes |
| 68 | rs12187443 | T | C | 0.0402 | 0.006 | 1.64E-10 | 0.6681 | 1 331 010 | No |
| 69 | rs9373590 | A | T | 0.0402 | 0.006 | 2.18E-11 | 0.5078 | 1 331 010 | No |
| 70 | rs4592425 | T | G | 0.0402 | 0.006 | 4.31E-10 | 0.6965 | 1 331 010 | No |
| 71 | rs11149313 | A | G | 0.0402 | 0.007 | 2.38E-09 | 0.7297 | 1 331 010 | No |
| 72 | rs6019663 | T | C | 0.0402 | 0.007 | 6.47E-10 | 0.293 | 1 331 010 | No |
| 73 | rs62194948 | C | G | 0.0392 | 0.007 | 4.64E-09 | 0.275 | 1 331 010 | Yes |
| 74 | rs6808140 | T | C | 0.0392 | 0.006 | 5.35E-11 | 0.5053 | 1 331 010 | Yes |
| 75 | rs35110063 | A | G | 0.0392 | 0.006 | 8.82E-11 | 0.4266 | 1 331 010 | Yes |
| 76 | rs1147852 | A | G | 0.0392 | 0.006 | 9.94E-10 | 0.3095 | 1 331 010 | Yes |
| 77 | rs324017 | A | C | 0.0392 | 0.007 | 1.61E-09 | 0.294 | 1 331 010 | No |
| 78 | rs6562066 | T | C | 0.0392 | 0.006 | 1.38E-10 | 0.3688 | 1 331 010 | No |
| 79 | rs1038093 | T | C | 0.0392 | 0.006 | 2.47E-10 | 0.6282 | 1 331 010 | Yes |
| 80 | rs11090039 | A | G | 0.0392 | 0.007 | 1.82E-09 | 0.2871 | 1 331 010 | No |
| 81 | rs1861412 | A | G | 0.0383 | 0.006 | 1.67E-10 | 0.4338 | 1 331 010 | No |
| 82 | rs6888135 | A | C | 0.0383 | 0.006 | 1.21E-10 | 0.4965 | 1 331 010 | No |
| 83 | rs940780 | T | C | 0.0383 | 0.006 | 8.50E-10 | 0.3588 | 1 331 010 | No |
| 84 | rs12924275 | T | C | 0.0383 | 0.007 | 1.93E-08 | 0.268 | 1 331 010 | No |
| 85 | rs2398144 | A | C | 0.0383 | 0.006 | 5.09E-10 | 0.3947 | 1 331 010 | Yes |
| 86 | rs11679943 | A | G | 0.0373 | 0.006 | 3.16E-09 | 0.3471 | 1 331 010 | Yes |
| 87 | rs6756610 | C | G | 0.0373 | 0.006 | 1.14E-09 | 0.6291 | 1 331 010 | No |
| 88 | rs62213452 | T | G | 0.0373 | 0.007 | 2.39E-08 | 0.2788 | 1 331 010 | No |
| 89 | rs7040224 | A | G | 0.0373 | 0.006 | 4.24E-09 | 0.3161 | 1 331 010 | No |
| 90 | rs72773790 | T | C | 0.0373 | 0.006 | 3.71E-09 | 0.6732 | 1 331 010 | No |
| 91 | rs5877 | T | C | 0.0363 | 0.006 | 1.23E-08 | 0.6691 | 1 331 010 | No |
| 92 | rs1530938 | A | G | 0.0363 | 0.006 | 8.82E-10 | 0.4423 | 1 331 010 | No |
| 93 | rs7625896 | A | G | 0.0363 | 0.006 | 5.28E-09 | 0.6545 | 1 331 010 | No |
| 94 | rs1264419 | C | G | 0.0363 | 0.006 | 8.91E-10 | 0.5128 | 1 331 010 | Yes |
| 95 | rs2737240 | A | G | 0.0363 | 0.007 | 3.37E-08 | 0.7076 | 1 331 010 | No |
| 96 | rs10756571 | T | C | 0.0363 | 0.006 | 1.80E-08 | 0.6853 | 1 331 010 | No |
| 97 | rs2221119 | C | G | 0.0363 | 0.006 | 2.00E-09 | 0.443 | 1 331 010 | No |
| 98 | rs9540729 | A | T | 0.0363 | 0.006 | 1.40E-09 | 0.4794 | 1 331 010 | Yes |
| 99 | rs34967082 | A | G | 0.0354 | 0.006 | 4.34E-09 | 0.4136 | 1 331 010 | No |
| 100 | rs2216427 | C | G | 0.0354 | 0.006 | 1.60E-08 | 0.6527 | 1 331 010 | Yes |
| 101 | rs6601080 | A | G | 0.0354 | 0.006 | 2.21E-08 | 0.6762 | 1 331 010 | No |
| 102 | rs2598293 | T | C | 0.0354 | 0.006 | 2.48E-09 | 0.4763 | 1 331 010 | No |
| 103 | rs871994 | A | C | 0.0354 | 0.006 | 5.50E-09 | 0.4352 | 1 331 010 | Yes |
| 104 | rs1167132 | T | C | 0.0354 | 0.006 | 8.73E-09 | 0.3917 | 1 331 010 | Yes |
| 105 | rs176644 | T | G | 0.0354 | 0.006 | 9.49E-09 | 0.4036 | 1 331 010 | No |
| 106 | rs12605642 | T | G | 0.0354 | 0.006 | 2.13E-09 | 0.4864 | 1 331 010 | No |
| 107 | rs9964420 | A | C | 0.0354 | 0.007 | 4.54E-08 | 0.301 | 1 331 010 | Yes |
| 108 | rs72820274 | A | G | 0.0344 | 0.006 | 1.28E-08 | 0.417 | 1 331 010 | No |
| 109 | rs10928256 | T | C | 0.0344 | 0.006 | 1.61E-08 | 0.4192 | 1 331 010 | No |
| 110 | rs4260410 | T | C | 0.0344 | 0.006 | 4.87E-08 | 0.332 | 1 331 010 | Yes |
| 111 | rs11722569 | T | C | 0.0344 | 0.006 | 2.91E-08 | 0.6586 | 1 331 010 | Yes |
| 112 | rs13138995 | A | G | 0.0344 | 0.006 | 1.97E-08 | 0.3896 | 1 331 010 | No |
| 113 | rs6978112 | T | C | 0.0344 | 0.006 | 2.11E-08 | 0.4105 | 1 331 010 | No |
| 114 | rs2030672 | C | G | 0.0344 | 0.006 | 1.10E-08 | 0.5589 | 1 331 010 | No |
| 115 | rs874168 | T | C | 0.0344 | 0.006 | 7.95E-09 | 0.5254 | 1 331 010 | Yes |
| 116 | rs10898940 | A | C | 0.0344 | 0.006 | 8.09E-09 | 0.5173 | 1 331 010 | No |
| 117 | rs1567084 | A | G | 0.0334 | 0.006 | 2.14E-08 | 0.4981 | 1 331 010 | No |
| 118 | rs1580173 | A | G | 0.0334 | 0.006 | 2.28E-08 | 0.5608 | 1 331 010 | No |
| 119 | rs1357685 | T | C | 0.0334 | 0.006 | 1.39E-08 | 0.4735 | 1 331 010 | No |
| 120 | rs4588900 | A | G | 0.0334 | 0.006 | 1.57E-08 | 0.5164 | 1 331 010 | No |
| 121 | rs28552587 | A | G | 0.0334 | 0.006 | 3.30E-08 | 0.5641 | 1 331 010 | No |
| 122 | rs10955647 | T | G | 0.0334 | 0.006 | 1.84E-08 | 0.5321 | 1 331 010 | No |
| 123 | rs6597649 | T | C | 0.0334 | 0.006 | 3.05E-08 | 0.3994 | 1 331 010 | No |
| 124 | rs10825503 | T | G | 0.0334 | 0.006 | 1.43E-08 | 0.4873 | 1 331 010 | No |
| 125 | rs667730 | T | C | 0.0334 | 0.006 | 2.26E-08 | 0.5788 | 1 331 010 | Yes |
| 126 | rs647905 | T | C | 0.0334 | 0.006 | 2.87E-08 | 0.5409 | 1 331 010 | Yes |
| 127 | rs10947987 | T | C | -0.0325 | 0.006 | 4.08E-08 | 0.4431 | 1 331 010 | No |
| 128 | rs4858708 | A | T | -0.0336 | 0.006 | 1.23E-08 | 0.5305 | 1 331 010 | No |
| 129 | rs2364921 | T | C | -0.0336 | 0.006 | 2.13E-08 | 0.4691 | 1 331 010 | No |
| 130 | rs238869 | T | C | -0.0336 | 0.006 | 3.36E-08 | 0.6229 | 1 331 010 | No |
| 131 | rs190073 | A | G | -0.0336 | 0.006 | 2.86E-08 | 0.4141 | 1 331 010 | No |
| 132 | rs9563886 | T | C | -0.0336 | 0.006 | 3.08E-08 | 0.6063 | 1 331 010 | No |
| 133 | rs2447094 | A | C | -0.0336 | 0.006 | 2.50E-08 | 0.4687 | 1 331 010 | No |
| 134 | rs1553754 | T | G | -0.0336 | 0.006 | 3.51E-08 | 0.5621 | 1 331 010 | Yes |
| 135 | rs11588755 | A | G | -0.0346 | 0.006 | 5.14E-09 | 0.522 | 1 331 010 | No |
| 136 | rs11119409 | T | C | -0.0346 | 0.006 | 1.19E-08 | 0.5866 | 1 331 010 | No |
| 137 | rs728017 | A | G | -0.0346 | 0.006 | 9.51E-09 | 0.3864 | 1 331 010 | No |
| 138 | rs1731951 | A | T | -0.0346 | 0.006 | 1.36E-08 | 0.4434 | 1 331 010 | No |
| 139 | rs4788203 | A | G | -0.0346 | 0.006 | 6.32E-09 | 0.4334 | 1 331 010 | Yes |
| 140 | rs12454003 | C | G | -0.0346 | 0.006 | 4.94E-09 | 0.4821 | 1 331 010 | No |
| 141 | rs910187 | A | G | -0.0346 | 0.006 | 1.63E-08 | 0.373 | 1 331 010 | No |
| 142 | rs34036083 | T | C | -0.0356 | 0.006 | 2.07E-08 | 0.6576 | 1 331 010 | No |
| 143 | rs12520974 | T | C | -0.0356 | 0.006 | 1.69E-09 | 0.4846 | 1 331 010 | No |
| 144 | rs701394 | A | G | -0.0356 | 0.006 | 6.83E-09 | 0.6376 | 1 331 010 | No |
| 145 | rs37445 | A | G | -0.0356 | 0.006 | 4.88E-09 | 0.3905 | 1 331 010 | No |
| 146 | rs17367725 | T | C | -0.0356 | 0.006 | 9.29E-09 | 0.3513 | 1 331 010 | Yes |
| 147 | rs9469434 | C | G | -0.0356 | 0.007 | 4.41E-08 | 0.2851 | 1 331 010 | Yes |
| 148 | rs10758593 | A | G | -0.0356 | 0.006 | 4.90E-09 | 0.3989 | 1 331 010 | Yes |
| 149 | rs7402939 | T | C | -0.0356 | 0.006 | 5.19E-09 | 0.376 | 1 331 010 | Yes |
| 150 | rs2838787 | A | G | -0.0356 | 0.006 | 7.65E-09 | 0.3924 | 1 331 010 | No |
| 151 | rs6702604 | A | G | -0.0367 | 0.006 | 1.30E-09 | 0.5843 | 1 331 010 | No |
| 152 | rs823247 | T | C | -0.0367 | 0.006 | 5.25E-10 | 0.479 | 1 331 010 | No |
| 153 | rs1519102 | C | G | -0.0367 | 0.006 | 1.90E-08 | 0.6891 | 1 331 010 | No |
| 154 | rs1064213 | A | G | -0.0367 | 0.006 | 6.41E-10 | 0.4789 | 1 331 010 | Yes |
| 155 | rs7599697 | T | C | -0.0367 | 0.006 | 5.00E-09 | 0.3583 | 1 331 010 | Yes |
| 156 | rs2388840 | A | G | -0.0367 | 0.006 | 1.37E-09 | 0.5757 | 1 331 010 | Yes |
| 157 | rs7475916 | C | G | -0.0367 | 0.006 | 6.70E-09 | 0.3533 | 1 331 010 | No |
| 158 | rs4767645 | T | G | -0.0367 | 0.006 | 6.47E-10 | 0.4614 | 1 331 010 | No |
| 159 | rs6510033 | A | G | -0.0367 | 0.007 | 4.66E-08 | 0.7253 | 1 331 010 | Yes |
| 160 | rs623025 | T | C | -0.0377 | 0.007 | 3.16E-08 | 0.2552 | 1 331 010 | No |
| 161 | rs73163783 | T | C | -0.0377 | 0.007 | 1.39E-08 | 0.7232 | 1 331 010 | No |
| 162 | rs10944696 | A | G | -0.0377 | 0.007 | 7.99E-09 | 0.2978 | 1 331 010 | No |
| 163 | rs6973090 | A | G | -0.0377 | 0.007 | 4.31E-08 | 0.25 | 1 331 010 | No |
| 164 | rs671985 | A | G | -0.0377 | 0.006 | 2.79E-10 | 0.4516 | 1 331 010 | No |
| 165 | rs11001276 | A | T | -0.0377 | 0.007 | 2.52E-08 | 0.74 | 1 331 010 | No |
| 166 | rs214934 | A | T | -0.0377 | 0.006 | 3.16E-09 | 0.3123 | 1 331 010 | Yes |
| 167 | rs6589988 | A | G | -0.0377 | 0.006 | 4.70E-09 | 0.6757 | 1 331 010 | Yes |
| 168 | rs8181889 | A | G | -0.0377 | 0.006 | 8.90E-10 | 0.3992 | 1 331 010 | No |
| 169 | rs1536053 | T | C | -0.0377 | 0.006 | 6.04E-09 | 0.3157 | 1 331 010 | Yes |
| 170 | rs3184470 | A | G | -0.0377 | 0.006 | 9.73E-10 | 0.3507 | 1 331 010 | No |
| 171 | rs8076183 | T | C | -0.0377 | 0.006 | 2.75E-10 | 0.4485 | 1 331 010 | Yes |
| 172 | rs1937447 | C | G | -0.0387 | 0.007 | 2.08E-08 | 0.7593 | 1 331 010 | Yes |
| 173 | rs7571486 | A | G | -0.0387 | 0.007 | 1.40E-08 | 0.251 | 1 331 010 | No |
| 174 | rs4502882 | T | C | -0.0387 | 0.006 | 7.96E-10 | 0.6578 | 1 331 010 | Yes |
| 175 | rs6457796 | T | C | -0.0387 | 0.007 | 1.12E-08 | 0.7313 | 1 331 010 | No |
| 176 | rs4090240 | T | C | -0.0387 | 0.007 | 8.46E-09 | 0.2775 | 1 331 010 | Yes |
| 177 | rs12251016 | A | T | -0.0387 | 0.006 | 3.89E-10 | 0.6559 | 1 331 010 | Yes |
| 178 | rs224029 | T | C | -0.0387 | 0.006 | 2.51E-10 | 0.3995 | 1 331 010 | No |
| 179 | rs566673 | T | G | -0.0387 | 0.006 | 1.18E-10 | 0.5351 | 1 331 010 | Yes |
| 180 | rs10502966 | A | G | -0.0387 | 0.006 | 8.54E-11 | 0.582 | 1 331 010 | Yes |
| 181 | rs7615602 | C | G | -0.0398 | 0.007 | 2.59E-09 | 0.2715 | 1 331 010 | No |
| 182 | rs521484 | A | G | -0.0398 | 0.007 | 1.53E-08 | 0.7668 | 1 331 010 | Yes |
| 183 | rs75932578 | T | C | -0.0398 | 0.007 | 4.15E-08 | 0.2159 | 1 331 010 | No |
| 184 | rs12790660 | T | C | -0.0398 | 0.006 | 4.49E-10 | 0.6844 | 1 331 010 | No |
| 185 | rs2389631 | A | C | -0.0398 | 0.006 | 2.03E-10 | 0.6666 | 1 331 010 | Yes |
| 186 | rs2089358 | T | C | -0.0408 | 0.007 | 2.75E-10 | 0.7038 | 1 331 010 | Yes |
| 187 | rs1289939 | T | C | -0.0408 | 0.007 | 6.00E-09 | 0.2325 | 1 331 010 | No |
| 188 | rs11803128 | A | G | -0.0408 | 0.006 | 6.85E-11 | 0.6541 | 1 331 010 | Yes |
| 189 | rs6545798 | A | T | -0.0408 | 0.006 | 1.19E-11 | 0.4104 | 1 331 010 | Yes |
| 190 | rs4664299 | T | C | -0.0408 | 0.007 | 4.95E-09 | 0.2349 | 1 331 010 | Yes |
| 191 | rs3774751 | T | G | -0.0408 | 0.006 | 7.32E-12 | 0.4621 | 1 331 010 | Yes |
| 192 | rs7044885 | C | G | -0.0408 | 0.006 | 5.67E-12 | 0.4417 | 1 331 010 | Yes |
| 193 | rs6734957 | T | G | -0.0419 | 0.007 | 1.82E-09 | 0.2388 | 1 331 010 | Yes |
| 194 | rs62301574 | C | G | -0.0419 | 0.007 | 1.37E-08 | 0.7996 | 1 331 010 | Yes |
| 195 | rs12917449 | A | C | -0.0419 | 0.008 | 2.97E-08 | 0.8061 | 1 331 010 | No |
| 196 | rs9889282 | A | C | -0.0419 | 0.006 | 4.70E-12 | 0.6129 | 1 331 010 | Yes |
| 197 | rs11126082 | C | G | -0.0429 | 0.006 | 8.26E-13 | 0.4398 | 1 331 010 | Yes |
| 198 | rs984306 | T | C | -0.0429 | 0.007 | 7.94E-10 | 0.7547 | 1 331 010 | No |
| 199 | rs314281 | T | C | -0.0429 | 0.006 | 6.03E-13 | 0.4531 | 1 331 010 | No |
| 200 | rs10761240 | A | G | -0.0429 | 0.006 | 2.12E-12 | 0.3963 | 1 331 010 | Yes |
| 201 | rs12912299 | T | C | -0.0429 | 0.006 | 4.42E-13 | 0.4893 | 1 331 010 | Yes |
| 202 | rs4238755 | A | C | -0.0429 | 0.007 | 2.30E-10 | 0.2638 | 1 331 010 | Yes |
| 203 | rs60565673 | T | G | -0.0429 | 0.006 | 1.59E-12 | 0.6211 | 1 331 010 | Yes |
| 204 | rs12983032 | A | G | -0.0429 | 0.006 | 1.07E-11 | 0.3434 | 1 331 010 | No |
| 205 | rs694786 | T | C | -0.0440 | 0.006 | 1.97E-13 | 0.4605 | 1 331 010 | Yes |
| 206 | rs17083297 | A | C | -0.0440 | 0.008 | 1.60E-08 | 0.1766 | 1 331 010 | No |
| 207 | rs3131638 | A | G | -0.0440 | 0.007 | 7.88E-10 | 0.2261 | 1 331 010 | No |
| 208 | rs6967168 | T | G | -0.0440 | 0.007 | 1.39E-10 | 0.7544 | 1 331 010 | No |
| 209 | rs524859 | A | G | -0.0440 | 0.006 | 1.48E-12 | 0.3601 | 1 331 010 | No |
| 210 | rs61921611 | T | C | -0.0440 | 0.006 | 7.84E-12 | 0.692 | 1 331 010 | Yes |
| 211 | rs7214267 | A | G | -0.0440 | 0.006 | 5.09E-13 | 0.581 | 1 331 010 | No |
| 212 | rs61765555 | T | C | -0.0450 | 0.007 | 4.00E-11 | 0.2552 | 1 331 010 | Yes |
| 213 | rs11605348 | A | G | -0.0450 | 0.006 | 7.01E-13 | 0.3495 | 1 331 010 | Yes |
| 214 | rs16990210 | T | C | -0.0460 | 0.008 | 1.97E-08 | 0.8478 | 1 331 010 | No |
| 215 | rs12540241 | A | T | -0.0460 | 0.008 | 1.58E-09 | 0.1926 | 1 331 010 | No |
| 216 | rs10947690 | A | G | -0.0471 | 0.007 | 4.04E-12 | 0.7408 | 1 331 010 | Yes |
| 217 | rs4702 | A | G | -0.0481 | 0.006 | 6.78E-16 | 0.5562 | 1 331 010 | Yes |
| 218 | rs73079014 | T | C | -0.0492 | 0.009 | 3.65E-08 | 0.126 | 1 331 010 | No |
| 219 | rs8180817 | C | G | -0.0492 | 0.006 | 1.83E-16 | 0.4304 | 1 331 010 | Yes |
| 220 | rs76145129 | T | G | -0.0502 | 0.009 | 2.73E-08 | 0.1238 | 1 331 010 | No |
| 221 | rs1031654 | A | C | -0.0513 | 0.007 | 3.88E-12 | 0.7996 | 1 331 010 | No |
| 222 | rs152555 | A | G | -0.0523 | 0.008 | 4.83E-10 | 0.8544 | 1 331 010 | No |
| 223 | rs2431108 | T | C | -0.0534 | 0.006 | 7.83E-17 | 0.672 | 1 331 010 | Yes |
| 224 | rs9394502 | T | C | -0.0545 | 0.006 | 7.76E-18 | 0.3343 | 1 331 010 | Yes |
| 225 | rs4709655 | T | C | -0.0545 | 0.009 | 3.09E-09 | 0.1191 | 1 331 010 | Yes |
| 226 | rs28582096 | A | G | -0.0545 | 0.007 | 1.74E-13 | 0.205 | 1 331 010 | Yes |
| 227 | rs4981170 | A | G | -0.0545 | 0.008 | 7.33E-13 | 0.1943 | 1 331 010 | Yes |
| 228 | rs72657797 | T | C | -0.0555 | 0.008 | 1.52E-12 | 0.1759 | 1 331 010 | Yes |
| 229 | rs8180457 | T | C | -0.0555 | 0.008 | 1.12E-11 | 0.1573 | 1 331 010 | Yes |
| 230 | rs73671843 | A | G | -0.0555 | 0.009 | 5.49E-10 | 0.1257 | 1 331 010 | No |
| 231 | rs17324524 | T | C | -0.0576 | 0.009 | 5.01E-10 | 0.883 | 1 331 010 | Yes |
| 232 | rs13010288 | T | G | -0.0598 | 0.009 | 9.26E-12 | 0.1326 | 1 331 010 | Yes |
| 233 | rs62383308 | A | G | -0.0598 | 0.011 | 3.98E-08 | 0.0805 | 1 331 010 | Yes |
| 234 | rs17643634 | T | C | -0.0598 | 0.008 | 1.34E-13 | 0.165 | 1 331 010 | Yes |
| 235 | rs66674044 | A | T | -0.0598 | 0.009 | 2.18E-12 | 0.8573 | 1 331 010 | Yes |
| 236 | rs6119267 | C | G | -0.0598 | 0.006 | 2.32E-20 | 0.6891 | 1 331 010 | Yes |
| 237 | rs492858 | T | C | -0.0661 | 0.011 | 3.46E-09 | 0.0759 | 1 331 010 | Yes |
| 238 | rs10947428 | T | C | -0.0683 | 0.007 | 9.06E-21 | 0.7858 | 1 331 010 | Yes |
| 239 | rs79693059 | C | G | -0.0726 | 0.011 | 1.61E-11 | 0.9158 | 1 331 010 | Yes |
| 240 | rs9527083 | A | G | -0.0758 | 0.006 | 1.61E-32 | 0.6705 | 1 331 010 | Yes |
| 241 | rs11838830 | A | G | -0.0801 | 0.013 | 5.20E-10 | 0.9436 | 1 331 010 | Yes |
| 242 | rs4699157 | T | C | -0.0812 | 0.015 | 3.98E-08 | 0.958 | 1 331 010 | No |
| 243 | rs7432782 | T | C | -0.0834 | 0.014 | 7.42E-09 | 0.9558 | 1 331 010 | No |
| 244 | rs13135092 | A | G | -0.0888 | 0.011 | 2.53E-16 | 0.9175 | 1 331 010 | Yes |
| 245 | rs17520265 | A | G | -0.0910 | 0.016 | 2.87E-08 | 0.0342 | 1 331 010 | No |
| 246 | rs78206187 | A | G | -0.0943 | 0.013 | 2.96E-13 | 0.9442 | 1 331 010 | No |
| 247 | rs117630493 | C | G | -0.1009 | 0.018 | 3.61E-08 | 0.9726 | 1 331 010 | Yes |
| 248 | rs138678612 | A | G | -0.1165 | 0.02 | 1.41E-08 | 0.9783 | 1 331 010 | No |

Table G2: Summary information of genetic variants identified for chronotype.

| Sr. No. | rsID | Effect Allele | Other Allele | Beta | Standard Error | P | Effect Allele Frequency | N | Replication in 23andMe |
| --- | --- | --- | --- | --- | --- | --- | --- | --- | --- |
| 1 | rs909757 | T | C | 0.0197 | 0.003283 | 1.96E-09 | 0.63 | 651 295 | No |
| 2 | rs61773390 | T | G | 0.0659 | 0.004687 | 6.55E-45 | 0.1931 | 651 295 | Yes |
| 3 | rs12065331 | T | C | -0.0244 | 0.003462 | 1.82E-12 | 0.3125 | 651 295 | No |
| 4 | rs17448682 | T | C | 0.0348 | 0.004059 | 9.97E-18 | 0.2339 | 651 295 | No |
| 5 | rs10917513 | T | C | -0.0309 | 0.003739 | 1.41E-16 | 0.6476 | 651 295 | Yes |
| 6 | rs10916892 | T | C | -0.0345 | 0.003695 | 9.90E-21 | 0.6203 | 651 295 | Yes |
| 7 | rs2506089 | T | G | 0.0198 | 0.003403 | 5.95E-09 | 0.5682 | 651 295 | No |
| 8 | rs12140153 | T | G | -0.0604 | 0.006493 | 1.36E-20 | 0.0892 | 651 295 | Yes |
| 9 | rs11208844 | A | G | -0.029 | 0.004742 | 9.61E-10 | 0.1408 | 651 295 | No |
| 10 | rs12040629 | A | G | 0.0727 | 0.004925 | 2.57E-49 | 0.16 | 651 295 | Yes |
| 11 | rs11588913 | A | G | -0.0236 | 0.003576 | 4.11E-11 | 0.4037 | 651 295 | No |
| 12 | rs5016898 | T | C | -0.0241 | 0.003694 | 6.83E-11 | 0.4238 | 651 295 | No |
| 13 | rs72720396 | A | G | -0.0416 | 0.004203 | 4.26E-23 | 0.7741 | 651 295 | No |
| 14 | rs481214 | A | T | 0.0232 | 0.003948 | 4.17E-09 | 0.6063 | 651 295 | No |
| 15 | rs11165655 | A | G | -0.0281 | 0.003708 | 3.48E-14 | 0.5281 | 651 295 | No |
| 16 | rs17575798 | A | G | -0.0336 | 0.004233 | 2.08E-15 | 0.1918 | 651 295 | No |
| 17 | rs6690292 | T | C | -0.0247 | 0.004165 | 3.04E-09 | 0.7285 | 651 295 | No |
| 18 | rs11102807 | A | G | -0.0219 | 0.003623 | 1.50E-09 | 0.5361 | 651 295 | No |
| 19 | rs9436119 | A | G | 0.0395 | 0.003314 | 9.31E-33 | 0.3842 | 651 295 | Yes |
| 20 | rs6665637 | A | G | -0.0196 | 0.003107 | 2.80E-10 | 0.2804 | 651 295 | No |
| 21 | rs115073088 | A | G | -0.0755 | 0.010399 | 3.88E-13 | 0.975 | 651 295 | No |
| 22 | rs975025 | T | C | -0.049 | 0.006244 | 4.21E-15 | 0.0758 | 651 295 | No |
| 23 | rs1144566 | T | C | 0.2306 | 0.010615 | 1.24E-104 | 0.029 | 651 295 | Yes |
| 24 | rs1221502 | A | C | 0.0197 | 0.00331 | 2.67E-09 | 0.7386 | 651 295 | No |
| 25 | rs4657983 | A | G | -0.0252 | 0.003698 | 9.45E-12 | 0.6511 | 651 295 | No |
| 26 | rs6429233 | A | G | 0.0202 | 0.003382 | 2.33E-09 | 0.4574 | 651 295 | No |
| 27 | rs13011556 | C | G | -0.0285 | 0.003663 | 7.21E-15 | 0.7624 | 651 295 | No |
| 28 | rs62124718 | A | G | -0.0447 | 0.005831 | 1.78E-14 | 0.895 | 651 295 | Yes |
| 29 | rs72796401 | A | T | 0.0246 | 0.003266 | 5.02E-14 | 0.1903 | 651 295 | No |
| 30 | rs6718511 | A | G | 0.0209 | 0.003483 | 1.96E-09 | 0.557 | 651 295 | No |
| 31 | rs11678584 | A | T | -0.0278 | 0.004512 | 7.24E-10 | 0.8599 | 651 295 | No |
| 32 | rs848552 | C | G | -0.0282 | 0.003571 | 2.86E-15 | 0.4755 | 651 295 | No |
| 33 | rs7602499 | T | C | 0.0223 | 0.003861 | 7.63E-09 | 0.3536 | 651 295 | No |
| 34 | rs75120545 | T | C | 0.0862 | 0.00985 | 2.11E-18 | 0.0319 | 651 295 | No |
| 35 | rs6544906 | A | C | 0.0233 | 0.003588 | 8.38E-11 | 0.5616 | 651 295 | No |
| 36 | rs17396357 | T | C | 0.0211 | 0.003602 | 4.68E-09 | 0.3818 | 651 295 | No |
| 37 | rs12470914 | A | T | 0.053 | 0.005982 | 8.00E-19 | 0.1005 | 651 295 | Yes |
| 38 | rs4672458 | T | C | -0.0224 | 0.003345 | 2.13E-11 | 0.4764 | 651 295 | No |
| 39 | rs13414393 | T | C | -0.0219 | 0.003595 | 1.12E-09 | 0.5405 | 651 295 | No |
| 40 | rs10175975 | T | C | 0.0252 | 0.003946 | 1.70E-10 | 0.1853 | 651 295 | No |
| 41 | rs359248 | T | G | -0.0275 | 0.003196 | 7.69E-18 | 0.4564 | 651 295 | Yes |
| 42 | rs812925 | C | G | -0.0306 | 0.003496 | 2.06E-18 | 0.6458 | 651 295 | No |
| 43 | rs113851554 | T | G | -0.054 | 0.006988 | 1.09E-14 | 0.0567 | 651 295 | No |
| 44 | rs2706762 | T | C | -0.037 | 0.004661 | 2.05E-15 | 0.1496 | 651 295 | No |
| 45 | rs12464387 | A | G | -0.0214 | 0.003347 | 1.62E-10 | 0.4638 | 651 295 | No |
| 46 | rs6727752 | A | G | 0.0256 | 0.003851 | 2.99E-11 | 0.3569 | 651 295 | No |
| 47 | rs10520176 | T | C | 0.0377 | 0.003555 | 2.80E-26 | 0.4926 | 651 295 | Yes |
| 48 | rs11681299 | T | C | 0.0237 | 0.003887 | 1.08E-09 | 0.2835 | 651 295 | No |
| 49 | rs34509802 | A | G | 0.0396 | 0.005328 | 1.07E-13 | 0.1785 | 651 295 | Yes |
| 50 | rs76064513 | T | C | 0.0338 | 0.005825 | 6.53E-09 | 0.1342 | 651 295 | No |
| 51 | rs77248969 | A | G | -0.0333 | 0.005431 | 8.69E-10 | 0.1116 | 651 295 | No |
| 52 | rs28380327 | A | T | 0.04 | 0.003695 | 2.61E-27 | 0.6317 | 651 295 | Yes |
| 53 | rs2166559 | T | C | -0.0329 | 0.005198 | 2.47E-10 | 0.8611 | 651 295 | No |
| 54 | rs747003 | T | C | 0.0196 | 0.003367 | 5.83E-09 | 0.6088 | 651 295 | No |
| 55 | rs13004345 | T | C | -0.0189 | 0.003014 | 3.62E-10 | 0.6475 | 651 295 | No |
| 56 | rs6433478 | T | C | -0.0253 | 0.003558 | 1.16E-12 | 0.4601 | 651 295 | Yes |
| 57 | rs4666682 | A | G | -0.0254 | 0.004109 | 6.34E-10 | 0.1797 | 651 295 | No |
| 58 | rs11677484 | T | G | 0.0229 | 0.003592 | 1.82E-10 | 0.2574 | 651 295 | No |
| 59 | rs1064213 | A | G | 0.0444 | 0.00365 | 4.78E-34 | 0.4826 | 651 295 | Yes |
| 60 | rs184033703 | A | G | 0.0577 | 0.007435 | 8.41E-15 | 0.0585 | 651 295 | No |
| 61 | rs80271258 | T | C | -0.0894 | 0.006197 | 3.52E-47 | 0.0844 | 651 295 | Yes |
| 62 | rs62182135 | A | C | -0.0242 | 0.003246 | 8.97E-14 | 0.3285 | 651 295 | No |
| 63 | rs35346733 | A | G | -0.0316 | 0.004832 | 6.14E-11 | 0.1942 | 651 295 | No |
| 64 | rs111261826 | A | C | -0.028 | 0.004043 | 4.32E-12 | 0.679 | 651 295 | No |
| 65 | rs149611468 | T | C | 0.1432 | 0.017515 | 2.94E-16 | 0.9888 | 651 295 | No |
| 66 | rs6794796 | A | G | 0.0253 | 0.003834 | 4.16E-11 | 0.2897 | 651 295 | No |
| 67 | rs9817910 | A | G | -0.0216 | 0.003231 | 2.30E-11 | 0.5576 | 651 295 | No |
| 68 | rs73050286 | T | C | 0.0297 | 0.004278 | 3.88E-12 | 0.7817 | 651 295 | No |
| 69 | rs2362775 | T | C | -0.0218 | 0.003576 | 1.09E-09 | 0.5342 | 651 295 | No |
| 70 | rs114848860 | A | T | -0.0768 | 0.009878 | 7.56E-15 | 0.9737 | 651 295 | No |
| 71 | rs78580841 | T | C | 0.0424 | 0.006106 | 3.81E-12 | 0.0694 | 651 295 | No |
| 72 | rs12636669 | T | C | 0.057 | 0.006106 | 1.01E-20 | 0.0801 | 651 295 | Yes |
| 73 | rs17007397 | C | G | 0.0231 | 0.004036 | 1.05E-08 | 0.5801 | 651 295 | No |
| 74 | rs7626335 | A | C | -0.0286 | 0.003965 | 5.44E-13 | 0.3305 | 651 295 | Yes |
| 75 | rs7429614 | T | G | 0.0349 | 0.003703 | 4.36E-21 | 0.416 | 651 295 | Yes |
| 76 | rs12631477 | T | C | 0.0277 | 0.004463 | 5.42E-10 | 0.7982 | 651 295 | No |
| 77 | rs1449403 | A | G | 0.0424 | 0.005626 | 4.81E-14 | 0.1226 | 651 295 | Yes |
| 78 | rs34967119 | A | G | 0.02 | 0.003337 | 2.04E-09 | 0.4958 | 651 295 | No |
| 79 | rs1398346 | T | C | 0.0259 | 0.004513 | 9.51E-09 | 0.8668 | 651 295 | No |
| 80 | rs1800828 | C | G | 0.0257 | 0.00437 | 4.09E-09 | 0.7508 | 651 295 | No |
| 81 | rs72950188 | T | C | 0.0447 | 0.006676 | 2.14E-11 | 0.9238 | 651 295 | No |
| 82 | rs72966564 | T | C | -0.0234 | 0.003861 | 1.36E-09 | 0.2507 | 651 295 | No |
| 83 | rs13065394 | T | G | -0.0274 | 0.003962 | 4.64E-12 | 0.286 | 651 295 | No |
| 84 | rs4550782 | T | G | 0.0282 | 0.003897 | 4.60E-13 | 0.6655 | 651 295 | No |
| 85 | rs7649164 | T | G | 0.0211 | 0.003466 | 1.15E-09 | 0.5745 | 651 295 | No |
| 86 | rs6440833 | A | G | 0.021 | 0.00327 | 1.35E-10 | 0.4638 | 651 295 | No |
| 87 | rs111867612 | A | C | -0.032 | 0.005572 | 9.30E-09 | 0.1021 | 651 295 | No |
| 88 | rs1599374 | A | G | 0.0309 | 0.003738 | 1.38E-16 | 0.5163 | 651 295 | Yes |
| 89 | rs3850174 | A | T | -0.0346 | 0.004181 | 1.28E-16 | 0.2575 | 651 295 | Yes |
| 90 | rs301218 | A | G | -0.0237 | 0.003527 | 1.81E-11 | 0.3916 | 651 295 | No |
| 91 | rs9836621 | T | C | -0.0279 | 0.003617 | 1.22E-14 | 0.5217 | 651 295 | Yes |
| 92 | rs1468945 | A | G | -0.0363 | 0.00428 | 2.21E-17 | 0.7852 | 651 295 | No |
| 93 | rs3796618 | A | T | -0.0229 | 0.00381 | 1.85E-09 | 0.5293 | 651 295 | No |
| 94 | rs4690085 | A | G | -0.0193 | 0.003282 | 4.10E-09 | 0.5348 | 651 295 | No |
| 95 | rs4698678 | C | G | 0.0307 | 0.004324 | 1.24E-12 | 0.2786 | 651 295 | Yes |
| 96 | rs1502249 | A | G | 0.0173 | 0.002969 | 5.67E-09 | 0.5223 | 651 295 | No |
| 97 | rs6838677 | A | C | -0.0213 | 0.003654 | 5.58E-09 | 0.6689 | 651 295 | No |
| 98 | rs4860734 | A | G | 0.0195 | 0.003345 | 5.55E-09 | 0.2897 | 651 295 | No |
| 99 | rs6816922 | A | C | -0.02 | 0.003483 | 9.30E-09 | 0.5377 | 651 295 | No |
| 100 | rs6846730 | T | C | -0.0323 | 0.004045 | 1.39E-15 | 0.2353 | 651 295 | No |
| 101 | rs2850979 | T | C | -0.0234 | 0.003771 | 5.45E-10 | 0.7603 | 651 295 | No |
| 102 | rs7700110 | A | G | 0.0242 | 0.003927 | 7.16E-10 | 0.2566 | 651 295 | No |
| 103 | rs17455138 | T | C | 0.0307 | 0.004533 | 1.27E-11 | 0.7655 | 651 295 | Yes |
| 104 | rs4241964 | T | G | -0.0287 | 0.003405 | 3.45E-17 | 0.5217 | 651 295 | No |
| 105 | rs938836 | A | G | -0.0212 | 0.003311 | 1.53E-10 | 0.467 | 651 295 | No |
| 106 | rs72729847 | T | C | -0.0303 | 0.004502 | 1.69E-11 | 0.8005 | 651 295 | No |
| 107 | rs9997394 | A | G | -0.0254 | 0.003971 | 1.60E-10 | 0.2866 | 651 295 | No |
| 108 | rs10058356 | T | C | -0.0207 | 0.003294 | 3.28E-10 | 0.6958 | 651 295 | No |
| 109 | rs7701529 | A | T | -0.0295 | 0.004077 | 4.67E-13 | 0.2393 | 651 295 | No |
| 110 | rs7721608 | T | G | 0.0204 | 0.003246 | 3.27E-10 | 0.4648 | 651 295 | No |
| 111 | rs66507804 | T | C | -0.0322 | 0.00465 | 4.38E-12 | 0.799 | 651 295 | No |
| 112 | rs4269995 | T | C | -0.0339 | 0.00354 | 1.01E-21 | 0.251 | 651 295 | Yes |
| 113 | rs77960 | A | G | 0.0221 | 0.003246 | 9.92E-12 | 0.3304 | 651 295 | No |
| 114 | rs1559253 | A | G | 0.0217 | 0.00355 | 9.82E-10 | 0.3588 | 651 295 | Yes |
| 115 | rs17140201 | A | G | -0.0285 | 0.004745 | 1.90E-09 | 0.172 | 651 295 | No |
| 116 | rs13172141 | A | T | 0.0217 | 0.003443 | 2.94E-10 | 0.5694 | 651 295 | No |
| 117 | rs67988891 | C | G | -0.0363 | 0.003945 | 3.50E-20 | 0.6821 | 651 295 | No |
| 118 | rs2901796 | A | G | 0.0246 | 0.003792 | 8.79E-11 | 0.3983 | 651 295 | No |
| 119 | rs42210 | C | G | -0.0291 | 0.004463 | 6.97E-11 | 0.7124 | 651 295 | No |
| 120 | rs12518401 | A | G | -0.0236 | 0.00352 | 2.01E-11 | 0.3844 | 651 295 | No |
| 121 | rs7735794 | A | G | 0.0344 | 0.005873 | 4.70E-09 | 0.2241 | 651 295 | No |
| 122 | rs465670 | T | C | 0.0241 | 0.003643 | 3.68E-11 | 0.5404 | 651 295 | No |
| 123 | rs9394154 | C | G | -0.0217 | 0.003373 | 1.25E-10 | 0.436 | 651 295 | No |
| 124 | rs9381812 | A | G | -0.0495 | 0.003905 | 8.15E-37 | 0.7051 | 651 295 | Yes |
| 125 | rs1811899 | T | C | -0.0298 | 0.004688 | 2.06E-10 | 0.7895 | 651 295 | No |
| 126 | rs9465253 | T | C | 0.023 | 0.0037 | 5.07E-10 | 0.2801 | 651 295 | No |
| 127 | rs766406 | T | G | -0.0236 | 0.003652 | 1.03E-10 | 0.6409 | 651 295 | Yes |
| 128 | rs486416 | A | G | -0.0201 | 0.003166 | 2.18E-10 | 0.6527 | 651 295 | No |
| 129 | rs13203140 | T | C | -0.0252 | 0.003985 | 2.56E-10 | 0.6353 | 651 295 | No |
| 130 | rs3923809 | A | G | -0.022 | 0.003792 | 6.55E-09 | 0.6947 | 651 295 | No |
| 131 | rs12206814 | C | G | 0.0254 | 0.004249 | 2.26E-09 | 0.4869 | 651 295 | No |
| 132 | rs2396004 | A | G | 0.0214 | 0.003443 | 5.14E-10 | 0.4354 | 651 295 | No |
| 133 | rs3857599 | A | C | 0.0322 | 0.005142 | 3.81E-10 | 0.1659 | 651 295 | No |
| 134 | rs2653349 | A | G | 0.0659 | 0.004304 | 6.58E-53 | 0.2073 | 651 295 | Yes |
| 135 | rs9476310 | T | C | 0.0263 | 0.003502 | 5.90E-14 | 0.5104 | 651 295 | No |
| 136 | rs1931814 | A | G | 0.0264 | 0.003453 | 2.08E-14 | 0.4787 | 651 295 | No |
| 137 | rs2881955 | T | C | 0.0271 | 0.003939 | 5.99E-12 | 0.2788 | 651 295 | No |
| 138 | rs12195792 | A | T | 0.034 | 0.004142 | 2.24E-16 | 0.2717 | 651 295 | Yes |
| 139 | rs11154718 | T | C | -0.0233 | 0.003672 | 2.20E-10 | 0.43 | 651 295 | No |
| 140 | rs60616179 | A | G | 0.0509 | 0.00772 | 4.31E-11 | 0.9443 | 651 295 | No |
| 141 | rs4535583 | T | C | 0.0212 | 0.003674 | 7.92E-09 | 0.6991 | 651 295 | No |
| 142 | rs9496623 | A | G | -0.0235 | 0.004025 | 5.27E-09 | 0.7328 | 651 295 | No |
| 143 | rs2050185 | A | G | 0.0217 | 0.00357 | 1.21E-09 | 0.6245 | 651 295 | No |
| 144 | rs9479402 | T | C | -0.2186 | 0.017396 | 3.26E-36 | 0.9883 | 651 295 | Yes |
| 145 | rs9347926 | A | T | 0.0264 | 0.0034 | 8.16E-15 | 0.4407 | 651 295 | No |
| 146 | rs9348050 | T | C | 0.0221 | 0.003233 | 8.19E-12 | 0.4864 | 651 295 | No |
| 147 | rs4027217 | A | C | -0.0258 | 0.004499 | 9.76E-09 | 0.217 | 651 295 | No |
| 148 | rs10237162 | T | C | 0.0368 | 0.004006 | 4.04E-20 | 0.7232 | 651 295 | Yes |
| 149 | rs10951325 | T | C | 0.0336 | 0.003517 | 1.24E-21 | 0.6319 | 651 295 | Yes |
| 150 | rs6967481 | T | C | 0.0321 | 0.003274 | 1.08E-22 | 0.5011 | 651 295 | Yes |
| 151 | rs4236237 | A | C | -0.0242 | 0.003574 | 1.27E-11 | 0.5992 | 651 295 | No |
| 152 | rs2944831 | A | G | 0.025 | 0.003674 | 1.02E-11 | 0.2946 | 651 295 | No |
| 153 | rs3807651 | A | T | 0.0246 | 0.00408 | 1.64E-09 | 0.4938 | 651 295 | No |
| 154 | rs10254050 | C | G | -0.0577 | 0.00491 | 7.01E-32 | 0.1905 | 651 295 | Yes |
| 155 | rs4729854 | A | T | -0.0485 | 0.003689 | 1.76E-39 | 0.4701 | 651 295 | Yes |
| 156 | rs2396719 | A | G | 0.0331 | 0.004431 | 8.04E-14 | 0.239 | 651 295 | No |
| 157 | rs17302081 | T | C | 0.022 | 0.003642 | 1.53E-09 | 0.4414 | 651 295 | No |
| 158 | rs6968240 | A | C | 0.0216 | 0.003262 | 3.54E-11 | 0.4171 | 651 295 | No |
| 159 | rs62465218 | A | C | -0.0274 | 0.004558 | 1.84E-09 | 0.1456 | 651 295 | No |
| 160 | rs6958557 | T | G | 0.0261 | 0.003591 | 3.62E-13 | 0.6067 | 651 295 | Yes |
| 161 | rs113161209 | A | G | 0.0442 | 0.007651 | 7.59E-09 | 0.077 | 651 295 | No |
| 162 | rs2072413 | T | C | -0.0208 | 0.00364 | 1.11E-08 | 0.2624 | 651 295 | No |
| 163 | rs62479736 | T | G | 0.0243 | 0.003709 | 5.73E-11 | 0.2915 | 651 295 | No |
| 164 | rs35524253 | A | G | 0.0342 | 0.003749 | 7.30E-20 | 0.3539 | 651 295 | Yes |
| 165 | rs2979139 | A | G | -0.0267 | 0.003368 | 2.24E-15 | 0.5048 | 651 295 | No |
| 166 | rs2322605 | A | G | -0.0215 | 0.003438 | 4.03E-10 | 0.4684 | 651 295 | No |
| 167 | rs71523448 | C | G | -0.0499 | 0.006454 | 1.06E-14 | 0.0788 | 651 295 | No |
| 168 | rs6993892 | T | C | -0.0347 | 0.003615 | 8.04E-22 | 0.6162 | 651 295 | Yes |
| 169 | rs6468316 | T | C | -0.0197 | 0.00339 | 6.22E-09 | 0.4708 | 651 295 | No |
| 170 | rs7845620 | A | C | -0.0425 | 0.004577 | 1.62E-20 | 0.8344 | 651 295 | Yes |
| 171 | rs10109566 | A | G | -0.0222 | 0.00368 | 1.62E-09 | 0.4849 | 651 295 | No |
| 172 | rs34054660 | A | G | 0.0249 | 0.004343 | 9.82E-09 | 0.5747 | 651 295 | Yes |
| 173 | rs187028 | A | T | -0.0222 | 0.003336 | 2.83E-11 | 0.3146 | 651 295 | No |
| 174 | rs16939162 | A | G | 0.038 | 0.004701 | 6.26E-16 | 0.8304 | 651 295 | No |
| 175 | rs6988733 | T | C | 0.0229 | 0.003934 | 5.84E-09 | 0.3491 | 651 295 | No |
| 176 | rs7006885 | A | G | 0.0301 | 0.004209 | 8.56E-13 | 0.2878 | 651 295 | Yes |
| 177 | rs3100052 | A | G | 0.0246 | 0.003525 | 2.99E-12 | 0.3886 | 651 295 | No |
| 178 | rs2737245 | T | G | 0.0335 | 0.003778 | 7.56E-19 | 0.2748 | 651 295 | No |
| 179 | rs1871729 | A | G | -0.0232 | 0.003834 | 1.44E-09 | 0.6816 | 651 295 | No |
| 180 | rs6477309 | T | C | 0.0309 | 0.003762 | 2.16E-16 | 0.6661 | 651 295 | Yes |
| 181 | rs2844016 | T | C | 0.0266 | 0.004253 | 3.99E-10 | 0.2935 | 651 295 | No |
| 182 | rs308521 | T | C | 0.0283 | 0.003206 | 1.06E-18 | 0.6039 | 651 295 | No |
| 183 | rs4878734 | A | T | 0.0218 | 0.003808 | 1.03E-08 | 0.5114 | 651 295 | No |
| 184 | rs6560218 | T | C | -0.0224 | 0.003766 | 2.72E-09 | 0.5183 | 651 295 | No |
| 185 | rs62553781 | T | C | -0.0689 | 0.009233 | 8.49E-14 | 0.0329 | 651 295 | No |
| 186 | rs12378543 | T | C | -0.0233 | 0.003797 | 8.43E-10 | 0.3841 | 651 295 | No |
| 187 | rs555784 | A | T | -0.0249 | 0.003777 | 4.33E-11 | 0.3803 | 651 295 | No |
| 188 | rs295268 | T | C | -0.0308 | 0.004596 | 2.05E-11 | 0.7403 | 651 295 | No |
| 189 | rs3138490 | A | T | 0.0235 | 0.003636 | 1.03E-10 | 0.515 | 651 295 | No |
| 190 | rs10759208 | T | C | -0.0248 | 0.003793 | 6.22E-11 | 0.6138 | 651 295 | No |
| 191 | rs11788633 | C | G | 0.0197 | 0.003247 | 1.30E-09 | 0.6547 | 651 295 | No |
| 192 | rs10818834 | T | C | 0.0302 | 0.004168 | 4.30E-13 | 0.7297 | 651 295 | No |
| 193 | rs10988239 | T | C | -0.0213 | 0.003063 | 3.56E-12 | 0.5117 | 651 295 | No |
| 194 | rs12380242 | T | C | -0.0206 | 0.003194 | 1.12E-10 | 0.5058 | 651 295 | No |
| 195 | rs28458909 | T | C | -0.0702 | 0.005531 | 6.65E-37 | 0.1225 | 651 295 | Yes |
| 196 | rs497338 | T | C | 0.0271 | 0.00406 | 2.48E-11 | 0.2881 | 651 295 | No |
| 197 | rs66617308 | T | C | 0.0184 | 0.003207 | 9.62E-09 | 0.6713 | 651 295 | No |
| 198 | rs9416744 | A | C | 0.0344 | 0.003935 | 2.29E-18 | 0.2595 | 651 295 | Yes |
| 199 | rs11597421 | A | G | -0.0238 | 0.004032 | 3.57E-09 | 0.4973 | 651 295 | No |
| 200 | rs12249410 | T | G | -0.0337 | 0.005395 | 4.22E-10 | 0.1077 | 651 295 | No |
| 201 | rs17712705 | A | G | -0.0247 | 0.003989 | 5.94E-10 | 0.3265 | 651 295 | No |
| 202 | rs2298117 | T | C | -0.0225 | 0.003653 | 7.31E-10 | 0.448 | 651 295 | No |
| 203 | rs10762434 | C | G | 0.0249 | 0.004246 | 4.50E-09 | 0.7752 | 651 295 | No |
| 204 | rs2648721 | T | G | -0.0238 | 0.004061 | 4.60E-09 | 0.7039 | 651 295 | No |
| 205 | rs61875203 | T | C | 0.0259 | 0.004118 | 3.19E-10 | 0.2773 | 651 295 | No |
| 206 | rs1163238 | A | G | -0.0237 | 0.004099 | 7.36E-09 | 0.3885 | 651 295 | No |
| 207 | rs7900191 | T | C | -0.0187 | 0.003269 | 1.07E-08 | 0.4035 | 651 295 | No |
| 208 | rs11200159 | A | C | -0.0234 | 0.003798 | 7.23E-10 | 0.6551 | 651 295 | No |
| 209 | rs3808964 | T | G | 0.02 | 0.00336 | 2.63E-09 | 0.6326 | 651 295 | No |
| 210 | rs9664044 | T | C | -0.0268 | 0.004263 | 3.26E-10 | 0.2319 | 651 295 | No |
| 211 | rs10830107 | A | G | 0.0283 | 0.004606 | 8.07E-10 | 0.7922 | 651 295 | No |
| 212 | rs76518095 | T | C | 0.0398 | 0.006579 | 1.45E-09 | 0.0772 | 651 295 | No |
| 213 | rs12771973 | A | G | -0.0223 | 0.0038 | 4.41E-09 | 0.2475 | 651 295 | No |
| 214 | rs10832648 | A | C | -0.0314 | 0.004374 | 7.03E-13 | 0.1989 | 651 295 | No |
| 215 | rs10742179 | A | G | 0.0352 | 0.004264 | 1.52E-16 | 0.2627 | 651 295 | Yes |
| 216 | rs4923541 | T | C | 0.0247 | 0.003807 | 8.69E-11 | 0.5104 | 651 295 | No |
| 217 | rs621421 | T | C | -0.0273 | 0.003409 | 1.17E-15 | 0.6256 | 651 295 | No |
| 218 | rs11032362 | A | G | 0.0704 | 0.006162 | 3.17E-30 | 0.0937 | 651 295 | Yes |
| 219 | rs7111582 | A | G | -0.0388 | 0.004898 | 2.36E-15 | 0.8956 | 651 295 | No |
| 220 | rs10838687 | T | G | 0.0345 | 0.004475 | 1.26E-14 | 0.7918 | 651 295 | Yes |
| 221 | rs12808544 | A | C | -0.0348 | 0.00421 | 1.37E-16 | 0.2409 | 651 295 | No |
| 222 | rs662094 | A | G | 0.0282 | 0.00373 | 4.01E-14 | 0.4944 | 651 295 | Yes |
| 223 | rs1278402 | A | G | 0.0281 | 0.004492 | 3.96E-10 | 0.736 | 651 295 | Yes |
| 224 | rs1508608 | A | G | 0.028 | 0.003734 | 6.42E-14 | 0.3199 | 651 295 | Yes |
| 225 | rs4121878 | C | G | 0.0219 | 0.003628 | 1.57E-09 | 0.5008 | 651 295 | No |
| 226 | rs17577073 | A | C | 0.0246 | 0.003691 | 2.64E-11 | 0.5636 | 651 295 | No |
| 227 | rs2514214 | A | G | 0.027 | 0.004294 | 3.22E-10 | 0.2691 | 651 295 | No |
| 228 | rs4936290 | A | C | -0.0229 | 0.0034 | 1.64E-11 | 0.6584 | 651 295 | No |
| 229 | rs3867239 | A | G | 0.0262 | 0.003725 | 2.02E-12 | 0.3762 | 651 295 | No |
| 230 | rs74357745 | A | G | 0.0312 | 0.004756 | 5.39E-11 | 0.8803 | 651 295 | No |
| 231 | rs7943634 | T | C | -0.0235 | 0.003655 | 1.28E-10 | 0.3075 | 651 295 | No |
| 232 | rs3782860 | T | C | 0.025 | 0.003646 | 7.05E-12 | 0.543 | 651 295 | No |
| 233 | rs1799464 | A | G | -0.02 | 0.003439 | 6.01E-09 | 0.2868 | 651 295 | No |
| 234 | rs12298405 | T | C | -0.0234 | 0.003495 | 2.16E-11 | 0.3301 | 651 295 | No |
| 235 | rs2433634 | A | C | -0.0268 | 0.004102 | 6.44E-11 | 0.7241 | 651 295 | No |
| 236 | rs11611435 | T | C | 0.0278 | 0.004063 | 7.75E-12 | 0.5561 | 651 295 | No |
| 237 | rs13377754 | T | C | 0.0485 | 0.003484 | 4.85E-44 | 0.6104 | 651 295 | Yes |
| 238 | rs1843888 | A | G | 0.0513 | 0.003546 | 1.89E-47 | 0.5433 | 651 295 | Yes |
| 239 | rs247929 | C | G | 0.031 | 0.003526 | 1.47E-18 | 0.5081 | 651 295 | Yes |
| 240 | rs7975791 | T | C | 0.0513 | 0.008373 | 8.94E-10 | 0.0384 | 651 295 | No |
| 241 | rs4761989 | T | C | -0.0333 | 0.005161 | 1.11E-10 | 0.868 | 651 295 | No |
| 242 | rs7299922 | A | G | 0.0239 | 0.003476 | 6.17E-12 | 0.6388 | 651 295 | No |
| 243 | rs487722 | T | G | 0.0273 | 0.004331 | 2.91E-10 | 0.2084 | 651 295 | No |
| 244 | rs10877962 | T | C | 0.0359 | 0.003943 | 8.59E-20 | 0.4083 | 651 295 | Yes |
| 245 | rs711098 | A | C | 0.0217 | 0.00356 | 1.10E-09 | 0.3979 | 651 295 | Yes |
| 246 | rs7959983 | T | C | -0.0299 | 0.003512 | 1.68E-17 | 0.5955 | 651 295 | No |
| 247 | rs7304278 | A | G | -0.0291 | 0.003613 | 7.97E-16 | 0.2827 | 651 295 | No |
| 248 | rs7298532 | T | C | 0.027 | 0.004016 | 1.79E-11 | 0.7213 | 651 295 | No |
| 249 | rs3955311 | T | C | 0.026 | 0.004464 | 5.71E-09 | 0.1491 | 651 295 | No |
| 250 | rs80097534 | T | G | -0.0357 | 0.00573 | 4.66E-10 | 0.0966 | 651 295 | No |
| 251 | rs9597241 | A | C | 0.0332 | 0.004252 | 5.83E-15 | 0.8106 | 651 295 | No |
| 252 | rs12871550 | A | G | 0.0268 | 0.003612 | 1.18E-13 | 0.3229 | 651 295 | No |
| 253 | rs9571526 | T | G | -0.0272 | 0.004442 | 9.15E-10 | 0.7706 | 651 295 | No |
| 254 | rs2593487 | A | G | -0.0285 | 0.003867 | 1.70E-13 | 0.3361 | 651 295 | No |
| 255 | rs495593 | A | G | 0.0226 | 0.003656 | 6.32E-10 | 0.7383 | 651 295 | No |
| 256 | rs45597035 | A | G | -0.0222 | 0.003774 | 4.06E-09 | 0.6469 | 651 295 | No |
| 257 | rs9573980 | A | G | 0.1265 | 0.009631 | 2.10E-39 | 0.9659 | 651 295 | Yes |
| 258 | rs1886205 | A | C | 0.0293 | 0.003726 | 3.73E-15 | 0.7604 | 651 295 | No |
| 259 | rs9558942 | T | C | -0.0192 | 0.003347 | 9.70E-09 | 0.6722 | 651 295 | No |
| 260 | rs3815983 | T | C | -0.0216 | 0.003299 | 5.83E-11 | 0.3604 | 651 295 | No |
| 261 | rs1163628 | A | C | -0.0287 | 0.004639 | 6.14E-10 | 0.8572 | 651 295 | No |
| 262 | rs61990287 | A | C | 0.0248 | 0.004068 | 1.09E-09 | 0.2742 | 651 295 | No |
| 263 | rs2878172 | A | G | -0.0213 | 0.003508 | 1.27E-09 | 0.5693 | 651 295 | No |
| 264 | rs962961 | T | C | -0.0219 | 0.003169 | 4.82E-12 | 0.3308 | 651 295 | No |
| 265 | rs6573308 | T | C | 0.0248 | 0.004136 | 2.02E-09 | 0.3941 | 651 295 | No |
| 266 | rs7143933 | T | G | 0.0246 | 0.004045 | 1.19E-09 | 0.2615 | 651 295 | No |
| 267 | rs2978382 | T | C | 0.0232 | 0.003814 | 1.18E-09 | 0.5852 | 651 295 | No |
| 268 | rs4903203 | A | G | 0.0247 | 0.003688 | 2.13E-11 | 0.3237 | 651 295 | No |
| 269 | rs12436039 | T | C | 0.0332 | 0.005718 | 6.40E-09 | 0.8801 | 651 295 | No |
| 270 | rs4550384 | T | G | 0.0237 | 0.003751 | 2.63E-10 | 0.758 | 651 295 | No |
| 271 | rs710284 | T | C | 0.022 | 0.00373 | 3.67E-09 | 0.5797 | 651 295 | No |
| 272 | rs11845599 | A | G | -0.0274 | 0.003621 | 3.83E-14 | 0.6353 | 651 295 | No |
| 273 | rs59986227 | C | G | -0.0307 | 0.004334 | 1.40E-12 | 0.7493 | 651 295 | No |
| 274 | rs12442008 | T | C | 0.0285 | 0.00414 | 5.83E-12 | 0.2593 | 651 295 | No |
| 275 | rs4775086 | A | G | -0.0267 | 0.004472 | 2.37E-09 | 0.2363 | 651 295 | No |
| 276 | rs12442674 | A | C | 0.0229 | 0.003788 | 1.49E-09 | 0.7296 | 651 295 | No |
| 277 | rs1873958 | A | G | 0.0279 | 0.003361 | 1.04E-16 | 0.4088 | 651 295 | Yes |
| 278 | rs72773411 | A | G | 0.0293 | 0.005032 | 5.78E-09 | 0.1529 | 651 295 | No |
| 279 | rs12445235 | C | G | -0.0211 | 0.003546 | 2.69E-09 | 0.4098 | 651 295 | No |
| 280 | rs2304467 | C | G | -0.0237 | 0.003901 | 1.24E-09 | 0.6066 | 651 295 | No |
| 281 | rs11641239 | T | C | 0.0229 | 0.003729 | 8.20E-10 | 0.2854 | 651 295 | No |
| 282 | rs7203707 | A | C | -0.0197 | 0.003022 | 7.12E-11 | 0.5193 | 651 295 | No |
| 283 | rs4785296 | C | G | 0.0264 | 0.004157 | 2.14E-10 | 0.231 | 651 295 | No |
| 284 | rs3743794 | A | G | -0.0215 | 0.003534 | 1.18E-09 | 0.6073 | 651 295 | No |
| 285 | rs12927162 | A | G | 0.0561 | 0.004154 | 1.48E-41 | 0.7276 | 651 295 | Yes |
| 286 | rs1421085 | T | C | -0.0419 | 0.003392 | 4.65E-35 | 0.5935 | 651 295 | Yes |
| 287 | rs2550298 | T | C | -0.04 | 0.003621 | 2.28E-28 | 0.3811 | 651 295 | Yes |
| 288 | rs8044054 | T | C | 0.0307 | 0.003509 | 2.18E-18 | 0.3878 | 651 295 | No |
| 289 | rs72790386 | T | G | 0.0604 | 0.010279 | 4.20E-09 | 0.0338 | 651 295 | No |
| 290 | rs17604349 | A | G | -0.0374 | 0.003972 | 4.62E-21 | 0.1848 | 651 295 | Yes |
| 291 | rs1061032 | T | G | 0.0644 | 0.006151 | 1.19E-25 | 0.094 | 651 295 | Yes |
| 292 | rs11545787 | A | G | -0.0498 | 0.004168 | 6.66E-33 | 0.2482 | 651 295 | Yes |
| 293 | rs12950382 | A | G | 0.0234 | 0.004087 | 1.03E-08 | 0.7198 | 651 295 | No |
| 294 | rs4365329 | A | T | -0.0194 | 0.003328 | 5.59E-09 | 0.5383 | 651 295 | No |
| 295 | rs2011528 | T | C | -0.0329 | 0.004728 | 3.45E-12 | 0.8254 | 651 295 | No |
| 296 | rs3760381 | A | G | 0.0274 | 0.004176 | 5.33E-11 | 0.2544 | 651 295 | No |
| 297 | rs7225002 | A | G | -0.0179 | 0.003124 | 1.00E-08 | 0.5916 | 651 295 | No |
| 298 | rs12600452 | A | G | 0.0255 | 0.004065 | 3.53E-10 | 0.2015 | 651 295 | No |
| 299 | rs12051 | A | G | -0.0264 | 0.003484 | 3.54E-14 | 0.6119 | 651 295 | No |
| 300 | rs55846845 | A | G | -0.021 | 0.003045 | 5.34E-12 | 0.5177 | 651 295 | No |
| 301 | rs72829706 | A | G | 0.0563 | 0.008348 | 1.54E-11 | 0.96 | 651 295 | No |
| 302 | rs8072058 | A | T | -0.0281 | 0.004658 | 1.61E-09 | 0.7812 | 651 295 | No |
| 303 | rs412000 | C | G | -0.0224 | 0.003546 | 2.67E-10 | 0.555 | 651 295 | No |
| 304 | rs58681483 | A | G | 0.0352 | 0.005855 | 1.83E-09 | 0.918 | 651 295 | No |
| 305 | rs72841368 | A | T | -0.0301 | 0.004301 | 2.60E-12 | 0.8104 | 651 295 | No |
| 306 | rs2916148 | A | G | 0.0279 | 0.003505 | 1.71E-15 | 0.4522 | 651 295 | No |
| 307 | rs2580160 | A | G | 0.0283 | 0.004086 | 4.32E-12 | 0.5554 | 651 295 | No |
| 308 | rs62082402 | T | G | 0.05 | 0.005375 | 1.36E-20 | 0.1936 | 651 295 | No |
| 309 | rs1788784 | A | G | -0.0273 | 0.004157 | 5.15E-11 | 0.3442 | 651 295 | Yes |
| 310 | rs1013987 | T | C | -0.0291 | 0.003851 | 4.11E-14 | 0.4033 | 651 295 | No |
| 311 | rs4419127 | A | G | 0.0444 | 0.003735 | 1.35E-32 | 0.6622 | 651 295 | Yes |
| 312 | rs9950528 | A | G | -0.0235 | 0.003819 | 7.57E-10 | 0.6505 | 651 295 | No |
| 313 | rs12969848 | T | C | 0.0355 | 0.003539 | 1.12E-23 | 0.5313 | 651 295 | Yes |
| 314 | rs9956387 | A | T | -0.0198 | 0.003397 | 5.57E-09 | 0.4953 | 651 295 | No |
| 315 | rs4800998 | A | T | 0.0394 | 0.004984 | 2.69E-15 | 0.1827 | 651 295 | Yes |
| 316 | rs9964420 | A | C | -0.0488 | 0.003645 | 6.97E-41 | 0.2983 | 651 295 | Yes |
| 317 | rs11152350 | A | C | -0.0281 | 0.003615 | 7.63E-15 | 0.467 | 651 295 | No |
| 318 | rs34329963 | T | C | -0.0318 | 0.005328 | 2.39E-09 | 0.113 | 651 295 | No |
| 319 | rs1025601 | T | C | -0.0218 | 0.00374 | 5.57E-09 | 0.3855 | 651 295 | No |
| 320 | rs10402849 | T | C | 0.0259 | 0.004165 | 5.04E-10 | 0.2002 | 651 295 | No |
| 321 | rs36055559 | A | G | -0.036 | 0.00501 | 6.69E-13 | 0.166 | 651 295 | No |
| 322 | rs7248205 | T | C | 0.0267 | 0.003572 | 7.78E-14 | 0.6003 | 651 295 | No |
| 323 | rs9636202 | A | G | -0.026 | 0.003749 | 4.06E-12 | 0.2662 | 651 295 | No |
| 324 | rs73026775 | A | G | -0.0335 | 0.005709 | 4.41E-09 | 0.1246 | 651 295 | No |
| 325 | rs4804951 | A | G | 0.0234 | 0.003806 | 7.83E-10 | 0.3306 | 651 295 | No |
| 326 | rs56113850 | T | C | -0.0229 | 0.003449 | 3.15E-11 | 0.4231 | 651 295 | No |
| 327 | rs58876439 | A | G | 0.0474 | 0.006806 | 3.31E-12 | 0.0674 | 651 295 | No |
| 328 | rs11670534 | T | C | -0.0305 | 0.004689 | 7.83E-11 | 0.1614 | 651 295 | No |
| 329 | rs6131805 | T | G | 0.0257 | 0.003643 | 1.72E-12 | 0.4022 | 651 295 | No |
| 330 | rs6131942 | A | G | -0.0263 | 0.003112 | 2.89E-17 | 0.418 | 651 295 | Yes |
| 331 | rs1474754 | A | G | -0.0214 | 0.003623 | 3.51E-09 | 0.2636 | 651 295 | No |
| 332 | rs6047481 | A | T | 0.0253 | 0.004084 | 5.81E-10 | 0.6709 | 651 295 | No |
| 333 | rs1737893 | T | C | -0.0253 | 0.003728 | 1.15E-11 | 0.3805 | 651 295 | No |
| 334 | rs2072727 | T | C | 0.0282 | 0.003427 | 1.89E-16 | 0.4327 | 651 295 | Yes |
| 335 | rs57236847 | C | G | 0.0269 | 0.004373 | 7.69E-10 | 0.6029 | 651 295 | No |
| 336 | rs695459 | T | C | -0.022 | 0.00368 | 2.25E-09 | 0.3906 | 651 295 | No |
| 337 | rs28459838 | T | C | 0.0266 | 0.004183 | 2.03E-10 | 0.2369 | 651 295 | No |
| 338 | rs118047999 | C | G | 0.0236 | 0.003982 | 3.10E-09 | 0.2459 | 651 295 | No |
| 339 | rs139911 | T | C | -0.0336 | 0.003644 | 2.94E-20 | 0.5722 | 651 295 | No |
| 340 | rs9611597 | A | T | 0.0374 | 0.005098 | 2.20E-13 | 0.8371 | 651 295 | No |
| 341 | rs6007594 | A | G | -0.025 | 0.003859 | 9.26E-11 | 0.2644 | 651 295 | No |

Table G3: Summary information of genetic variants identified for sleep duration.

| Sr. No. | rsID | Effect Allele | Other Allele | Beta | Standard Error | P | Effect Allele Frequency | N |
| --- | --- | --- | --- | --- | --- | --- | --- | --- |
| 1 | rs7556815 | A | G | 2.443 | 0.164 | 1.3E-49 | 0.219 | 446 118 |
| 2 | rs75539574 | C | A | 2.175 | 0.244 | 6.9E-19 | 0.086 | 446 118 |
| 3 | rs12607679 | T | C | 1.208 | 0.156 | 8.3E-15 | 0.738 | 446 118 |
| 4 | rs915416 | C | G | 1.156 | 0.150 | 9.9E-15 | 0.290 | 446 118 |
| 5 | rs9940646 | C | G | 1.017 | 0.137 | 1.2E-13 | 0.578 | 446 118 |
| 6 | rs13109404 | T | G | 1.872 | 0.264 | 1.4E-12 | 0.928 | 446 118 |
| 7 | rs8050478 | G | A | 0.960 | 0.136 | 1.7E-12 | 0.500 | 446 118 |
| 8 | rs56372231 | T | C | 1.017 | 0.144 | 2.2E-12 | 0.334 | 446 118 |
| 9 | rs13088093 | G | T | 0.976 | 0.144 | 7E-12 | 0.336 | 446 118 |
| 10 | rs2079070 | C | G | 1.053 | 0.154 | 7.5E-12 | 0.265 | 446 118 |
| 11 | rs34556183 | A | G | 1.015 | 0.151 | 2.3E-11 | 0.720 | 446 118 |
| 12 | rs3095508 | C | A | 0.921 | 0.138 | 3.1E-11 | 0.594 | 446 118 |
| 13 | rs34731055 | T | C | 1.168 | 0.177 | 3.7E-11 | 0.181 | 446 118 |
| 14 | rs73219758 | G | A | 0.984 | 0.150 | 5.6E-11 | 0.708 | 446 118 |
| 15 | rs10973207 | T | G | 1.226 | 0.187 | 6E-11 | 0.158 | 446 118 |
| 16 | rs2139261 | G | C | 1.122 | 0.174 | 8.50E-11 | 0.749 | 446 118 |
| 17 | rs4592416 | G | A | 0.881 | 0.136 | 9.3E-11 | 0.464 | 446 118 |
| 18 | rs365663 | A | G | 0.878 | 0.137 | 1E-10 | 0.546 | 446 118 |
| 19 | rs1517572 | C | A | 0.879 | 0.138 | 1.5E-10 | 0.581 | 446 118 |
| 20 | rs7915425 | T | C | 1.144 | 0.179 | 2E-10 | 0.175 | 446 118 |
| 21 | rs330088 | C | T | 0.868 | 0.137 | 2.7E-10 | 0.547 | 446 118 |
| 22 | rs8038326 | A | G | 0.955 | 0.152 | 2.8E-10 | 0.727 | 446 118 |
| 23 | rs460692 | C | T | 1.263 | 0.200 | 3.6E-10 | 0.137 | 446 118 |
| 24 | rs9382445 | T | C | 0.872 | 0.140 | 4.8E-10 | 0.623 | 446 118 |
| 25 | rs4767550 | G | A | 0.858 | 0.139 | 6.3E-10 | 0.414 | 446 118 |
| 26 | rs11885663 | T | C | 0.973 | 0.157 | 8.6E-10 | 0.248 | 446 118 |
| 27 | rs1991556 | G | A | 0.994 | 0.163 | 1.00E-09 | 0.774 | 446 118 |
| 28 | rs1057703 | G | T | 1.164 | 0.192 | 1.1E-09 | 0.147 | 446 118 |
| 29 | rs4128364 | C | T | 0.876 | 0.143 | 1.4E-09 | 0.339 | 446 118 |
| 30 | rs61796569 | T | C | 0.927 | 0.154 | 1.5E-09 | 0.270 | 446 118 |
| 31 | rs10483350 | G | A | 1.042 | 0.172 | 1.5E-09 | 0.195 | 446 118 |
| 32 | rs7115226 | A | C | 1.594 | 0.261 | 1.7E-09 | 0.074 | 446 118 |
| 33 | rs269054 | A | T | 0.819 | 0.138 | 2.1E-09 | 0.422 | 446 118 |
| 34 | rs112230981 | A | G | 1.892 | 0.314 | 2.2E-09 | 0.950 | 446 118 |
| 35 | rs11602180 | C | T | 1.095 | 0.184 | 2.3E-09 | 0.837 | 446 118 |
| 36 | rs2192528 | A | G | 0.802 | 0.136 | 2.7E-09 | 0.480 | 446 118 |
| 37 | rs12246842 | A | G | 0.804 | 0.136 | 3.9E-09 | 0.460 | 446 118 |
| 38 | rs205024 | T | C | 0.830 | 0.140 | 3.9E-09 | 0.384 | 446 118 |
| 39 | rs12567114 | A | G | 0.890 | 0.152 | 4.3E-09 | 0.276 | 446 118 |
| 40 | rs7616632 | T | G | 0.792 | 0.136 | 4.3E-09 | 0.522 | 446 118 |
| 41 | rs6575005 | T | C | 0.934 | 0.159 | 4.4E-09 | 0.758 | 446 118 |
| 42 | rs1776776 | T | C | 1.198 | 0.205 | 4.9E-09 | 0.874 | 446 118 |
| 43 | rs11621908 | C | T | 1.446 | 0.250 | 5.6E-09 | 0.917 | 446 118 |
| 44 | rs10421649 | A | T | 0.798 | 0.138 | 6.9E-09 | 0.557 | 446 118 |
| 45 | rs2072727 | T | C | 0.795 | 0.137 | 7.9E-09 | 0.436 | 446 118 |
| 46 | rs113113059 | T | C | 0.968 | 0.164 | 8.4E-09 | 0.780 | 446 118 |
| 47 | rs374153 | C | T | 1.057 | 0.186 | 9.1E-09 | 0.158 | 446 118 |
| 48 | rs151014368 | A | G | 0.966 | 0.169 | 9.1E-09 | 0.206 | 446 118 |
| 49 | rs62120041 | T | C | 1.567 | 0.274 | 9.6E-09 | 0.934 | 446 118 |
| 50 | rs7503199 | C | T | 0.885 | 0.154 | 1.00E-08 | 0.734 | 446 118 |
| 51 | rs1939455 | G | T | 1.226 | 0.214 | 1.20E-08 | 0.879 | 446 118 |
| 52 | rs7951019 | G | T | 2.213 | 0.391 | 1.20E-08 | 0.032 | 446 118 |
| 53 | rs17732997 | C | G | 0.776 | 0.137 | 1.20E-08 | 0.569 | 446 118 |
| 54 | rs61985058 | T | C | 1.116 | 0.194 | 1.30E-08 | 0.143 | 446 118 |
| 55 | rs17427571 | A | G | 0.830 | 0.146 | 1.30E-08 | 0.684 | 446 118 |
| 56 | rs7806045 | T | C | 0.887 | 0.158 | 1.40E-08 | 0.755 | 446 118 |
| 57 | rs35531607 | C | T | 0.770 | 0.136 | 1.50E-08 | 0.474 | 446 118 |
| 58 | rs7644809 | T | C | 0.784 | 0.138 | 1.60E-08 | 0.422 | 446 118 |
| 59 | rs9345234 | C | A | 0.781 | 0.138 | 1.80E-08 | 0.578 | 446 118 |
| 60 | rs12791153 | T | A | 1.413 | 0.253 | 1.90E-08 | 0.081 | 446 118 |
| 61 | rs1263056 | A | G | 0.768 | 0.137 | 2.00E-08 | 0.519 | 446 118 |
| 62 | rs55658675 | C | T | 0.788 | 0.142 | 2.00E-08 | 0.645 | 446 118 |
| 63 | rs11567976 | T | C | 0.768 | 0.137 | 2.10E-08 | 0.571 | 446 118 |
| 64 | rs180769 | T | C | 0.763 | 0.138 | 2.30E-08 | 0.425 | 446 118 |
| 65 | rs1553132 | G | A | 0.870 | 0.155 | 2.50E-08 | 0.258 | 446 118 |
| 66 | rs9903973 | C | T | 0.766 | 0.136 | 2.60E-08 | 0.467 | 446 118 |
| 67 | rs11614986 | A | G | 0.983 | 0.177 | 2.70E-08 | 0.821 | 446 118 |
| 68 | rs2231265 | G | A | 0.897 | 0.162 | 2.70E-08 | 0.772 | 446 118 |
| 69 | rs174560 | C | T | 0.815 | 0.146 | 2.80E-08 | 0.314 | 446 118 |
| 70 | rs10173260 | C | T | 0.770 | 0.139 | 2.90E-08 | 0.606 | 446 118 |
| 71 | rs72804080 | G | A | 1.068 | 0.192 | 2.90E-08 | 0.150 | 446 118 |
| 72 | rs12611523 | A | G | 0.758 | 0.137 | 3.10E-08 | 0.545 | 446 118 |
| 73 | rs11643715 | G | C | 0.834 | 0.150 | 3.20E-08 | 0.291 | 446 118 |
| 74 | rs4538155 | T | C | 0.779 | 0.142 | 3.60E-08 | 0.647 | 446 118 |
| 75 | rs34354917 | C | A | 0.825 | 0.150 | 3.90E-08 | 0.710 | 446 118 |
| 76 | rs80193650 | G | A | 1.010 | 0.184 | 4.10E-08 | 0.162 | 446 118 |
| 77 | rs10761674 | C | T | 0.740 | 0.136 | 4.20E-08 | 0.477 | 446 118 |
| 78 | rs11190970 | G | A | 0.923 | 0.169 | 4.60E-08 | 0.799 | 446 118 |

Table G4: Summary information of genetic variants identified for short sleep.

| Sr. No. | rsID | Effect Allele | Other Allele | Beta | Standard Error | P | Effect Allele Frequency | N |
| --- | --- | --- | --- | --- | --- | --- | --- | --- |
| 1 | rs2863957 | C | A | 0.05449 | 0.0072 | 2.60E-18 | 0.782 | 411 934 |
| 2 | rs13107325 | T | C | 0.07511 | 0.0109 | 2.50E-13 | 0.075 | 411 934 |
| 3 | rs1229762 | T | C | 0.03730 | 0.0064 | 1.10E-12 | 0.665 | 411 934 |
| 4 | rs1380703 | G | A | 0.03537 | 0.0062 | 1.60E-11 | 0.384 | 411 934 |
| 5 | rs12963463 | C | T | 0.02859 | 0.0064 | 1.90E-11 | 0.299 | 411 934 |
| 6 | rs75539574 | A | C | 0.04497 | 0.0107 | 8.40E-11 | 0.915 | 411 934 |
| 7 | rs17388803 | C | A | 0.05259 | 0.0097 | 6.50E-10 | 0.106 | 411 934 |
| 8 | rs4585442 | G | A | 0.03053 | 0.0062 | 8.10E-10 | 0.311 | 411 934 |
| 9 | rs1607227 | G | T | 0.03053 | 0.0067 | 1.50E-09 | 0.705 | 411 934 |
| 10 | rs2820313 | G | A | 0.03053 | 0.0059 | 2.30E-09 | 0.341 | 411 934 |
| 11 | rs17005118 | A | G | 0.02956 | 0.0067 | 2.50E-09 | 0.265 | 411 934 |
| 12 | rs5757675 | G | T | 0.03440 | 0.0067 | 2.70E-09 | 0.260 | 411 934 |
| 13 | rs12567114 | G | A | 0.03633 | 0.0066 | 4.10E-09 | 0.725 | 411 934 |
| 14 | rs142180737 | C | T | 0.15444 | 0.0317 | 4.40E-09 | 0.009 | 411 934 |
| 15 | rs2186122 | T | A | 0.02372 | 0.0060 | 4.80E-09 | 0.562 | 411 934 |
| 16 | rs11763750 | G | A | 0.03537 | 0.0076 | 5.10E-09 | 0.814 | 411 934 |
| 17 | rs12518468 | C | T | 0.03150 | 0.0062 | 8.50E-09 | 0.328 | 411 934 |
| 18 | rs9367621 | T | A | 0.02372 | 0.0057 | 1.60E-08 | 0.431 | 411 934 |
| 19 | rs3776864 | A | C | 0.03150 | 0.0064 | 1.70E-08 | 0.667 | 411 934 |
| 20 | rs60882754 | A | T | 0.05543 | 0.0123 | 1.80E-08 | 0.939 | 411 934 |
| 21 | rs59779556 | T | G | 0.02469 | 0.0060 | 2.00E-08 | 0.554 | 411 934 |
| 22 | rs2014830 | C | T | 0.02956 | 0.0064 | 2.70E-08 | 0.698 | 411 934 |
| 23 | rs205024 | C | T | 0.03053 | 0.0062 | 2.70E-08 | 0.617 | 411 934 |
| 24 | rs12661667 | T | C | 0.02762 | 0.0067 | 2.80E-08 | 0.263 | 411 934 |
| 25 | rs7939345 | T | G | 0.03537 | 0.0071 | 4.00E-08 | 0.208 | 411 934 |
| 26 | rs9321171 | C | T | 0.03150 | 0.0059 | 4.20E-08 | 0.540 | 411 934 |
| 27 | rs7524118 | C | T | 0.02956 | 0.0064 | 4.90E-08 | 0.708 | 411 934 |

Table G5: Summary information of genetic variants identified for long sleep.

| Sr. No. | rsID | Effect Allele | Other Allele | Beta | Standard Error | P | Effect Allele Frequency | N |
| --- | --- | --- | --- | --- | --- | --- | --- | --- |
| 1 | rs6737318 | G | A | 0.07603 | 0.0111 | 3.40E-13 | 0.222 | 339 926 |
| 2 | rs75458655 | T | C | 0.18482 | 0.0294 | 5.40E-12 | 0.023 | 339 926 |
| 3 | rs17688916 | T | A | 0.07139 | 0.0124 | 1.10E-11 | 0.796 | 339 926 |
| 4 | rs17817288 | A | G | 0.03922 | 0.0093 | 8.90E-09 | 0.518 | 339 926 |
| 5 | rs549961083 | T | C | 0.53357 | 0.1166 | 9.60E-09 | 0.001 | 339 926 |
| 6 | rs3751046 | G | A | 0.06953 | 0.0131 | 2.00E-08 | 0.147 | 339 926 |
| 7 | rs7534398 | A | T | 0.04688 | 0.0117 | 2.10E-08 | 0.201 | 339 926 |
| 8 | rs10899257 | A | G | 0.06766 | 0.0131 | 4.60E-08 | 0.144 | 339 926 |
